# Preventing antimalarial drug resistance with triple artemisinin-based combination therapies

**DOI:** 10.1101/2022.10.21.22281347

**Authors:** Tran Dang Nguyen, Bo Gao, Chanaki Amaratunga, Mehul Dhorda, Thu Nguyen-Anh Tran, Nicholas J White, Arjen M Dondorp, Maciej F Boni, Ricardo Aguas

## Abstract

**Background:** Increasing levels of artemisinin and partner drug resistance threaten malaria control and elimination globally. Triple artemisinin-based combination therapies (TACTs) which combine artemisinin derivatives with two partner drugs are efficacious and well tolerated in clinical trials, including in areas of multidrug-resistant malaria. Whether early TACT adoption could delay the emergence and spread of antimalarial drug resistance is a question of vital importance.

**Methods:** Using two independent individual-based models of *Plasmodium falciparum* epidemiology and evolution, we evaluated whether introduction of either artesunate-mefloquine-piperaquine or artemether-lumefantrine-amodiaquine resulted in lower long-term artemisinin-resistance levels and treatment failure rates compared with continued ACT use.

**Findings:** In countries with 1% *P. falciparum* prevalence, immediate adoption of TACTs would result in substantially lower frequency of artemisinin-resistant alleles 10 years later. Median estimates were 70%, 33%, and 18% lower allele frequency for countries currently deploying dihydroartemisinin-piperaquine, artesunate-amodiaquine, or artemether-lumefantrine, respectively. Corresponding median treatment failure rate decreases are 74%, 34%, and 17%. Delaying TACT introduction increases future resistance frequencies and treatment failure rates. The most significant threat to the success of TACTs is the emergence of a triple-resistant genotype. which if above 0.01 frequency may undermine elimination efforts in low-prevalence regions.

**Interpretation:** Introduction of TACTs could delay the emergence and spread of artemisinin resistance and treatment failure, extending the useful therapeutic life of current antimalarial drugs and improving the chances of malaria elimination. Immediate introduction of TACTs should be considered by policy makers in areas of emerging artemisinin resistance.

## Introduction

The introduction of artemisinin-based combination therapies (ACTs) into routine clinical use for uncomplicated *Plasmodium falciparum* malaria has saved millions of lives over the past two decades^1^. These drugs are the mainstay of antimalarial treatment and remain highly effective in most malaria endemic regions^2^. However, resistance to artemisinins has emerged in several malaria endemic regions, first in the Greater Mekong Subregion (GMS) of Southeast Asia^3-5^, and subsequently in South America^6^, Papua New Guinea^7^, and most recently in Eastern Africa^8,9^. In the GMS, artemisinin resistance was compounded by ACT partner drug resistance, causing ACT treatment failure ^10,11^. Increasing drug resistance threatens global malaria control and elimination efforts. Artemisinin partial resistance, caused by mutations in the *pfkelch13* gene^12,13^, results in slower parasite clearance, increased rates of treatment failure, increased transmissibility, and reduced protection of the partner drugs from resistance emergence and spread. The recent emergence of artemisinin resistance in East Africa is particularly concerning as the previous reduction in the malaria burden has stalled in many parts of the continent. Malaria mortality, which fell substantially between 2000 and 2015, is estimated to have plateaued over the past seven years^2^. Worsening drug resistance will result in an increasing death toll, with most of these preventable deaths occurring in African children.

The current high efficacy of antimalarial treatments must be maintained if a lethal reversal in malaria trends is to be avoided. New antimalarial drugs are promised^14^, but even if their development continues successfully, they may not become available for years and will be more expensive^15^. Multiple first-line therapies (MFT), which mathematical modelling suggests can slow drug resistance spread ^16,17^, has become policy in more than a dozen endemic countries^2^. However, there is yet no field evidence demonstrating the success of MFT at slowing or delaying resistance evolution. A potential alternative solution, based on the same biological principles of combining two different slowly eliminated antimalarial drugs with an artemisinin derivative, are triple artemisinin-based combinations (TACTs), which should provide protection against partner drug resistance and therefore preserve high treatment efficacy, given the extreme rarity of parasites acquiring mutations conferring resistance against both partner drugs over a short period of time. Randomised clinical trials in Asia with dihydroartemisinin-piperaquine-mefloquine (DHA-PPQ-MQ) and artemether-lumefantrine-amodiaquine (ALAQ) have shown that these combinations are well tolerated, safe and effective, including in areas with multidrug-resistant falciparum malaria^18,19^. Dose-optimized TACTs are now being tested in large trials in African and Asian countries (clinicaltrials.gov identifiers NCT03923725 and NCT03939104, respectively). However, the long-term evolutionary benefits of TACT deployment cannot be assessed with clinical trials. Here, we use a consensus mathematical modelling approach to project the potential long-term evolutionary dynamics and clinical treatment outcomes of TACT deployment in different malaria epidemiological settings. The results can inform proactive policies aiming to contain the emergence and spread of antimalarial drug resistance.

## Methods

### Individual-based malaria simulation models

Two published independently built individual-based mathematical models of *Plasmodium falciparum* transmission and resistance evolution (“MORU”^20^ and “PSU”^16,21^) were used to compare the population-wide benefits of deploying TACTs versus continued ACT use. The parameterizations used here follow those in a previous consensus exercise^22^, with three key differences: (1) population size was set to one million individuals, (2) private-market drug use was included, and (3) TACTs were included as antimalarial treatment. The inclusion of TACTs required incorporation of their pharmacokinetic properties as well as pharmacodynamic parameters calibrated to treatment efficacies^23^ consistent with those found in recent clinical trials^18^ (Supplementary Figure 1). As in previous publications^21^, both models include six key drug-resistance loci: *pfcrt* K76T, *pfmdr1* N86Y, *pfmdr1* Y184F, *pfkelch13* C580Y, copy number of *pfmdr1*. These are determinants of drug resistance to chloroquine, amodiaquine, lumefantrine, artemisinins and mefloquine. *Pfplasmepsin-2,3* copy number was included also as a marker of piperaquine resistance. All parasites at model initialization carried wild-type *pfkelch13*, wild-type *pfmdr1* Y184, and single-copy *pfmdr1* and *pfplasmepsin2,3*. All four configurations of the K76T and N86Y loci (K76-N86, K76-86Y, 76T-N86, and 76T-86Y) were set to be equally represented in the parasite population, at the start of the simulation, to reflect diverse amodiaquine, lumefantrine, and chloroquine resistance profiles. Additional details can be found in Supplementary Methods.

**Figure 1.**
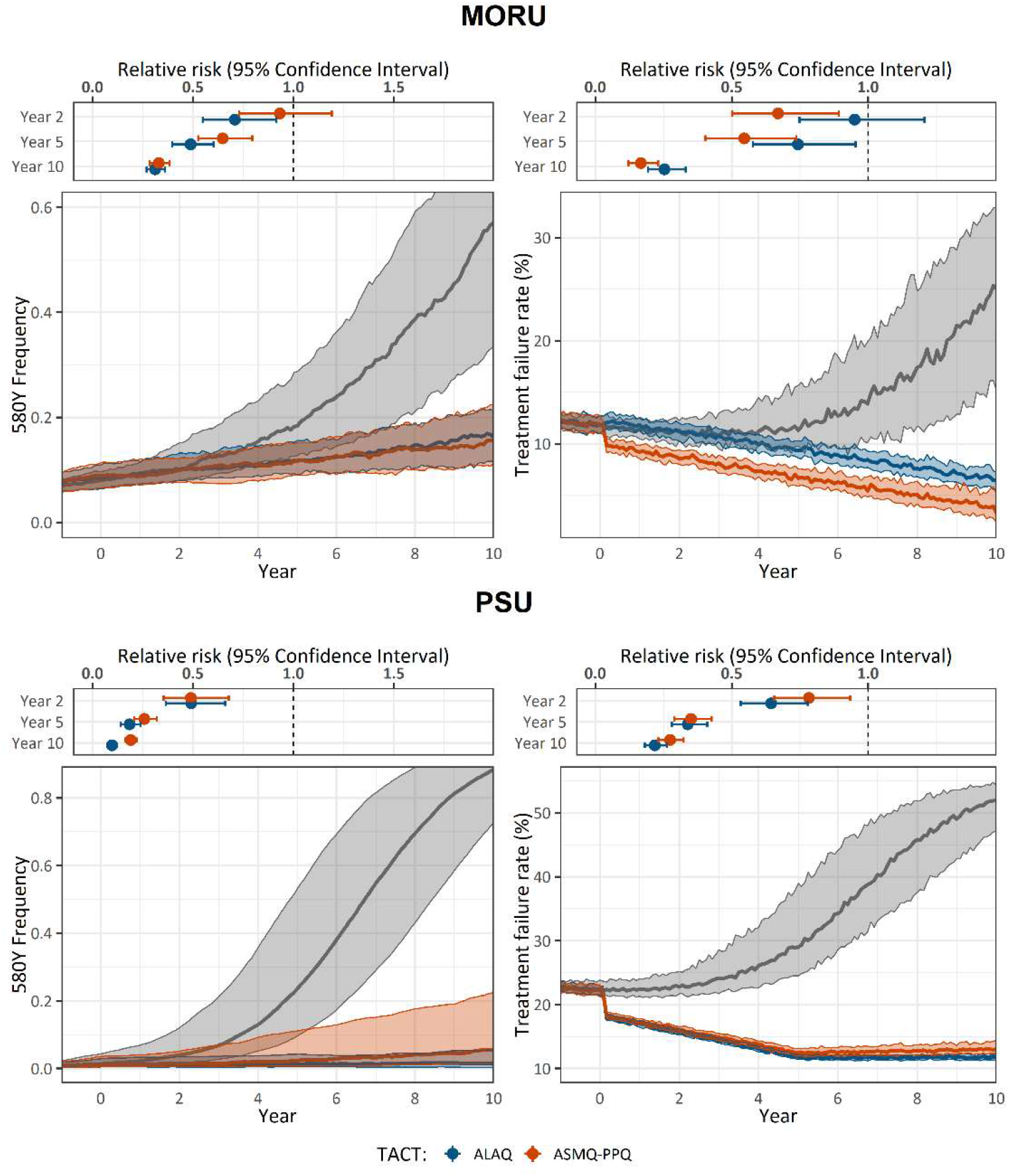
Comparison of ACT and TACT deployment at 1% PfPR and 50% treatment coverage, with DHA-PPQ used as the baseline ACT before TACTs are deployed at year zero; grey lines (medians from 100 simulations) show the evolution of the *pfkelch13* 580Y allele or treatment failure rates under continued DHA-PPQ use. Red lines show how these processes are slowed down by deployment of ASMQ-PPQ. Blue lines show how these processes are slowed down by deployment of ALAQ. All shaded areas show inter-quartile ranges. Panels above each graph show an individual’s relative risk of 580Y infection (under TACT deployment versus ACT deployment) after 2, 5, and 10 years of deployment; bars show 95% confidence intervals assuming a sample size of *n*=1000 for each deployment. Results for baseline ASAQ and AL use are shown in Figures S52 and S53.

### Scenarios evaluated

Twenty-seven historic epidemiological scenarios were evaluated, each assuming unique combinations of all-age malaria prevalence (PfPR) (0.1%, 1%, 10%), treatment coverage (25%, 50%, 75%), and baseline ACT choice (DHA-PPQ, artesunate-amodiaquine - ASAQ, AL). Throughout, we will refer to the 1% PfPR and 50% treatment coverage with DHA-PPQ scenario as the standard evaluation scenario. For each historic scenario, we explored three different prospective scenarios: (1) continued ACT use as first-line therapy (FLT); (2) switch to artesunate-mefloquine-piperaquine - ASMQ-PPQ; (3) switch to ALAQ. We analysed additional scenarios where TACTs were introduced late, introduced gradually, or with introduction of ASMQ-PPQ at a time of varying genotype frequency of resistance markers for all three components of this TACT.

### Simulation protocol

Each simulation consisted of three stages:

1. 10 burn-in years calibrated to each historic scenario during which mutation and within-host selection were not allowed (to allow the model to equilibrate at the appropriate *Pf*PR with no additional drug-resistance markers).
2. 15 years during which mutation and selection processes are active. The last year of this period marks the end of the historic scenario and is denoted as ‘year zero’.
3. 10 year long prospective scenarios simulating introduction of TACTs or continued use of ACTs.

At the start of stage 1, public-sector drug use was set to 5% of all treatments and increased linearly over ^24^ twenty years until plateauing at 80% of all treatments in year 20. We assumed that private providers offer sulphadoxine-pyrimethamine (SP), chloroquine (CQ), amodiaquine (AQ) and artemether lumefantrine (AL) in equal proportions.

### Outcome measures

Two primary model outcome measures were reported: *pfkelch13* 580Y allele frequency over time, and the treatment failure (TF) rate (the proportion of treatments that result in 28-day parasitological failure as measured by microscopy). The number of simulations of scenarios resulting in malaria elimination over a ten-year period was also recorded. Elimination in both models was defined as all-age PfPR falling below 0.01%, indicative of infections resulting mostly from imported infections.

## Results

The model predicts that replacing artemisinin-based combination therapies (ACTs) with triple artemisinin-based combination therapies (TACTs) substantially slows the emergence and evolution of artemisinin-resistant alleles and restores the clinical efficacy of first-line therapy (Figure 1). Notably at year zero, the mutant *pfkelch13* 580Y alleles were present in both models; at 0.013 (IQR: 0.006 – 0.043) allele frequency in the PSU model and 0.080 (IQR: 0.065 – 0.096) allele frequency in the MORU model. Under continued DHA-PPQ use, the allele frequency of *pfkelch13* 580Y increased to a median frequency of 0.884 (IQR: 0.725 – 0.968; PSU) or 0.571 (IQR: 0.335 – 0.769; MORU) after ten years. In both models, under continued ACT use, ten years was sufficient time for artemisinin-resistant genotypes to become established (Figure 1 and Figures S2-7). If ASMQ-PPQ was deployed, *pfkelch13* 580Y alleles were projected to reach frequencies of 0.056 (IQR: 0.014 – 0.226; PSU) or 0.155 (IQR: 0.112 – 0.224; MORU) after ten years. Under ALAQ, 580Y emergence was slower under the PSU model – predicted median allele frequency of 0.015 after ten years (IQR: 0.003 – 0.054) – but similar under the MORU model (median = 0.162; IQR: 0.116 – 0.213). The relative benefit of ALAQ over ASMQ-PPQ was sensitive to the model used and scenario examined (Figure 2). The risk of an infection carrying the *pfkelch13* 580Y allele after only 2 years of ALAQ (TACT) deployment was significantly lower than that with continued ACT use, with mean relative risks of 0.49 (95% CI: 0.36-0.66; PSU) and 0.71 (95% CI: 0.55-0.91; MORU). With ASMQ-PPQ (TACT) deployment, the relative risks were 0.49 (95% CI: 0.35 - 0.68; PSU) and 0.93 (95% CI: 0.73-1.19; MORU). The corresponding relative risks at year 10 were 0.19 (95% CI: 0.16 - 0.22; PSU) and 0.33 (95% CI: 0.29 - 0.38; MORU) for ASMQ-PPQ and 0.10 (95% CI: 0.08 - 0.12; PSU) and 0.31 (95% CI: 0.27 - 0.36; MORU) for ALAQ.

**Figure 2.**
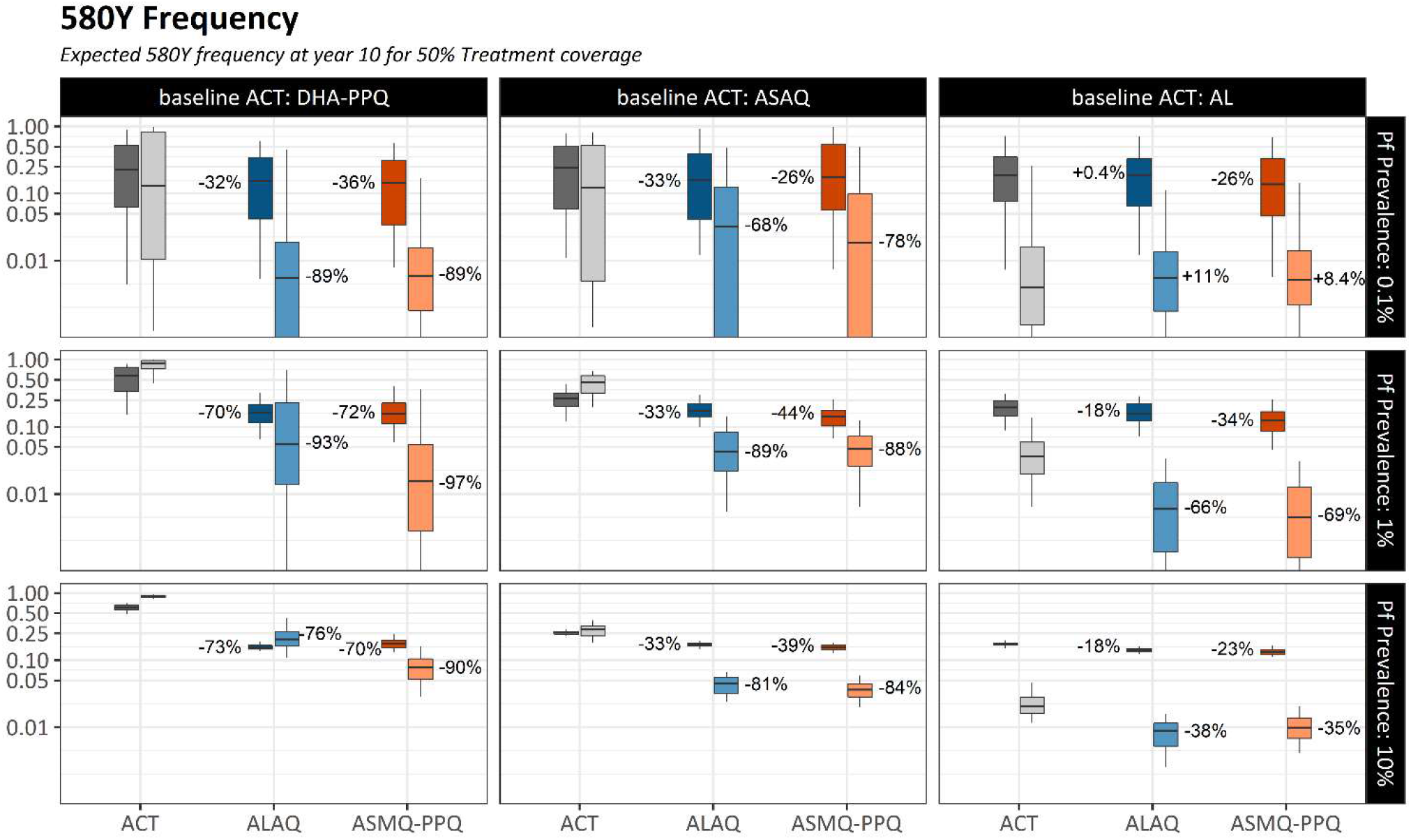
*Pfkelch13* 580Y allele frequencies after 10 years of TACT deployment or ACT deployment, in three baseline scenarios of ACT use (columns) and three prevalence settings (rows). Treatment coverage is 50%; Figures S22 and S23 show results for treatment coverages of 25% and 75%. The grey boxplots in each panel show 580Y allele frequencies (*y* axes) ten years later under a status quo ACT policy. The blue (ALAQ) and red (ASMQ-PPQ) boxplots show 580Y frequencies after 10 years of a TACT policy. Boxplot pairs have MORU model results on the left and PSU model results on the right. The percent reduction in median 580Y allele frequency from ACT to TACT deployment is shown next to the median of each TACT boxplot. All boxplots summarize 100 simulations, and whiskers show 1.5 times the IQR.

Both models agreed that the lower *pfkelch13* 580Y frequencies resulting from TACT deployment would result in lower population-wide treatment failure rates. For the standard evaluation scenario, the projected treatment failure rates (at 28 days post-treatment) after 10 years were 52.0% (IQR: 47.1% – 54.4%) for the PSU model and 20.6% (IQR: 12.6% – 29.1%) for the MORU model. After introduction of TACTs (either ASMQ-PPQ or ALAQ), the upper quartile of treatment failure rates was projected to stay below 15% (for both models and both TACTs) for the entire ten-year period. This resulted from (*i*) the increased efficacy of TACTs relative to ACTs (Figure S1) and (*ii*) the much slower emergence and spread of *pfkelch13* 580Y alleles (Figures 2; S2-7). Despite the different model predictions for the speed of spread of artemisinin-resistant alleles, both models projected that TACT introduction (at 1% prevalence and 50% treatment coverage) would lead to a >74% reduction in treatment failure rates for countries currently using baseline DHA-PPQ, >34% treatment failure rate reduction in scenarios with ASAQ as baseline, and >17% reductions when AL was the baseline treatment (Figures 3; S8-13).

**Figure 3.**
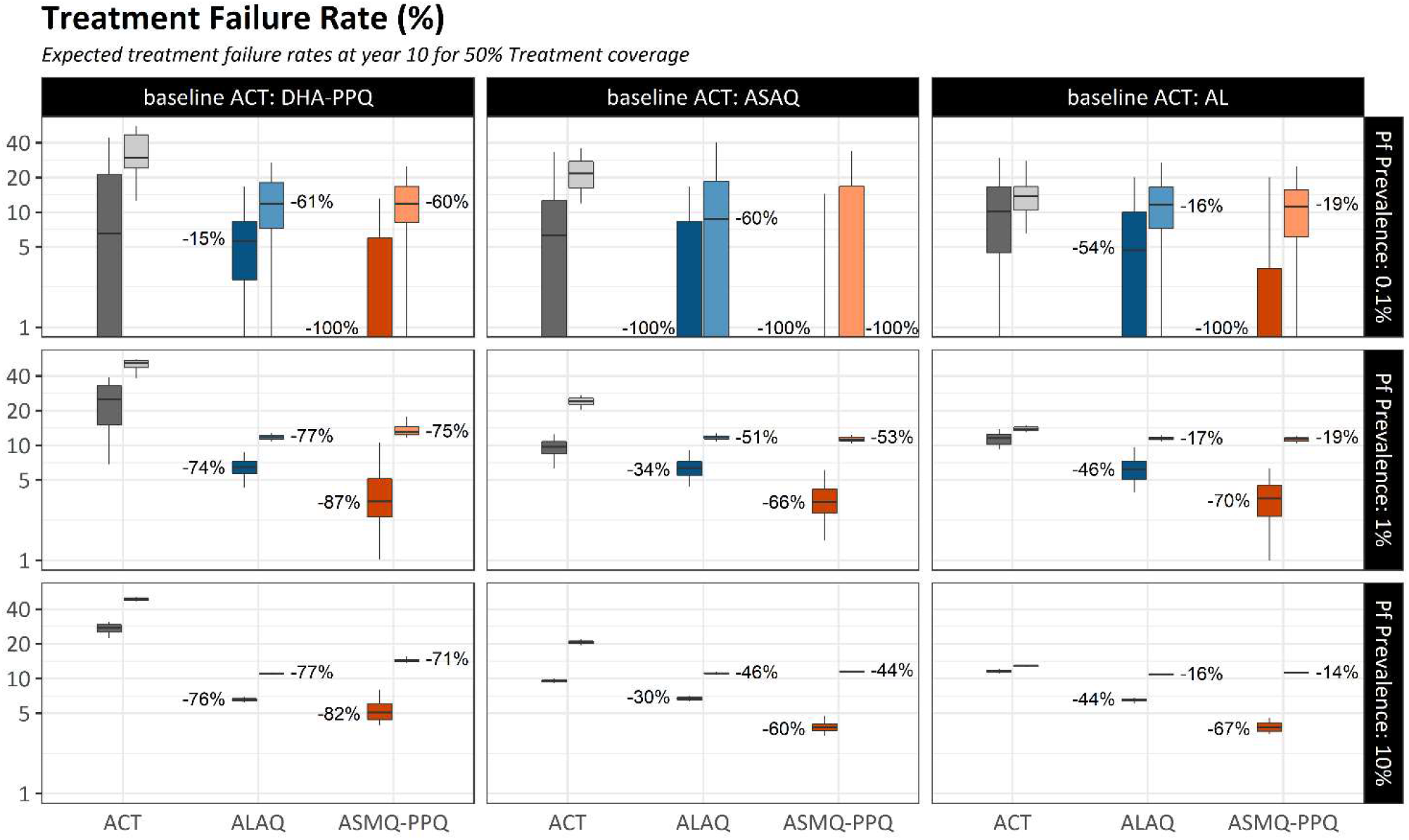
Treatment failure rates after 10 years of TACT deployment or ACT deployment, in three baseline scenarios of ACT use (columns) and three prevalence settings (rows). Treatment coverage is 50%; Figures S24 and S25 show results for treatment coverages of 25% and 75%. The grey boxplots in each panel show the TF rates (*y* axes, %) ten years later under a status quo ACT policy. The blue (ALAQ) and red (ASMQ-PPQ) boxplots show treatment failure rate outcomes after 10 years of a TACT policy. Boxplot pairs have MORU model results on the left and PSU model results on the right. The percent reduction in median TF rates from ACT to TACT is shown next to the median of each TACT boxplot. All boxplots summarize 100 simulations, and whiskers show 1.5 times the IQR..

In 52 out of 54 scenarios examined (27 per model) the 10-year treatment failure rates were lower when TACTs were introduced compared to continued ACT deployment (Figure 3; nearly all Mann-Whitney *p*-values <10^−4^; see Figures S20-21). In the other two scenarios, both in a low-transmission setting (75% treatment coverage, 0.1% prevalence, baseline AL use, PSU model; and 25% treatment coverage, 0.1% prevalence, baseline DHA-PPQ use, MORU model) the simulations projected median prevalence levels below 0.05% after ten years with median treatment failure rates of zero for both continued ACT use and switch to TACT. Thus, no scenarios showed any advantages of ACT deployment over TACT deployment – Figures S22-28.

### Preventing drug-resistance

The short-term benefits of a switch to TACTs (Figure 1) are likely to be critically dependent on the initial conditions at year 0 and the expected evolutionary trajectories of drug resistance allele frequencies. Treatment failure and allele frequency dynamics are highly nonlinear and grow exponentially in the early phases (Figures S29-34), suggesting that the timing of TACT introduction (i.e. as early as possible) could be critical to its long- and short-term success.

Following the standard scenarios (Figure 1), switching to TACTs led to median artemisinin resistance allele frequencies ranging from 0.01 to 0.17 (across the two models and two TACTs) ten years after TACT deployment. If TACT introduction were delayed by three years, the median projected 10-year *pfkelch13* 580Y allele frequency ranged from 0.08 to 0.22 and shifted to 0.24 to 0.45 if TACT introduction were delayed by 5 years. This behaviour is consistent between models across different prevalence settings although the magnitude of the effect over ten years differed between the models (Figures 4A and 4B). In very low prevalence settings delays in TACT implementation could preclude elimination (Figure 5).

**Figure 4.**
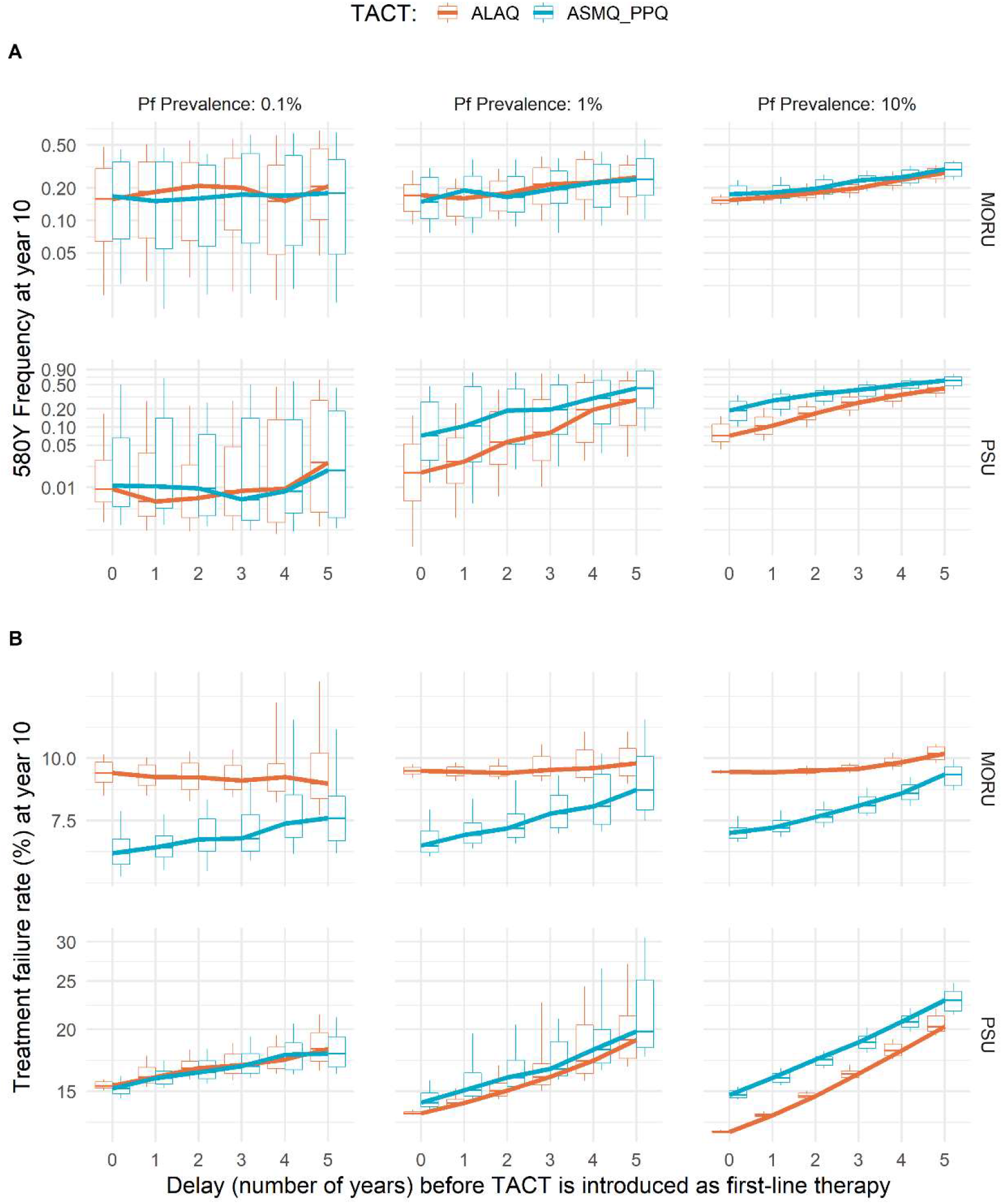
Increases in artemisinin-resistance frequency due to late adoption of TACTs. Results are shown for the MORU model (top row) and the PSU model (bottom row), for three different prevalence levels (columns), fixing treatment coverage at 50%. In each panel, the *x*-axis shows the number of years of delay before a TACT is introduced, and the *y*-axis shows the artemisinin-resistant allele’s frequency at year 10 (panel A) or treatment failure at year 10 (panel B). Box plots show interquartile ranges and whiskers show 1.5 times the IQR.

**Figure 5.**
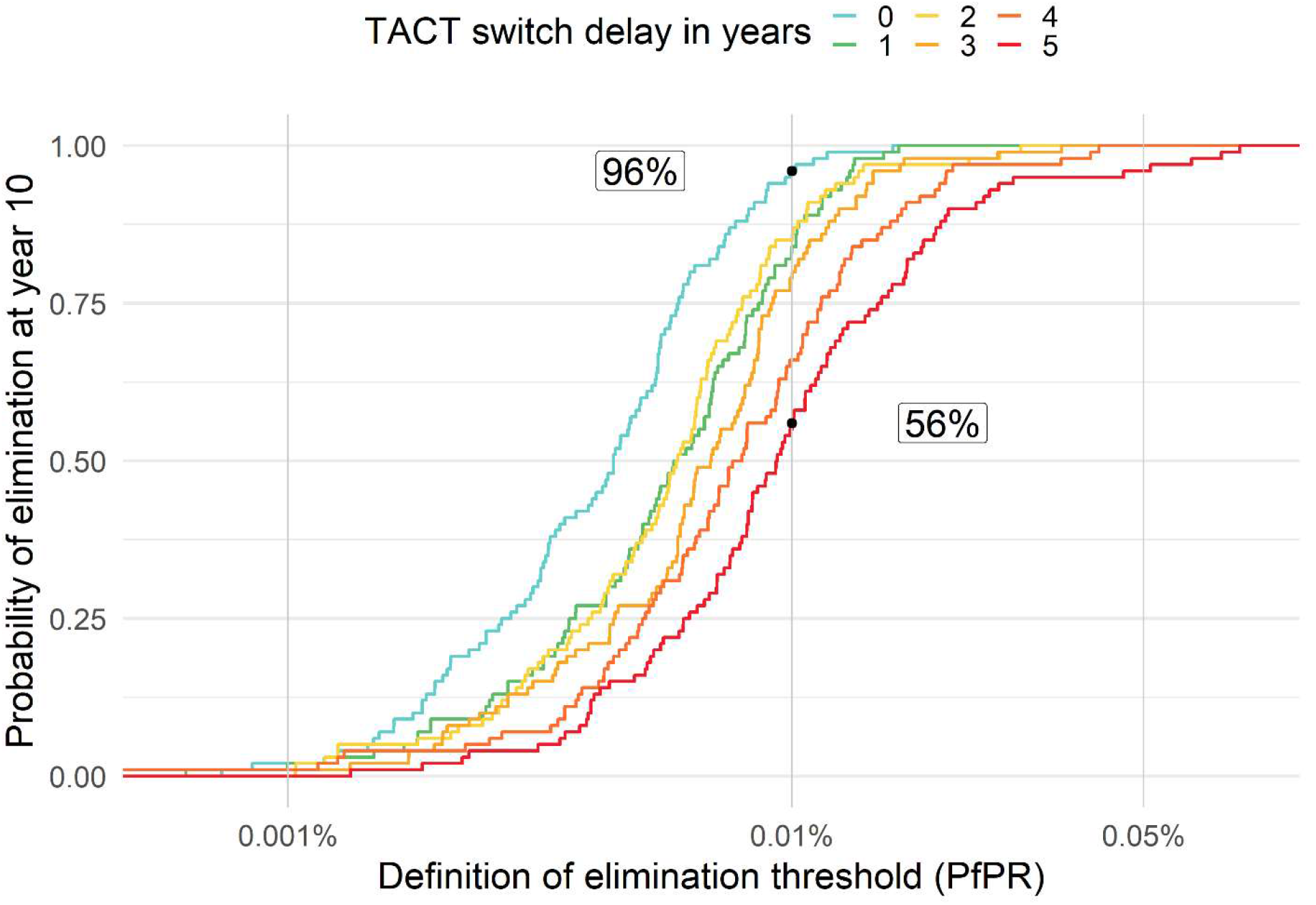
Impact of TACT implementation delay on elimination prospects. Assuming a low-transmission setting with 0.1% *PfPR* where DHA-PPQ is used as first-line therapy, we simulate a switch to ALAQ at year 0, assuming 50% treatment coverage. The probability of reaching elimination for each elimination threshold is given by the coloured lines which indicate different delays in TACT adoption. Assuming no delay in switch to TACTs, the probability of attaining a PfPR of ≤0.01% was 96%. The corresponding probability is 56% when a 5-year delay in TACT implementation is imposed.

### Pre-existing triple resistance mechanisms

The major long-term benefit of combination therapy, that multi-drug resistance emerges much later under combination therapy than single-drug resistance does under monotherapy, relies on multi-resistant genotypes being absent from the population at the time of deployment. To evaluate the risk posed by pre-existing multi-drug resistance, we explored a range of starting ASMQ-PPQ triple-resistant genotype frequencies in a PfPR setting of 0.1%. If the triple-resistant genotype’s frequency was lower than 0.01 at the time of TACT deployment, the models showed no major effect on the emergence of parasites resistant to all 3 drugs (Figure 6). However, a marked increase in the risk of triple drug-resistant mutant spread was predicted when the initial frequency of these mutants was between 0.01 to 0.05 (Figure 6A). For example, when triple mutant frequency was set to 0.04 at year 0, natural selection operated efficiently (i.e., no stochastic disappearance due to genetic drift) to produce median frequencies of 0.35 (IQR: 0.11 – 0.62; PSU) and 0.25 (IQR: 0.00– 0.55; MORU) after 10 years. This selection is enabled by the assumed 71% to 73% treatment efficacy of ASMQ-PPQ for infections with the triple-resistant genotype (PKPD Section of supplementary methods).

**Figure 6.**
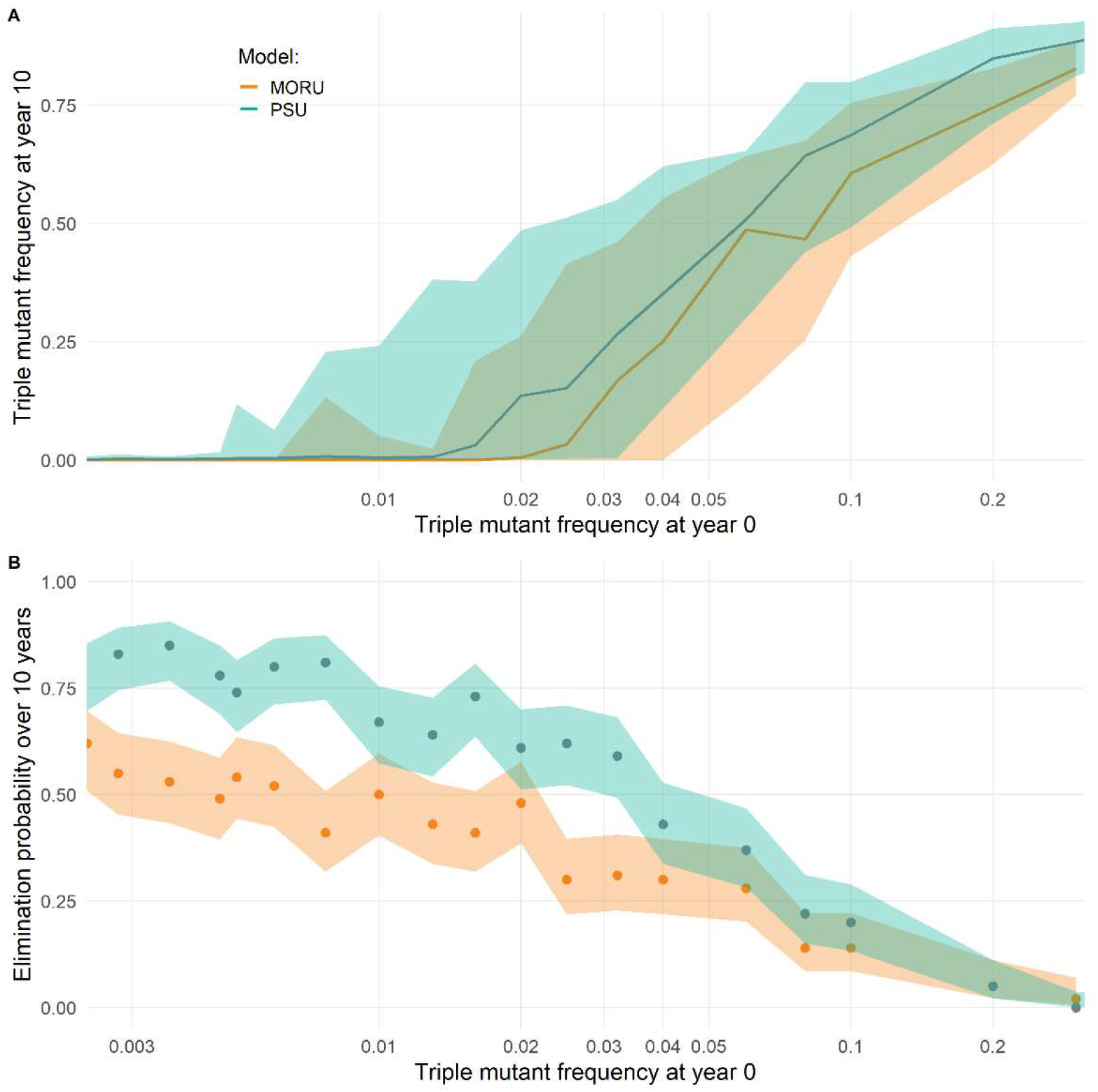
Relationship between triple mutant frequency and probability of elimination. Assuming a low-transmission setting with 0.1% PfPR where DHA-PPQ is used as first-line therapy, we simulate a switch to ASMQ-PPQ at year 0. At the time of first-line therapy switch, different frequencies of a pre-existing triple-mutant genotype conferring resistance against artemisinin, mefloquine, and piperaquine were imposed. For each initial frequency of the triple-resistant mutant (*x*-axis) 100 simulations were run. After 10 years of simulation, the triple mutant frequency was recorded, and the probability of elimination was calculated as the proportion of simulations reaching a PfPR lower than 0.01%. (A) Lines and the shaded bands show the median and interquartile range of obtained triple mutant frequencies at year 10 for both models. (B) Lines and shaded bands show the model predicted average and respective 95% confidence intervals across the 100 simulations for each value of triple-mutant frequency. Confidence intervals were generated using a one-sample proportions test without continuity correction.

Following selection of triple-resistant parasites, the probability of reaching elimination within ten years in a 0.1% prevalence setting declined substantially. For triple-resistant frequencies below 0.01 at the start of TACT deployment, 10-year elimination probabilities were approximately 80% (PSU) and 90% (MORU). However, a starting triple-resistance frequency of 0.04 led to 43% (PSU) and 30% (MORU) probabilities of elimination, with a steep drop to a <5% probability for falciparum malaria elimination with initial triple-resistant frequencies above 0.20 (Figure 6B). If prevalence is higher than 0.1%, or if treatment coverage is higher than 50%, natural selection of the triple resistant is stronger and less susceptible to the action of random genetic drift at small population sizes. Under these conditions, a triple-resistant genotype frequency >0.01 at the start of TACT deployment predictably led to rapid selection of the triple-resistant and near-term increases in treatment failure (Figures S34-36).

## Discussion

The efficacy of triple artemisinin-based combination therapy (TACT) for the treatment of drug resistant and sensitive malaria can be measured in randomized controlled trials. However, their long-term population-wide impact on prevalence, treatment failures, and drug resistance can only be predicted with mathematical models that relate malaria case management to malaria epidemiology. Consistent with expectations from evolutionary biology, the two independently validated models^16,20^ of malaria epidemiology and drug-resistance evolution evaluated here found that triple therapies (TACTs) are more effective than double therapies (ACTs) at delaying the spread of drug resistance. Safeguarding the long-term efficacy of antimalarial therapy is essential for reaching the ultimate goal of malaria elimination. The two models agreed that the main driver of loss of ACT therapeutic efficacy is the emergence and spread of artemisinin-resistance (Figures S37-39). We showed that a switch to TACTs as first-line therapy can delay the selection of *pfkelch13* 580Y mutant parasite lineages substantially, even when these are already present at low prevalence, because TACT deployment inhibits their spread when both partner-drug resistant mutations are absent. In evolution, the acquisition of mutations or additional gene copies in the same genome occurs nonlinearly in time: if it takes *y* years for a *P. falciparum* lineage to acquire a particular set of mutations it will take *y*^2^ years to acquire twice as many. An exception to this rule is predicted only under very high rates of recombination ^25,26^.

Comparing the two TACT options evaluated, the persistence of artemisinin-resistant alleles is predicted to be disrupted most, and thus resistance delayed for longer, by deployment of ALAQ. This is because of the inverse relationship between resistance to amodiaquine and lumefantrine^27,28^Our results suggest that in unusual low to moderate drug selection pressure settings, i.e., low treatment coverage (≤50%) and low *Pf* prevalence (≤1%), the 10-year benefits of ALAQ compared to ASMQ-PPQ are not substantially different. This results from (*i*) the higher efficacy of PPQ as compared to the other partner drugs (Figure S1) and (*ii*) the higher efficacy of ASMQ-PPQ on its single-resistant and double-resistant mutants. In contrast, in areas of higher transmission intensity with good treatment coverage (≥75%), ALAQ is superior in preventing the spread of artemisinin resistance and conserving therapeutic efficacy (Figures S29-31) Notably all TACT policies evaluated here are non-adaptive, i.e., they do not use concurrent surveillance information to adjust treatment guidelines. An adaptive TACT strategy, based on real time resistance data, would lead to an even more favourable TACT assessment than the one discussed here.

It is generally agreed that investing in preventing the emergence of resistance to any anti-infective therapy is preferable to reacting to an epidemic of drug resistance^29,30^. It is cost-beneficial, and it results in less morbidity and mortality. Where resistance arises very rapidly within an individual, such as in tuberculosis or HIV infection, triple or quadruple therapies were soon adopted as the standard of care since the value of resistance prevention to individual patient outcomes was obvious. Antimalarial drug resistance is more insidious. Over the past seventy years, antimalarial drug resistance has killed millions, mainly African children^31^. Yet, public health response has generally been slow^32^, with treatment policies only changing once resistance had become firmly established and the antimalarial drugs were either failing or completely ineffective. If TACTs were adopted now in Africa, where artemisinin resistance is still mostly absent, they could have a large impact on the course of artemisinin-resistance evolution, and thereby avoid preventable malaria morbidity and mortality. In areas such as Rwanda and Uganda, where several *pfkelch13* mutants have increased to >0.20 allele frequency regionally, TACTs could have a clear and immediate impact^8,9^. To quantify how the benefits of delaying artemisinin and partner drug resistance can be compromised by decision making inertia, we evaluated scenarios where TACTs were introduced with delays of one to five years. The two independent models agree that delays in TACT deployment will result in higher long-term *pfkelch13* 580Y frequency and more treatment failures (Figure 4), increasing morbidity and compromising the likelihood of elimination in low transmission settings (Figure 5). Immediate but gradual TACT adoption will have a similar impact on *pfkelch13* 580Y frequency and treatment failure rates (Figure S54).

Use of individual antimalarial drugs at low doses and/or with poor adherence, or with low quality medicines, enables the emergence of resistance. In Southeast Asia, *P. falciparum* has acquired resistance mechanisms to all the antimalarial drugs that have been deployed, including mefloquine and piperaquine. We therefore investigated to what extent triple-resistant mutants undermine malaria elimination efforts and resistance containment strategies. The potential for selection of a multi-drug resistant genotype is the Achilles’ heel of any combination therapy. Our models predicted that even a 0.01 multi-drug resistance genotype frequency affecting all three components of the ASMQ-PPQTACT results in efficient and predictable natural selection of this triple resistant mutant. Above this frequency threshold, this multi-drug resistant genotype will no longer be susceptible to stochastic loss through bottlenecking or random genetic drift, and natural selection will act efficiently to bring the multi-resistant type up to high frequencies. Molecular surveillance is thus particularly important for this TACT.

### Limitations

Our two independent models make different assumptions on mechanisms of immunity, symptoms presentation (Supplementary Table 1), and within-host evolution. This is important for TACT evaluation because the selection pressure exerted by TACTs will differ by genotype, but the impact of changes in fitness conferred by each genotype in any drug-resistance model is dependent on treatment coverage^33^ and symptoms presentation^17^. If these have been parameterized incorrectly, the true patterns of selection may be different than modelled here. To address this issue, we calibrated model parameters to force the two models to reach 0.01 artemisinin-resistant allele frequency at the same time under a specified set of conditions. However, selection pressures across models do differ for the artemisinin resistant 580Y allele frequencies above 0.01. Together with the dynamic use of four or more different drugs over time, this renders calibrating both models to the exact same epidemiological scenario impossible. This results in slightly different initial starting allelic frequencies (particularly *pfkelch13* 580Y), as well as diverging treatment failure rates, at the start of each prospective scenario. Reassuringly, the benefits of TACT over ACT deployment are robust to these model differences (Figures 1-3).

There is limited information on the direction of resistance selection under ALAQ treatment as AQ selects for some alleles (76T, 86Y, Y184) while lumefantrine selects for their counterparts (K76, N86, 184F). The strengths of these selective pressures are difficult to estimate^23^ particularly as relative strengths of intra-host selection change over time because lumefantrine is eliminated from the blood more rapidly than amodiaquine. We assumed that triple-resistant genotypes to ALAQ cannot emerge, as collateral sensitivity at three separate loci ensures that evolution towards lumefantrine resistance increases amodiaquine sensitivity^28^. However, this could be compromised by novel lumefantrine resistance mechanisms which are not conferred by *pfmdr1* and *pfcrt*^34^. Triple resistance to ASMQ-PPQ, however, can and does emerge in the model simulations, by successive acquisition of the *pfkelch13* 580Y allele (or a similar *pfkelch13* allele conferring delayed parasite clearance), additional *pfmdr1* gene copies, and an additional copy of the *pfplasmepsin-2,3* genes. In addition, spread of artemisinin resistance could also be driven by additional factors, such as increased transmissibility of *pfkelch13* mutant infections. In this scenario, the resistance delaying effect of TACTs on artemisinin resistance spread would be less than predicted by our model, but the mutually protective effect between ACT partner drugs would remain the same.

For specific in-country evaluations, the geography, variation in prevalence, monthly case burden, and drug access should all be locally parameterized to provide detailed model forecasts of the potential benefits of TACT deployment.

### Conclusions

The two independent models agreed that deployment of TACTs will delay the emergence and spread of artemisinin and partner drug resistance significantly. This would secure effective antimalarial treatment for the next 5 to 10 years, a prerequisite for successful malaria elimination and control. Critically, the results advocate for an immediate deployment of TACTs, as each year TACT deployment is delayed translates into diminishing long-term benefits. This emphasises the importance of preventing antimalarial resistance rather than allowing resistance to establish and then trying to reverse it.

## Research in context

### Evidence before this study

The emergence and spread of antimalarial resistant parasites have threatened malaria control in the Greater Mekong Subregion. The presence of both artemisinin and partner drug resistant parasites has resulted in high treatment failure rates of artesunate-mefloquine on the Thai-Myanmar border and of dihydroartemisinin-piperaquine in Cambodia, Thailand and Vietnam. A multi-site clinical study has demonstrated that triple artemisinin combination therapies (TACTs) are highly efficacious even in contexts where ACT failure rates were extremely high.

### Added value of this study

While it is reassuring that antimalarial therapeutic efficacy can be restored in areas of widespread drug resistance, field studies cannot elucidate the long-term benefit of deploying TACTs in countries with little to inexistent artemisinin-resistant parasites (i.e. African countries). The question of whether immediate introduction of TACTs can delay the spread of artemisinin resistance and limit the increase in treatment failure rates and morbidity can only be addressed by mathematical models.

Using two independent modelling approaches, the current work describes the projected epidemiological impact (over a 10-year period) of TACT deployment. This entails comprehensive comparisons of epidemiological outcomes obtained with a switch to TACTs as first line therapy versus continued ACT usage. These comparisons are performed for a wide range of scenarios with respect to the drugs used, malaria prevalence, treatment coverage, delay in introduction of TACTs, and the effect of pre-existing multiple resistance.

### Implications of all the available evidence

The use of TACTs could significantly delay the emergence and spread of artemisinin and partner drug resistance thus improving the chances of success of malaria control and elimination efforts. The potential benefits of TACT progressively decrease with each year of delay in their deployment, implying that the switch to TACTs must be made well before multidrug resistance is established.

## Data Availability

All data produced are available online at https://github.com/ATOME-MORU/deTACT

## Author contribution

AD, CA, MD, MB, and RA conceptualised the study. TDN, BG, MB, and RA set-up the simulation protocols. TDN and BG performed the necessary model calibrations, ran the simulations, and compiled the model outputs. TDN, BG, MB, and RA conducted the subsequent analyses. MB and RA wrote the initial draft. All authors reviewed the manuscript draft. All authors read and approved the final manuscript.

## Acknowledgements

This research falls under the auspices of the Development of Triple Artemisinin-based Combination Therapies (DeTACT) project funded by UK Aid and the UK Government’s Foreign, Commonwealth and Development Office. Funding from Wellcome Trust (220211) is also acknowledged.

MFB, TDN, TN-AT acknowledge funding from the National Institutes of Health grant NIAID R01AI153355 and The Bill and Melinda Gates Foundation INV-005517 grant awarded to Pennsylvania State University.

RA acknowledges funding from the Bill and Melinda Gates Foundation (OPP1193472).

