## Supplementary Materials for "Preventing antimalarial drug resistance with triple artemisinin-based combination therapies"

**Supplementary Material** for "Preventing antimalarial drug resistance with triple artemisinin-based combination therapies"

### Model Description

Two individual-based microsimulations were used to compare patterns of artemisinin-resistance evolution in different contexts of prevalence, treatment coverage, and pre-existing partner-drug resistance. The two models are called (1) the 'MORU Model', developed at the Mahidol-Oxford Research Unit (MORU), Nuffield Department of Medicine, University of Oxford; and (2) the 'PSU Model', developed at the Center for Infectious Disease Dynamics (CIDD), Department of Biology, Pennsylvania State University. The models all run as daily time-step discrete-event simulations of individuals (humans) who can be infected with *Plasmodium falciparum* malaria and subsequently pass on their infection to other individuals in the simulation via mosquitoes which are also explicitly modelled.

*Locus structure.* The current state of the science in malaria individual-based modelling allows for only a limited number of loci to be included in an evolutionary analysis of drug resistance, as (1) there is no general genotype-phenotype relationship among all the known falciparum drug-resistant genotypes and their clinical/parasitological phenotypes and (2) there are computational limitations on the required size of recombination tables (necessary for modelling the sexual stage of falciparum reproduction) which are of dimension $2^n \times 2^n \times 2^n$ when $n$ loci are included. Following previous work(1), both models include seven key drug-resistance loci: *pfcrt* K76T, *pfmdr1* N86Y, *pfmdr1* Y184F, *PfKelch13* C580Y, copy number of *pfmdr1* – which are major determinants of drug resistance to artemisinin, lumefantrine, amodiaquine, piperaquine, mefloquine, and chloroquine – as well as *plasmepsin-2,3* copy number – associated with piperaquine resistance. All parasites at model initialization carried wild-type *PfKelch13*, wild-type *pfmdr1* Y184, and single-copy variants for both *pfmdr1* and the *plasmepsin* genes. All configurations of the K76T and N86Y loci (K76-N86, K76-86Y, 76T-N86, and 76T-86Y) were set to be equally represented in the parasite population to reflect diverse amodiaquine, lumefantrine, and chloroquine resistance landscapes. Several key *pfcrt* mutations that are known to be associated with piperaquine resistance(2) are not included in the model, and we assume a worst-case scenario in this analysis, i.e. that these background mutations are present bringing DHA-PPQ efficacy down to 41.5% on its double-resistant genotype(3, 4) (580Y, double-copy plasmepsin). The 580 locus in *PfKelch13* can be viewed as a proxy for the evolution of 561H in Rwanda(5) as mutations in this locus have been associated with delayed parasite clearance(6). We have used *pfplasmepsin2-3* amplification as the marker of piperaquine resistance, although the primary causal mutations are in *pfcrt* downstream from the chloroquine resistance locus(4, 7).

*Mutation and importation*. Mutations and copy number increases/decreases can occur at all loci, and as in most modelling analyses in evolutionary epidemiology, the mutation process is modelled as the conversion of an entire within-host parasite population from one genotype to another. In other words, this process models the combined dynamics of mutation and within-host fixation of a new genotype. In the PSU model, this can only occur during a period of drug treatment and only from a less resistant genotype to a more resistant genotype (defined by higher treatment failure of the current therapy on that genotype). In the MORU model this same process is complemented by reverse mutations, where a very resistant genotype can mutate to a lesser resistant genotype in the absence of drug pressure. As this mutation-plus-fixation process is difficult to parameterize from field data, a previous calibration was used where both models' mutation processes are aligned to achieve 0.01 580Y allele frequency after 7.0 years of DHA-PPQ use at 40% coverage and 10% PfPR; this ensures that one model does not produce a larger number of *de novo* mutants than the other. Artemisinin-resistant 580Y mutants are also imported once every 100 days via a Poisson process.

*Drugs used*. A proportion of infectious mosquito bites in the model result in successful infection and symptoms (depending on a person's malaria history and current level of immunity) causing an individual to seek and receive antimalarial treatment with some probability. This probability (the 'treatment coverage') is set to 25%, 50%, and 75% in different sets of simulations, allowing us to examine scenarios with poor access to drugs or variation in individual choices to seek treatment or not. In our scenarios, the baseline ACTs available in the public sector – i.e., the recommended first-line therapies prior to TACT introduction – are dihydroartemisinin-piperaquine (DHA-PPQ), artesunate-amodiaquine (ASAQ), and artemether-lumefantrine (AL). The four therapies available in the public sector are chloroquine, amodiaquine, sulphadoxine-pyrimethamine, and AL; sulphadoxine-pyrimethamine (SP) is assumed to have a fixed efficacy of 40%. Two triple ACTs that will be available for deployment in the model are artesunate-mefloquine-piperaquine (ASMQ-PPQ) and artemether-lumefantrine-amodiaquine (ALAQ).

*PKPD*

As mentioned above, 64 genotypes are modelled reflecting all combinations of two possible alleles at each of six loci. This meant we had to generate a list of and $6 \times 64 = 384$ EC50 values, representing the pharmacodynamic (PD) properties of a particular drug compound on each parasite genotype (see Supplementary Materials 2 in Nguyen et al(8)). We assumed drugs have independent parasite-killing activity and pharmacokinetic (PK) dynamics, i.e., each drug's EC50 values and half-life are unaffected by the presence of other drugs. Enrolling 10,000 simulated patients in a therapeutic efficacy study, with initial parasitaemia ranging from 2000 to 200,000 parasites/µl (uniform distribution on the $\log_{10}$ scale of 3.3 to 5.3), gives a 28-day efficacy of 99.89% for ASMQ-PPQ and 99.44% for ALAQ, consistent with field data(9) - Supplementary Figure 1.

*Metrics of evolutionary dynamics*

In malaria infections, as in all of evolutionary biology, the amount of time it takes a drug-resistant allele to establish and reach fixation depends on the size of its fitness advantage. In our context this is defined by a resistant genotype's ability to recrudesce despite a complete course of ACT or TACT. This is because recrudescence is necessary for resistant parasites to reach transmissible densities in the primary selection event, and thereafter recrudescence is an important driver of spread. These recrudescence rates – or treatment failure rates – differ by genotype and treatment as mentioned in the PKPD section above and determine whether a particular therapy has an advantage over another in slowing or controlling drug-resistance. Throughout the paper we refer to treatment failure rates as the percentage of patients who present with a parasitaemia greater than 10 parasites/µl on day 28 following treatment.

In each simulation we track relevant epidemiological, clinical, and genetic indicators that may provide insights into the evolutionary dynamics at play for each of the explored settings. Alongside treatment failure, the main metrics reported throughout are *Pf* prevalence (*PfPR*) – measured as the proportion of all individuals in the population with a parasitaemia over the limit of microscopy detection – and allelic frequencies for the tracked loci – weighted number of parasite-positive individuals carrying genotype X divided by the total number of parasite-positive individuals (the weight for each person describes the fraction of their clonal populations carrying genotype X; e.g. an individual hosting five clonal infections, of which two are caused by genotype X, would be given a weight of 2/5.

*Statistical analyses*

For each scenario explored, we ran 100 model instances. We consistently summarize the distributions of outcomes of interest using the median and interquartile ranges. Where relevant, we provide the 5$^{th}$ and 95$^{th}$ percentiles of said distributions. We mostly use median results to extrapolate relevant differences across scenarios as in Figures 2-4. We also provide more rigorous statistical metrics to infer the significance of the observed differences when comparing continued ACT use with introduction of TACTs (ASMQ-PPQ or ALAQ). Figures S20 and S21 show the Mann-Whitney p-value for each of the 108 comparisons of the two main outcomes: 580Y frequency and treatment failure rate. These p-values overwhelmingly support rejection of the null hypothesis – that the distributions of each of the outcomes is identical. As discussed at length in the main text, a switch to TACTs is generally expected to yield a lower frequency of 580Y as well as lower treatment failure rates. A second statistical evaluation of the same comparators involved the estimation of the independent relative risks of acquiring an infection with a parasite carrying a 580Y allele and failing treatment. For each scenario, we took distribution ω composed of the simulated 580Y frequencies at year 10 for each of the 100 runs as well as distribution θ from the equivalent treatment failure rates. Taking the population level 580Y frequency at year 10 as the probability that any infection with a malaria parasite will include the 580Y allele, we can then randomly sample X values from ω and compared them with a uniformally distributed random number (between 0 and 1) to get a synthetic population of X infected individuals, a subset of which carry the 580Y allele. This allows us to extrapolate a measure of relative risk by comparing the subsets carrying the 580Y allele generated from distributions ω obtained from continue ACT use (non-exposed or control group); against the equivalent subsets obtained from the TACT switch scenarios (exposed or intervention group). For example, for a treatment coverage of 50% and Pf prevalence of 1%, we aggregate the final 580Y frequency for all 100 runs with both continued ACT use and switch to TACT and:

1- Sample 1,000 values $F$ of 580Y frequency (with replacement) from the distribution of outcomes resulting from 100 model simulations for each scenario.
2- Sample 1,000 values $U$ from a uniform distribution between 0 and 1 and compare them against the 1,000 values obtained in step 1. If $U_i < F_i$ (where $i$ indicates each sample), then Pi = 1, otherwise Pi = 0. P is the set of synthetic infected individuals with cardinality 1,000 and a sum equal to the number of individuals expected to have been infected with a 580Y allele.
3- After obtaining a set P for the continued ACT use scenario ($P_{act}$) and an equivalent set for a switch to TACT scenario ($P_{tact}$), we can calculate the relative risk of having an infection with a 580Y allele in the ACT scenario vs. the TACT scenario - Probability of infection with 580Y in the ACT scenario / Probability of infection with 580Y in the TACT scenario - simply by using the sums of the corresponding sets, knowing both sets are composed of 1,000 samples.

The same calculations were performed for the treatment failure rate outcome and these results are illustrated through forest plots in Figure 1.

**Supplementary Table 1.** Comparative model features

| | **PSU Model** | **MORU Model** |
|---|---|---|
| **Key representative publications** | Multiple first line therapies study in Lancet Global Health (Nguyen et al. 2015) and therapeutic efficacy by genotype study bioRxiv preprint (Nguyen et al. 2021)) | Mass drug administration study in eLife (Gao et al. 2020) |
| **Accessibility (Either Github or description of code availability etc.)** | Code is open source at https://github.com/bonilab/malariaibm-MMC-WP2-partnerdrugresistance (version 3.2) | Code is open source at https://github.com/ATOME-MORU/malaria-model-v1.0-20.3.19. |
| **Seasonality** | Yes; not used in present analyses. | Yes; not used in present analyses. |
| **Heterogeneity in exposure** | Yes | Yes |
| **Blood-stage parasite densities modelled** | Yes | No |
| **Parameterization for clinical incidence** | Calibrated to data sets assembled for five different African studies measuring age-specific clinical incidence and EIR. No formal model fitting done for parameterization in current analysis. | We parameterized the age dependent risk of infection during the first 10 years of life using data from 8 endemic countries in sub-Saharan Africa (Aguas et al., 2008). Given the age-dependent force of infection function, we fit the model-predicted age-dependent clinical disease incidence against two separate data sets from SE Asia (one with age, one without). Details in ((Gao et al. 2020). |
| **Parameterization for severe disease and mortality incidence** | Mortality only. No tracking of severe malaria. Treatment failures and untreated cases are associated with 4% mortality (age groups 0-1), 2% mortality (age groups 2-5), 0.4% mortality (age groups 6-10), and 0.1% mortality (age groups 11 and older); see section 6 of supplement to (Nguyen et al. 2015). Mortality is zero for successfully treated malaria cases. | Of those clinical infections, we estimate the proportion that results in severe disease and hospitalization (by age) from data in (Marsh and Snow 1999). The (Marsh and Snow 1999) dataset also informs the mortality rates per hospitalized case per age group. Maximum mortality rates in hospitalized children was 5% (<2 year olds). Older children had a greater treatment success rate with mortality rates of less than 0.5%. |
| **Drug interventions – PK- PD Drug action etc** | Single compartment PK model used, with daily parasite killing, as a function of drug concentrations and parasite genotypes, for PD model. | Parasite clearance modelled using PD data from parasite clearance studies, with daily parasite killing, as a function of initial drug concentration, drug IC50 and parasite genotype. |
| **Genotype tracking and sensitivity to drugs** | Yes, four key resistance loci and two copy number variants tracked. 64 total genotypes. | Yes, four key resistance loci and two copy number variants tracked. 64 total genotypes. |
| **Vector control Interventions** | Indirect vector control only, via transmission parameter that determines the daily amount of biting. | Yes – LLIN, IRS, Larval Control |

| | | |
|---|---|---|
| **Treatment interventions** | Yes, many types of drug policies such as multiple first-line therapies, cycling, adaptive cycling, triple therapy, mass drug administration, and private-market drug sales. | Yes, treatment of clinical disease, MDA, MSAT, adjunctive primaquine, TME, iPT, and private market-drug sales. |
| **Treatment seeking and drug coverage** | Explored in the sensitivity analysis, at 25%, 50%, and 75% coverage. | Explored in the sensitivity analysis, at 25%, 50%, and 75% coverage. |
| **Spatial dynamic model** | Present analyses run in a single location. | Present analyses run in a single location. |
| **Super-infections, co-infections, multiplicity of infection** | Yes, tracked explicitly as coinfections arising from additional bites on already infected individuals. | Yes, each human individual can carry up to 10 different parasite populations (one per inoculum). Each inoculum is considered to be a clonal population upon emergence from the liver, with one parasite acquired from each infectious bite. |
| **Heterogeneity in Exposure** | Exposure varies both by age and between individuals. | Exposure varies both by age and between individuals. |
| **Duration of infection** | Not drawn from a predetermined distribution. The duration of infection is determined by explicit modelling of parasitaemia and how drugs and the immune system act on parasites. Calibrated to malariatherapy data. Durations of infection in the model range from 60 to 281 days. | Asymptomatic infection duration based on malariatherapy data and clinical follow-up data from endemic areas, as well as drug efficacy data (PD). Also estimated from a set of 8 Endemic areas from Sub-Saharan Africa (Aguas et al 2008). |
| **Clinical disease and history of exposure** | A proportion of infected individuals go on to develop clinical disease. Immunity to clinical disease develops with exposure and age. Simulation also has a maternally acquired component. | Proportion developing clinical disease depending on cumulative immunity from prior exposures and immunity level (indicator of recent exposure). |
| **Decay of natural immunity** | Exponential decay of naturally acquired immunity. | Exponential decay of naturally acquired immunity as estimated in (Aguas et al. 2008) |
| **Infectiousness and gametocyte models** | Human infectiousness to mosquitos is a function of asexual parasite density, with a time lag built in to model the fact that infectious gametocytemia lags asexual parasitaemia. | Infectiousness depends on lagged development of sexual stage parasites and seasonal transmission equation from fitting to incidence data. It is informed by parasite density in an indirect way: clinical individuals are assumed to have a higher mean infectiousness compared to asymptomatic individuals as they carry higher parasite density loads. |
| **Entomological models** | 11-day lag built in so that FOI today depends on the biting done by mosquitoes 11 days ago. No other entomological built-in features. | Full IBM component to mosquito dynamics. For this exercise we use a simplified version where only infectious mosquitoes are tracked individually. |
| **Recombination model** | Recombination can occur in a mosquito bite on a multi-clonal host. Parasites are taken up by the mosquito in proportion to their parasite density. A full recombination table is built for all possible forces of infection resulting from this host's contribution to the next generation of infectious sporozoites, according to the normal rules of Mendelian genetics. No interrupted feeding allowed in present analyses. | No recombination. No interrupted feeding. |

| | | |
|---|---|---|
| **Mutation model** | No back mutation. Mutation can occur during treatment only when the mutation confers a resistance benefit to the current treatment. | No back mutation. Mutation can occur during treatment (higher rate), and in the absence of treatment (lower rate). The higher rate was calibrated during the calibration exercise (Watson et al, 2022). |
| **Stochasticity** | All model components are stochastic and described by defined probability distributions defined in (Nguyen et al. 2015) and (Nguyen et al. 2021). The only non-stochastic elements are the delay from mosquito oocyst formation to rupturing, which is modelled as a fixed duration. | All model components are stochastic and described by defined probability distributions defined in (Gao et al. 2020). |
| **PfPR range models calibrated against.** | Immunity-symptom relationship calibrated to data sets where PfPR > 5%, across an EIR range of 10 to 200 (Nguyen et al. 2015). | Calibrated based on 8 data sets with a $PfPR_{2-10}$ minimum of 2% (Aguas et al. 2008). |

#### Supplementary Figures

**Figure S1.** Simulated therapeutic efficacy study using the Penn State model's single-compartment PK/PD model and the EC50 parameterizations from (10). A total of 20,000 patients with baseline parasitaemia of 2000 parasites/µl to 200,000 parasites/µl (distributed uniformly on the $\log_{10}$-scale of 3.3 to 5.3) were given a standard 3-day regimen of each drug (*n*=10,000 patients), without any loss to follow-up. The density plots show the asexual parasitaemia distribution among the 10,000 patients each day following treatment. Three solid orange lines mark the 99$^{th}$, 98$^{th}$, and 97$^{th}$ percentiles of those distributions, and the two dashed orange lines show the 95$^{th}$ and 90$^{th}$ percentiles. Orange percentile markers are not visible for all days as they may overlay each other at 0.00002 parasites/µl. The therapeutic efficacy for each drug is given as a percentage above the last day's density plot and illustrates the percentage of patients with a parasitaemia lower than 10 parasites/µl on day 28 following treatment.

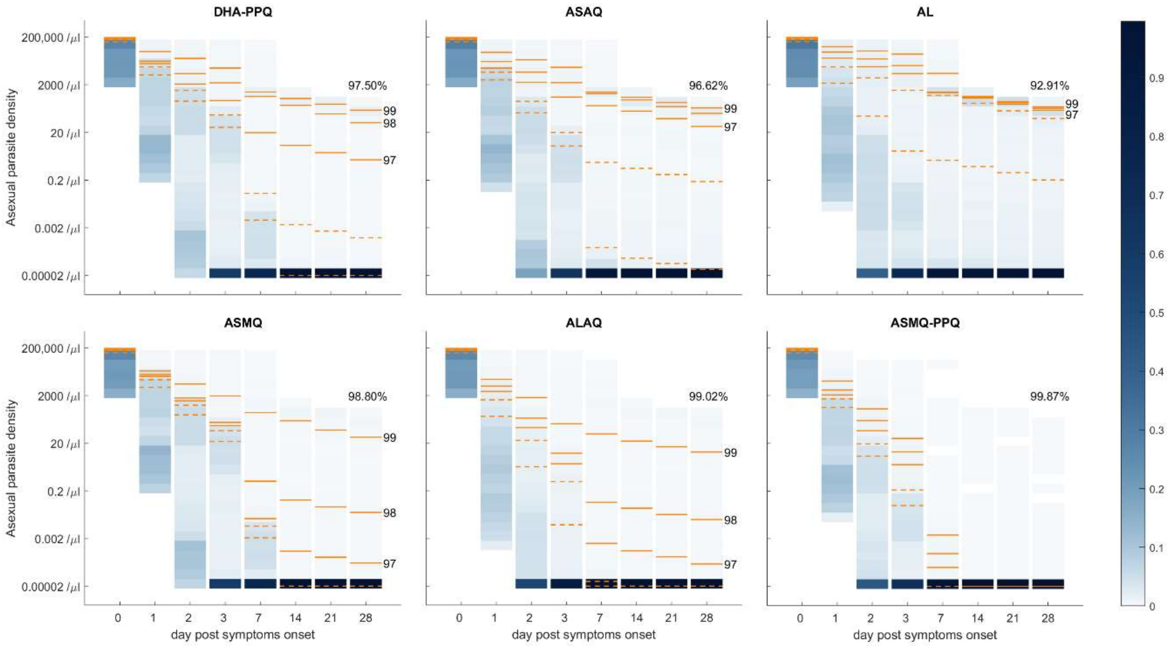

**Figure S2. 580Y allele frequency over time.** Panels show the evolution of 580Y allele frequency in a population of 1,000,000 individuals, in the years following a switch of first line therapy to TACTs. Each panel corresponds to a combination of transmission intensity: 0.1% PfPR (top row), 1% PfPR (middle row), 10% PfPR (bottom row) and baseline ACTs used before TACTs are deployed at year zero: DHA-PPQ (left column), ASAQ (middle column), AL (right column). Median trajectories are shown from 100 simulations, and the shaded area shows the range from the 5th to the 95th percentile. Grey lines show the evolution of the 580Y allele frequency under continued baseline ACT use. Red lines and blue lines show the evolution of the 580Y allele frequency after the deployment of ASMQ-PPQ and ALAQ, respectively. Figure title shows model used, outcome tracked, and treatment coverage (TC) level.

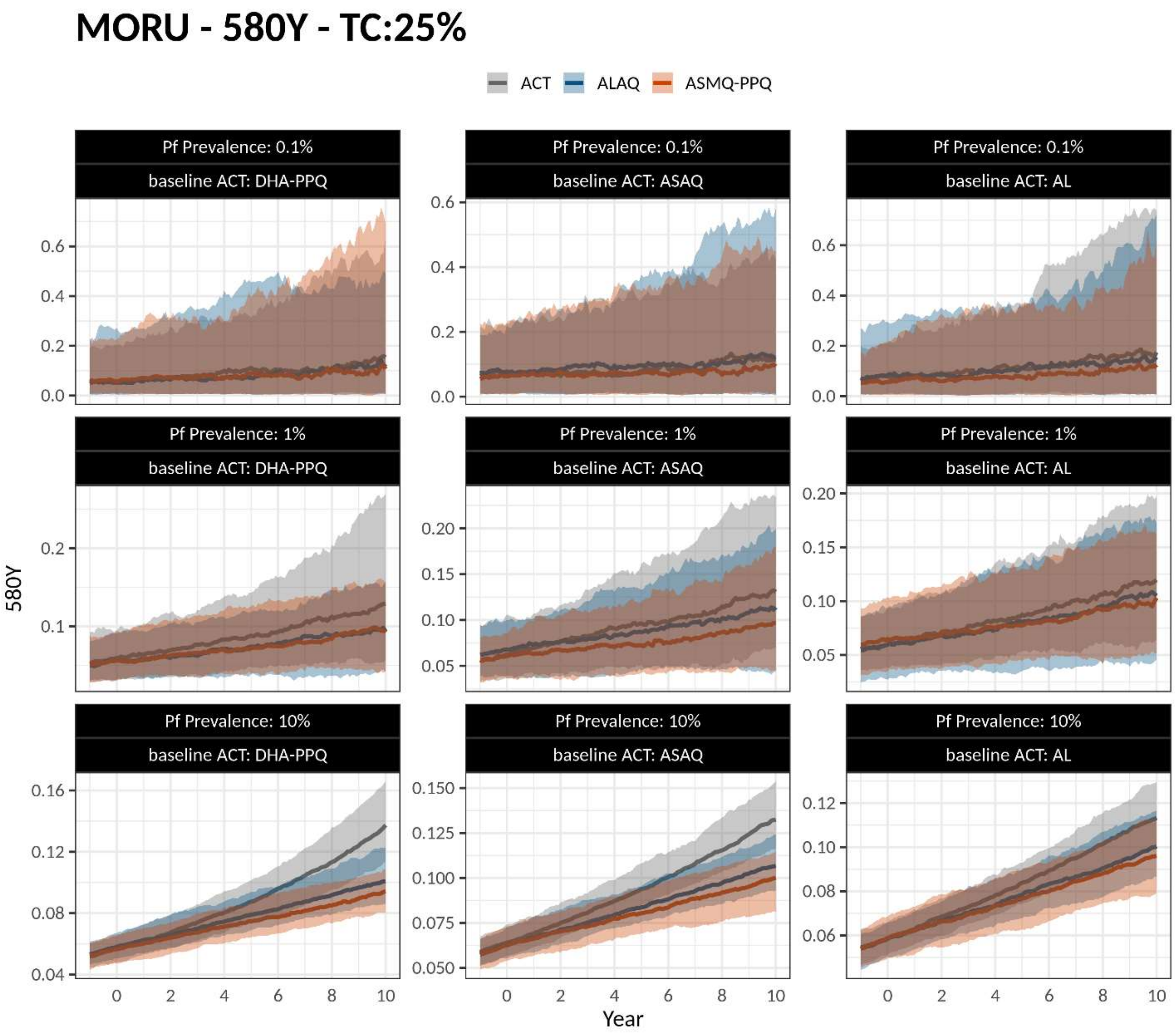

**Figure S3.** Simulations settings as in Figure S2. Figure title shows model used, outcome tracked, and treatment coverage (TC) level.

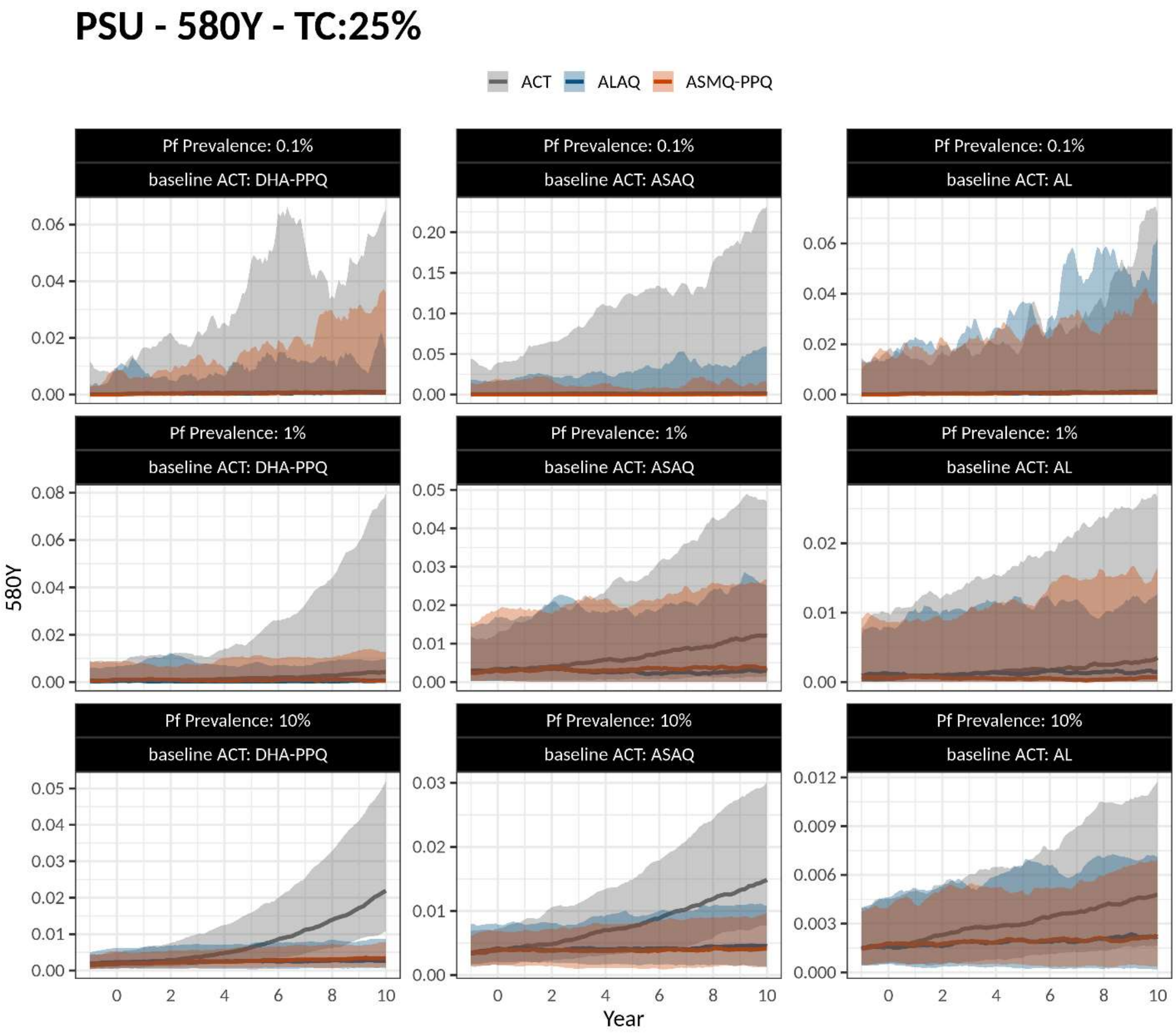

**Figure S4.** Simulations settings as in Figure S2. Figure title shows model used, outcome tracked, and treatment coverage (TC) level.

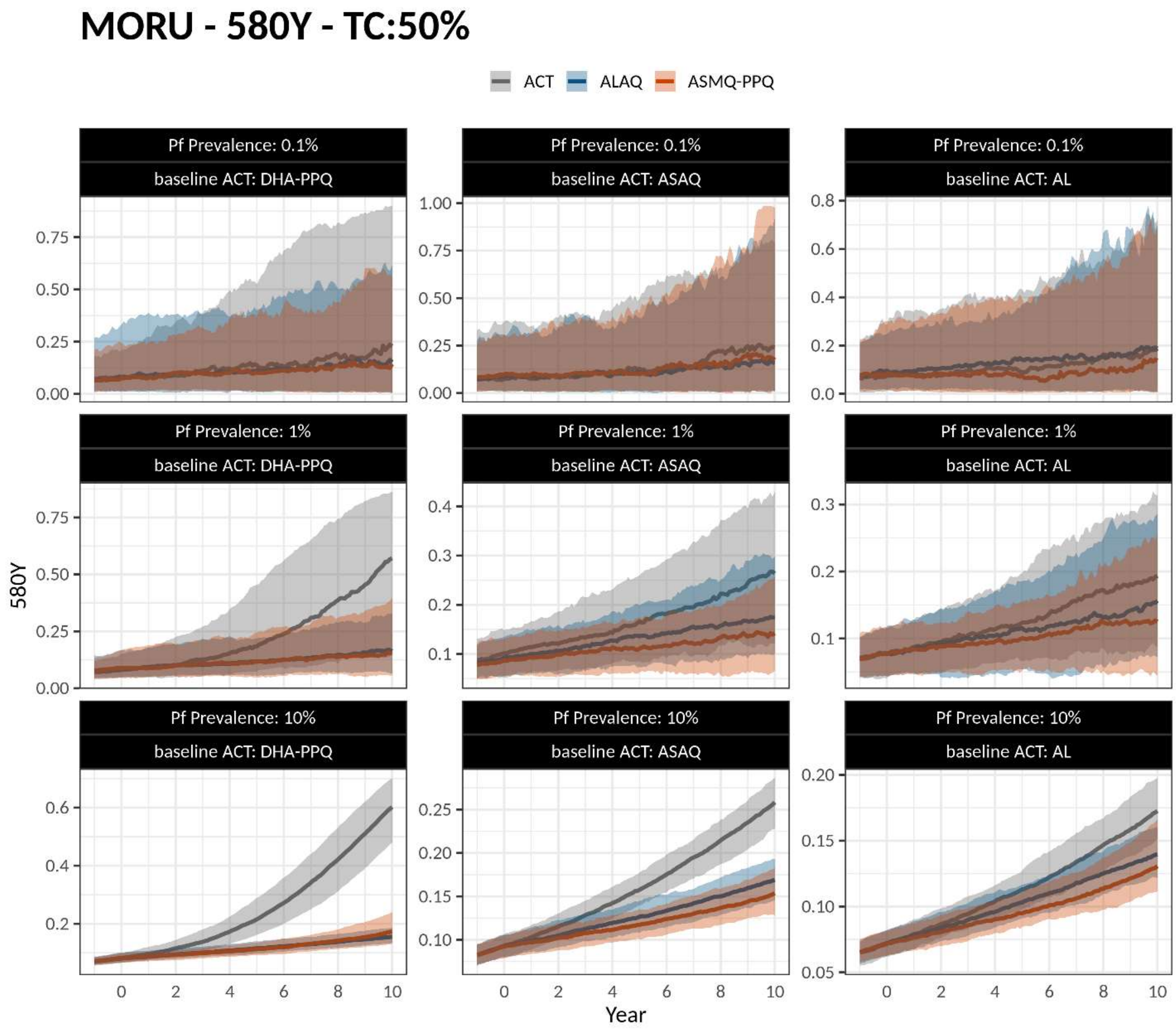

**Figure S5.** Simulations settings as in Figure S2. Figure title shows model used, outcome tracked, and treatment coverage (TC) level.

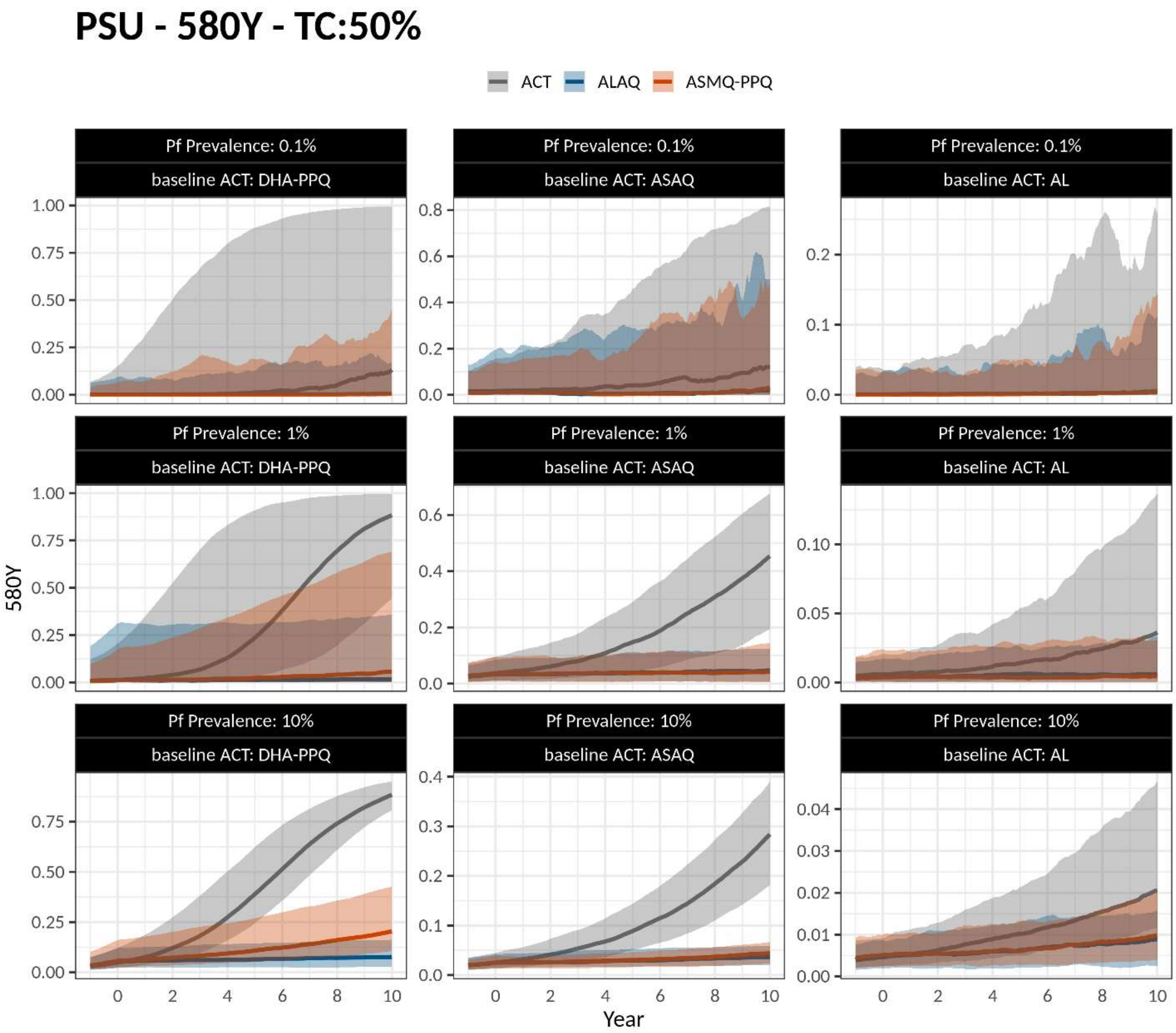

**Figure S6.** Simulations settings as in Figure S2. Figure title shows model used, outcome tracked, and treatment coverage (TC) level.

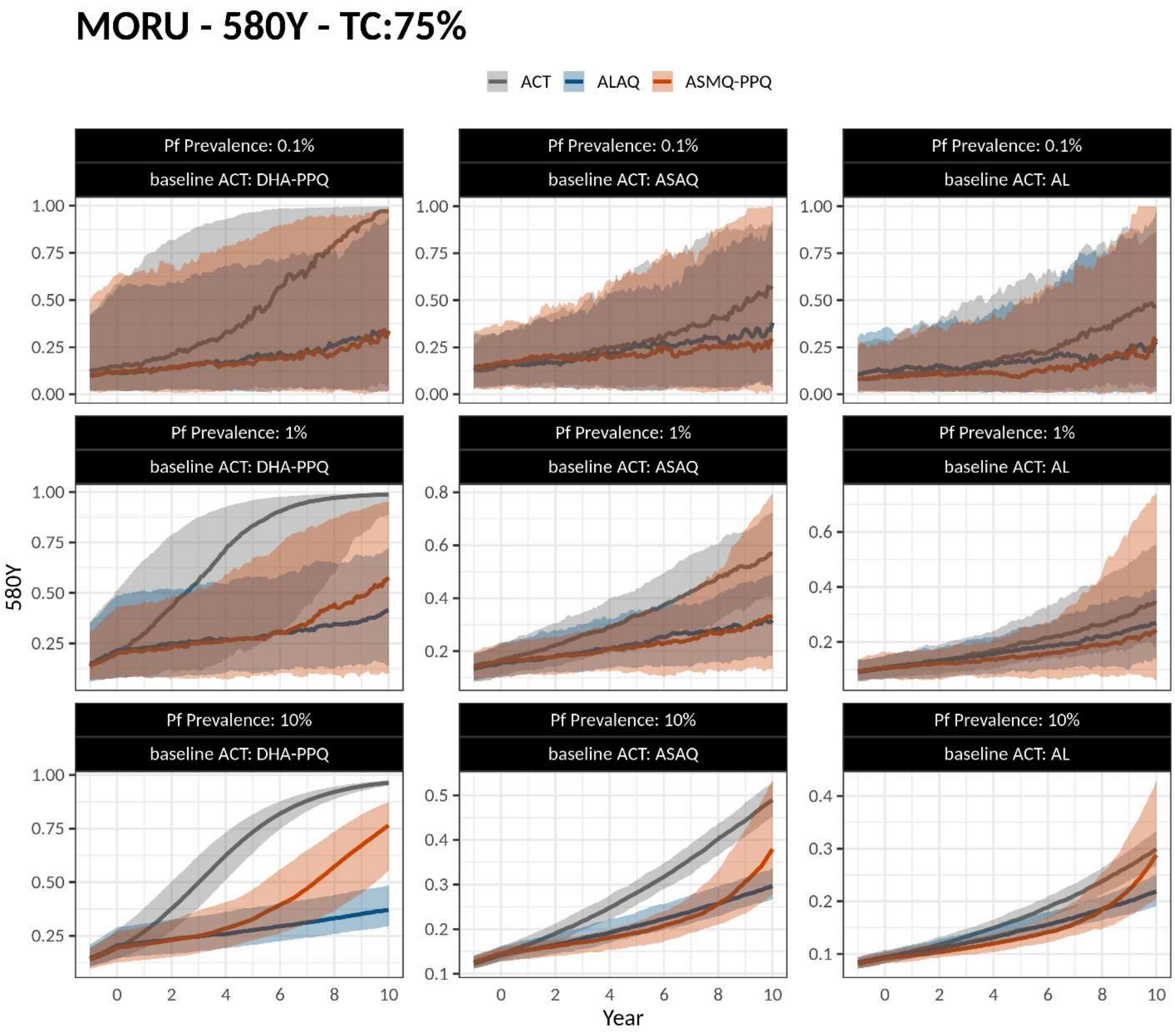

**Figure S7.** Simulations settings as in Figure S2. Figure title shows model used, outcome tracked, and treatment coverage (TC) level.

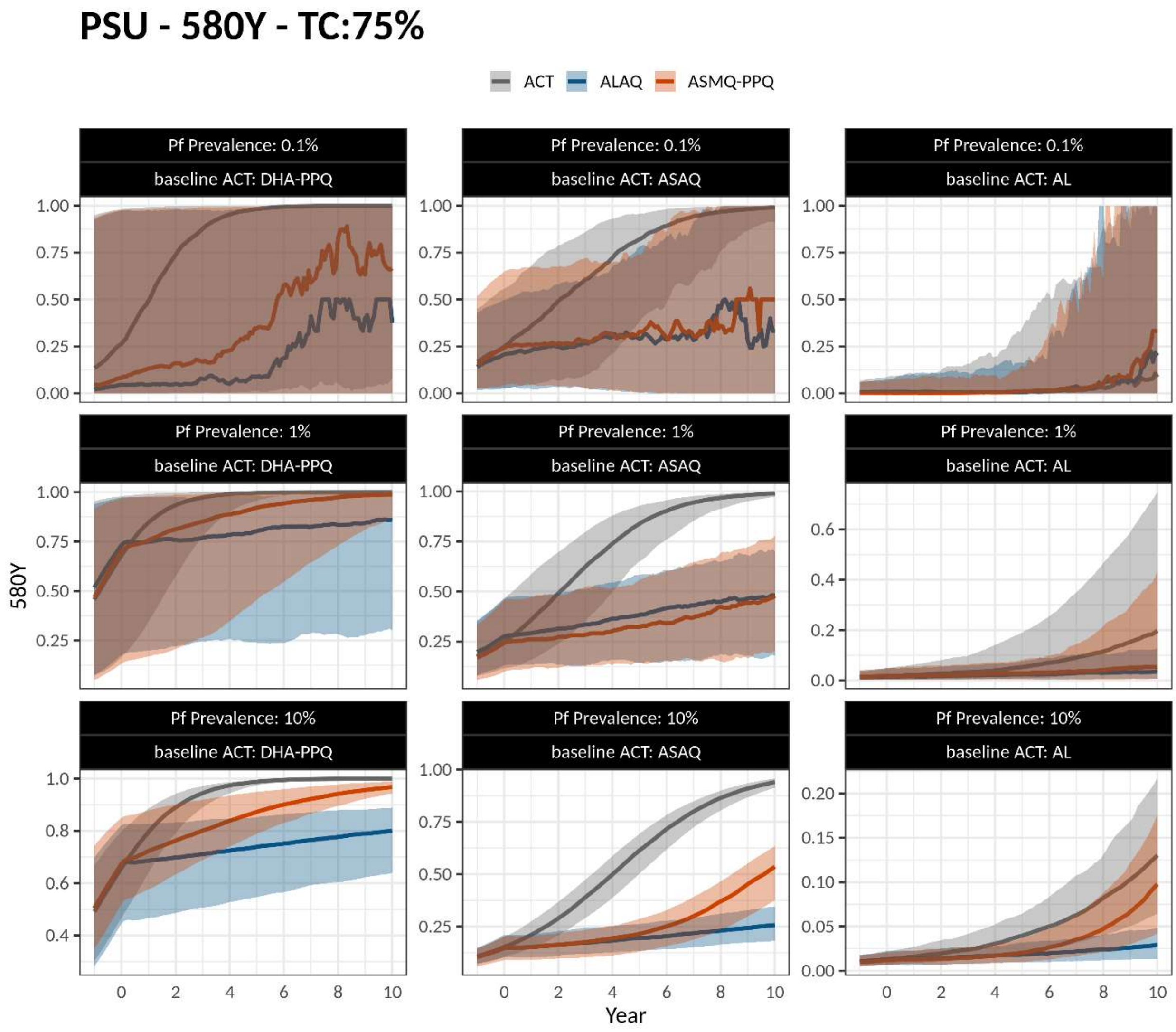

**Figure S8.** Simulations settings as in Figure S2. Figure title shows model used, outcome tracked, and treatment coverage (TC) level. Panels show monthly treatment failure rate (TFR) over 10 years.

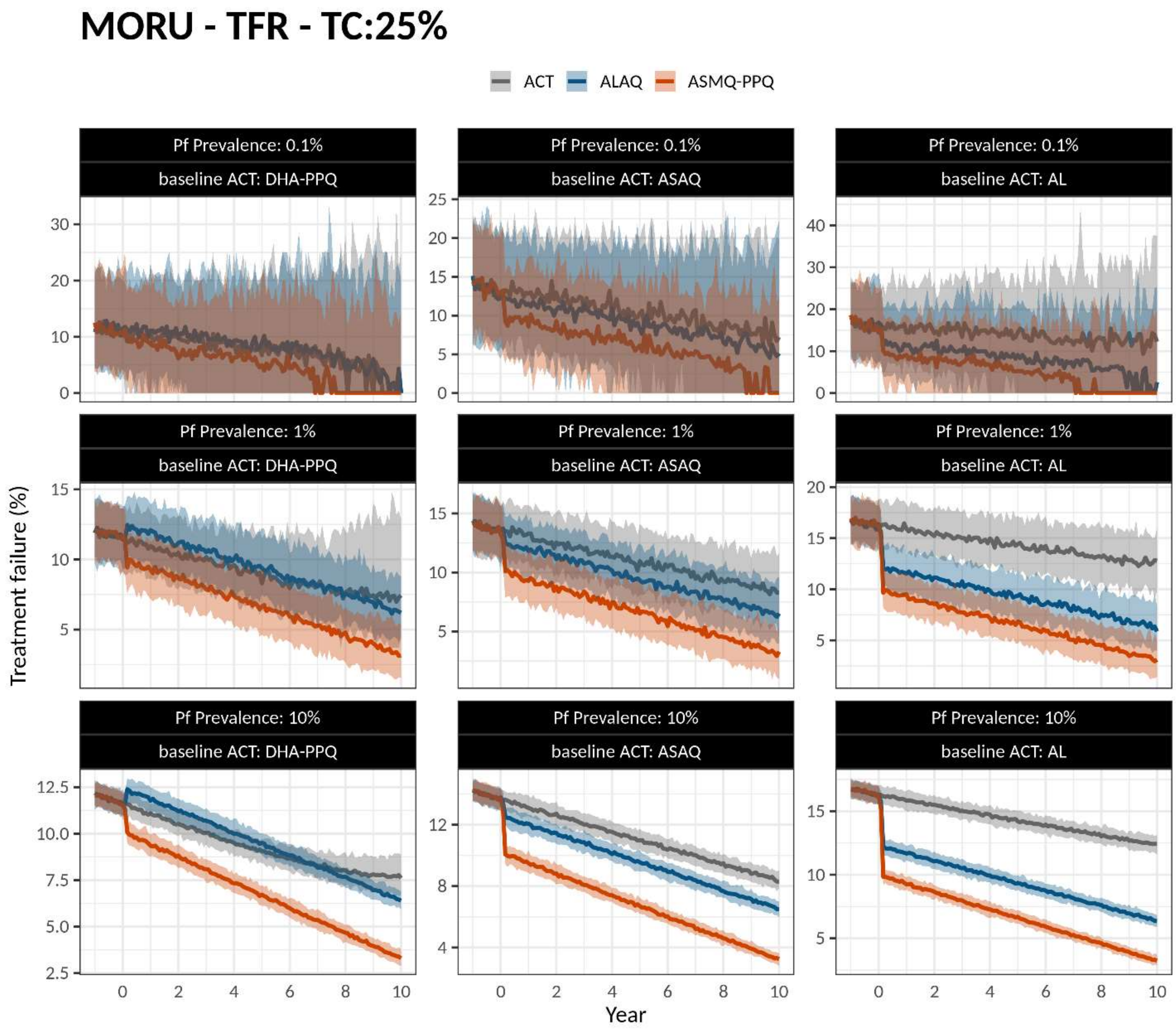

**Figure S9.** Simulations settings as in Figure S2. Figure title shows model used, outcome tracked, and treatment coverage (TC) level. Panels show monthly treatment failure rate (TFR) over 10 years.

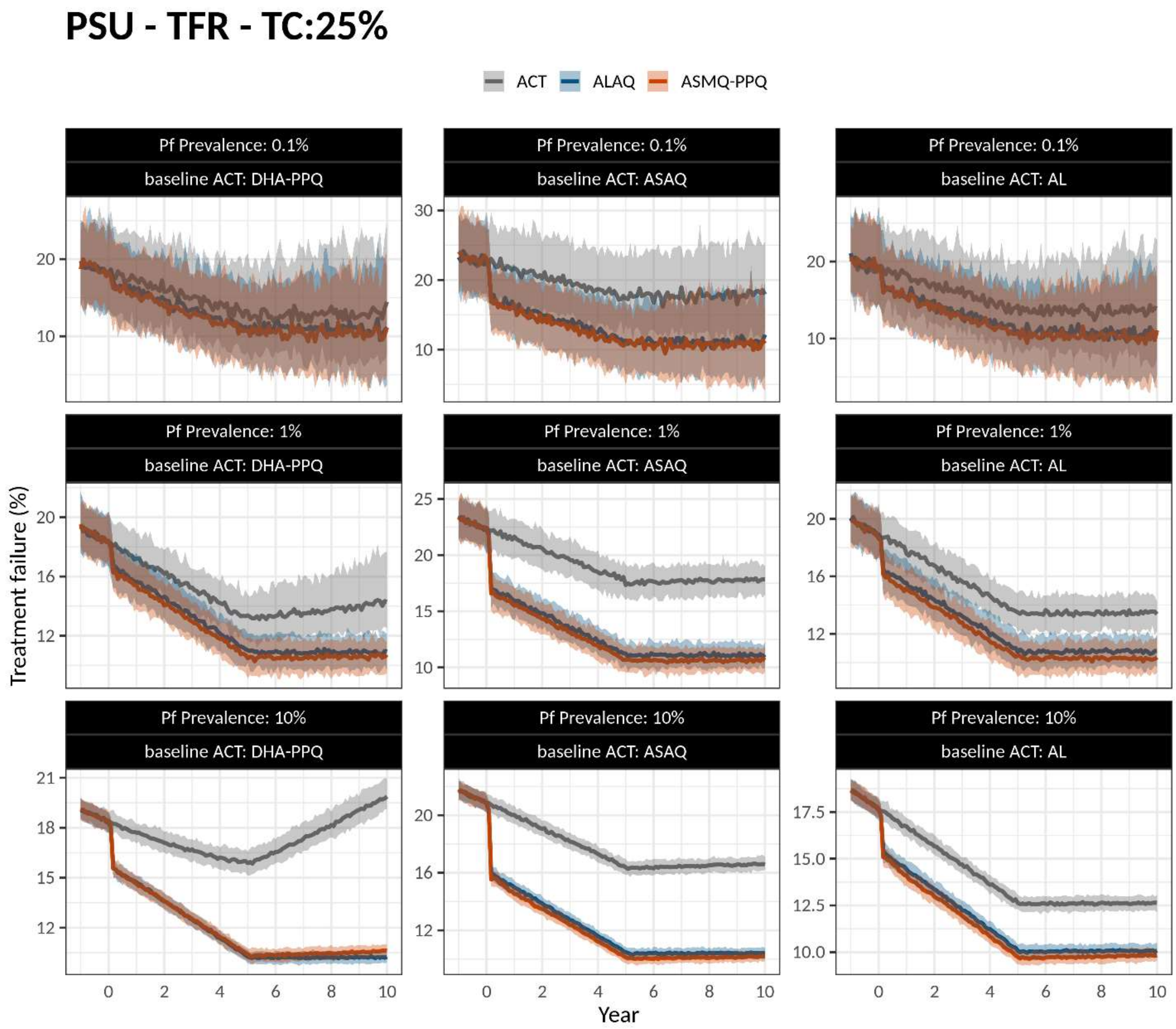

**Figure S10.** Simulations settings as in Figure S2. Figure title shows model used, outcome tracked, and treatment coverage (TC) level. Panels show monthly treatment failure rate (TFR) over 10 years.

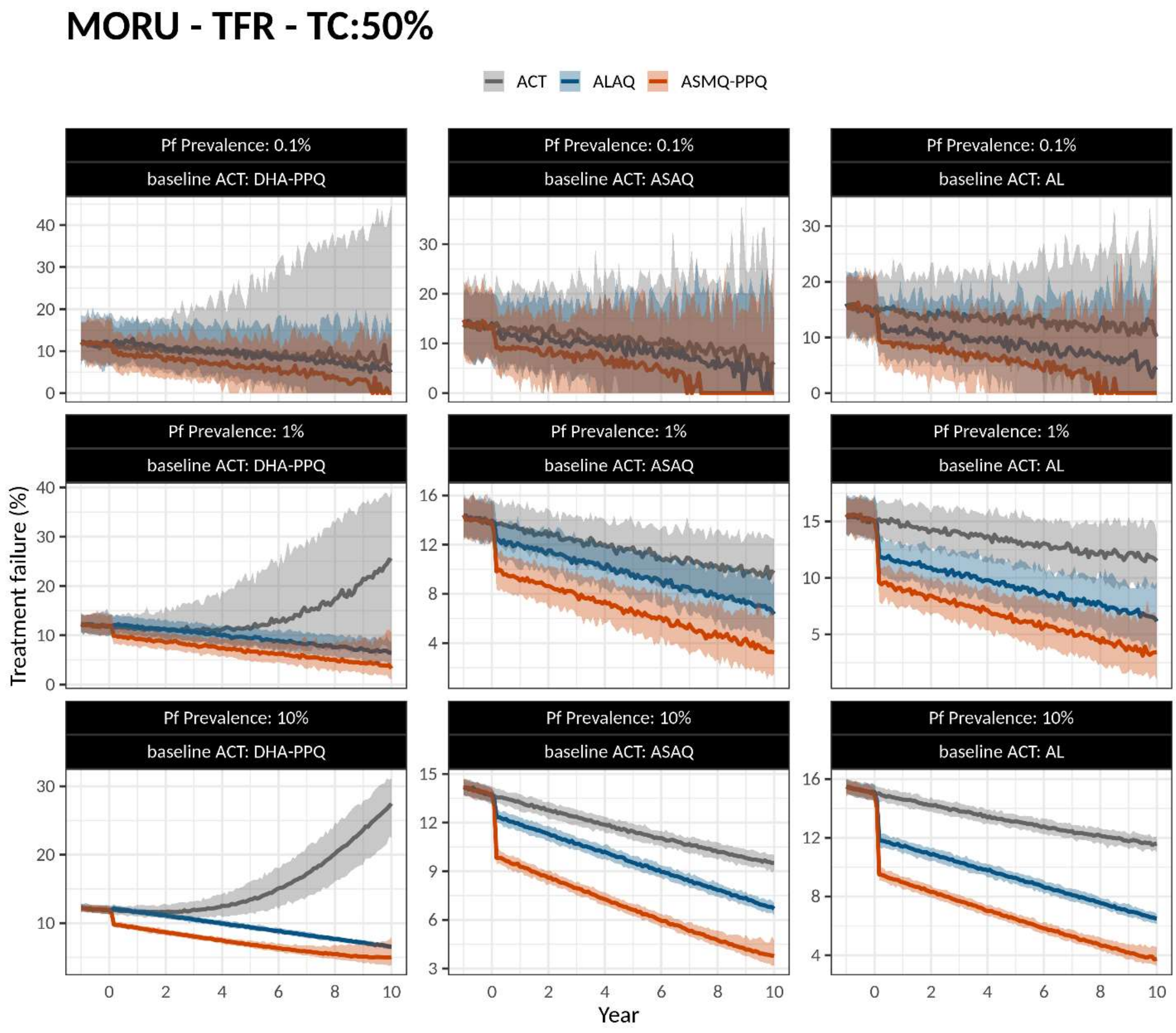

**Figure S11.** Simulations settings as in Figure S2. Figure title shows model used, outcome tracked, and treatment coverage (TC) level. Panels show monthly treatment failure rate (TFR) over 10 years.

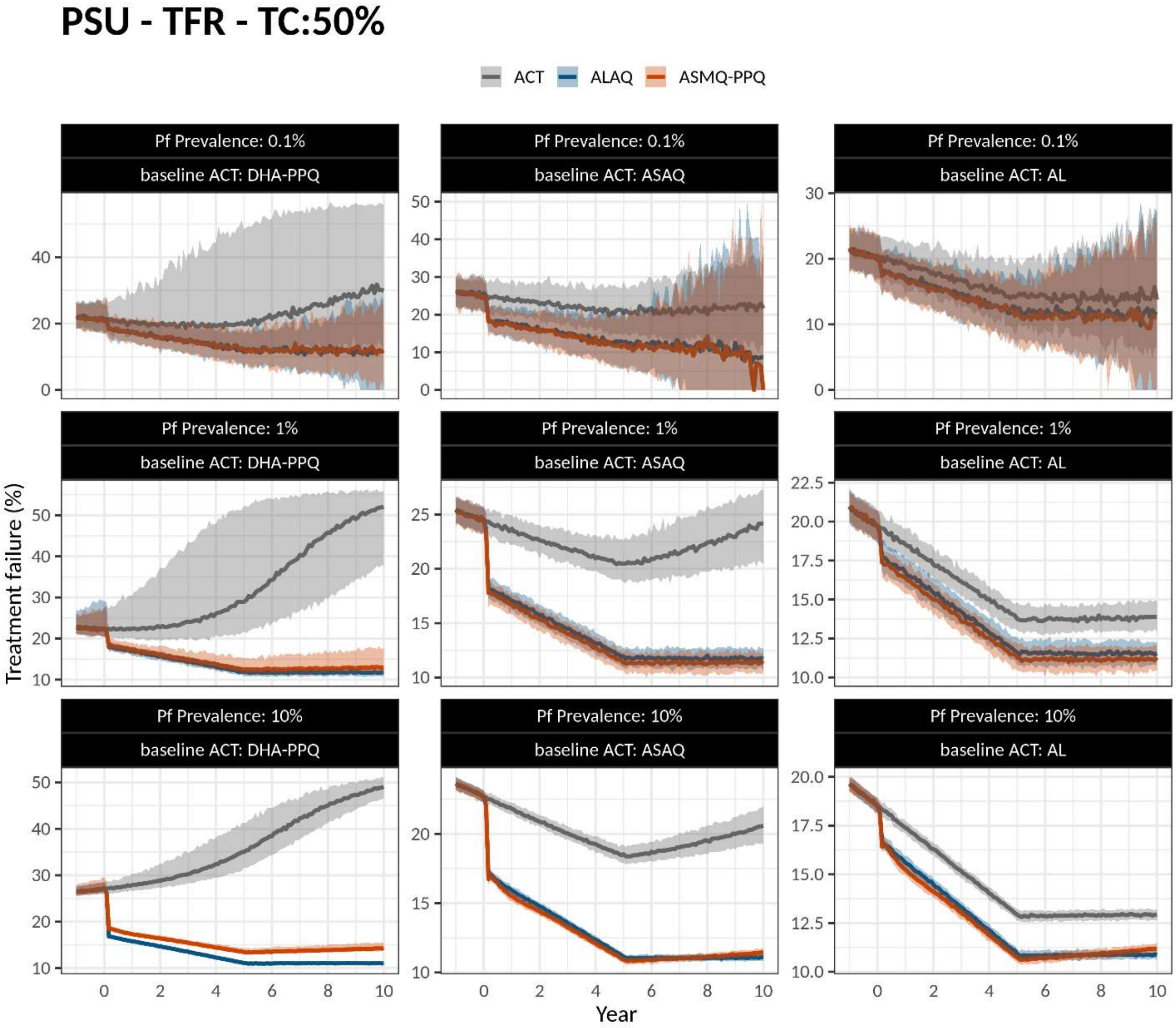

**Figure S12.** Simulations settings as in Figure S2. Figure title shows model used, outcome tracked, and treatment coverage (TC) level. Panels show monthly treatment failure rate (TFR) over 10 years.

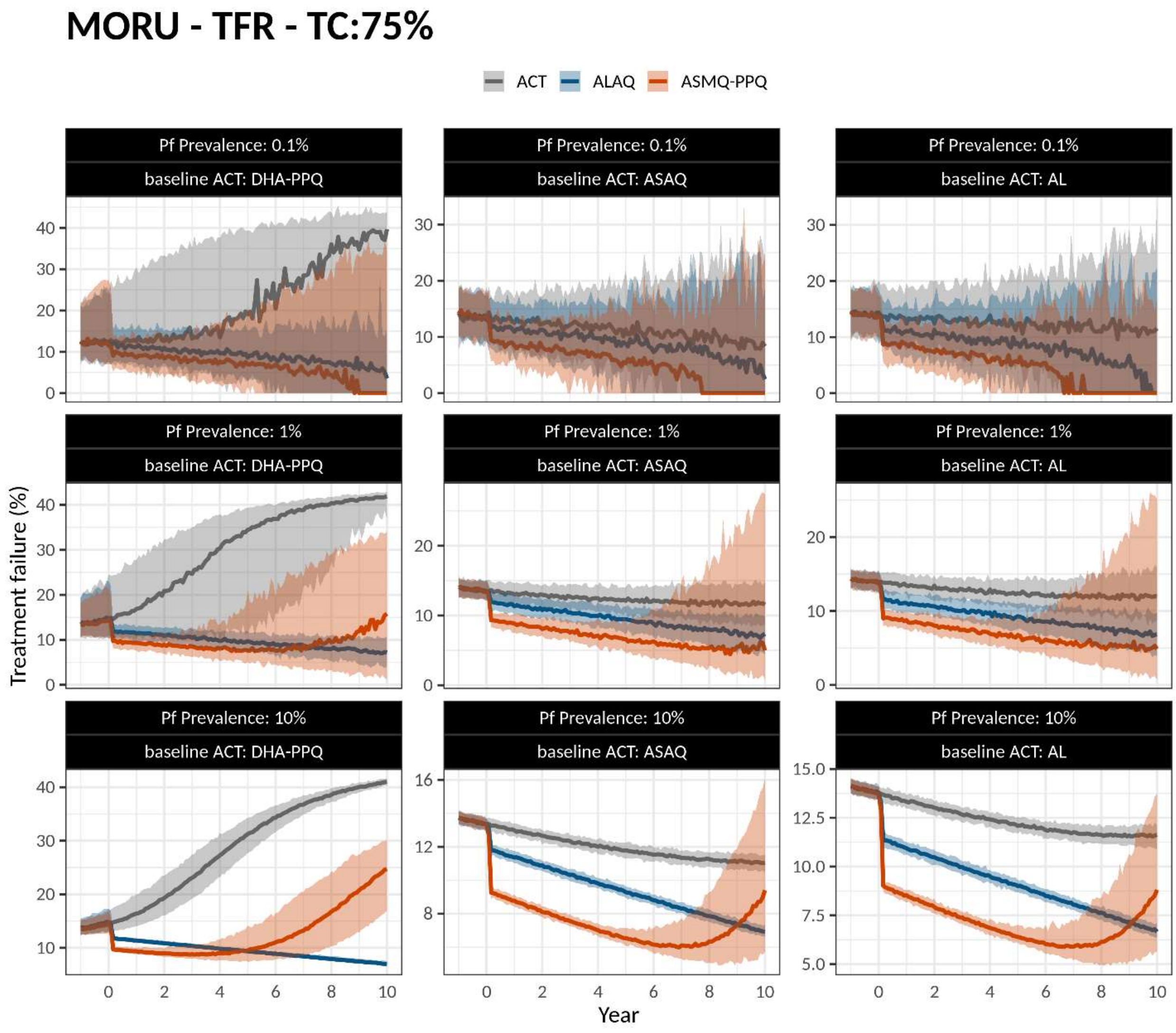

**Figure S13.** Simulations settings as in Figure S2. Figure title shows model used, outcome tracked, and treatment coverage (TC) level. Panels show monthly treatment failure rate (TFR) over 10 years.

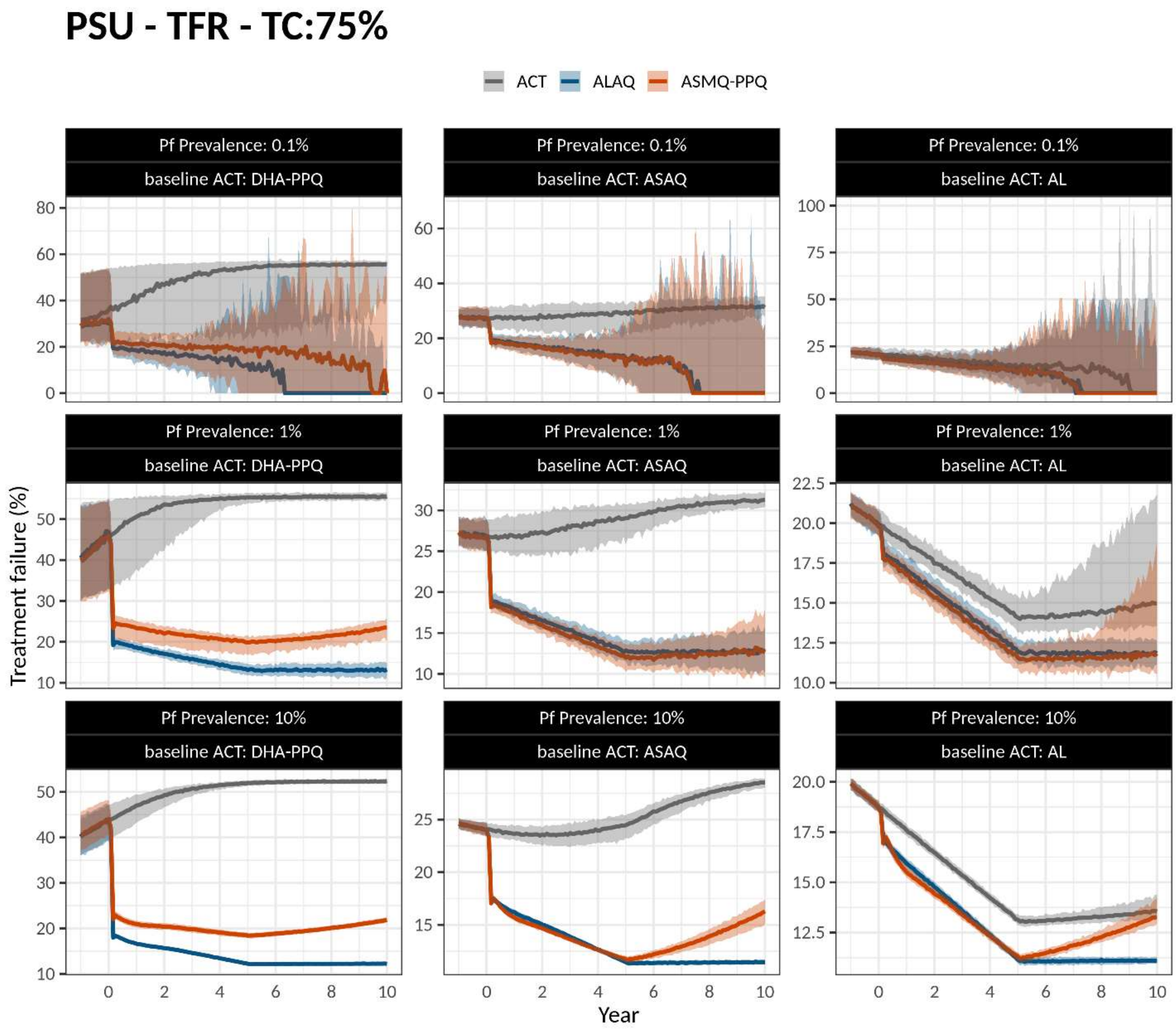

**Figure S14.** Simulations settings as in Figure S2. Figure title shows model used, outcome tracked, and treatment coverage (TC) level. Panels show all-age prevalence as measured by microscopy (*PfPR*) over 10 years.

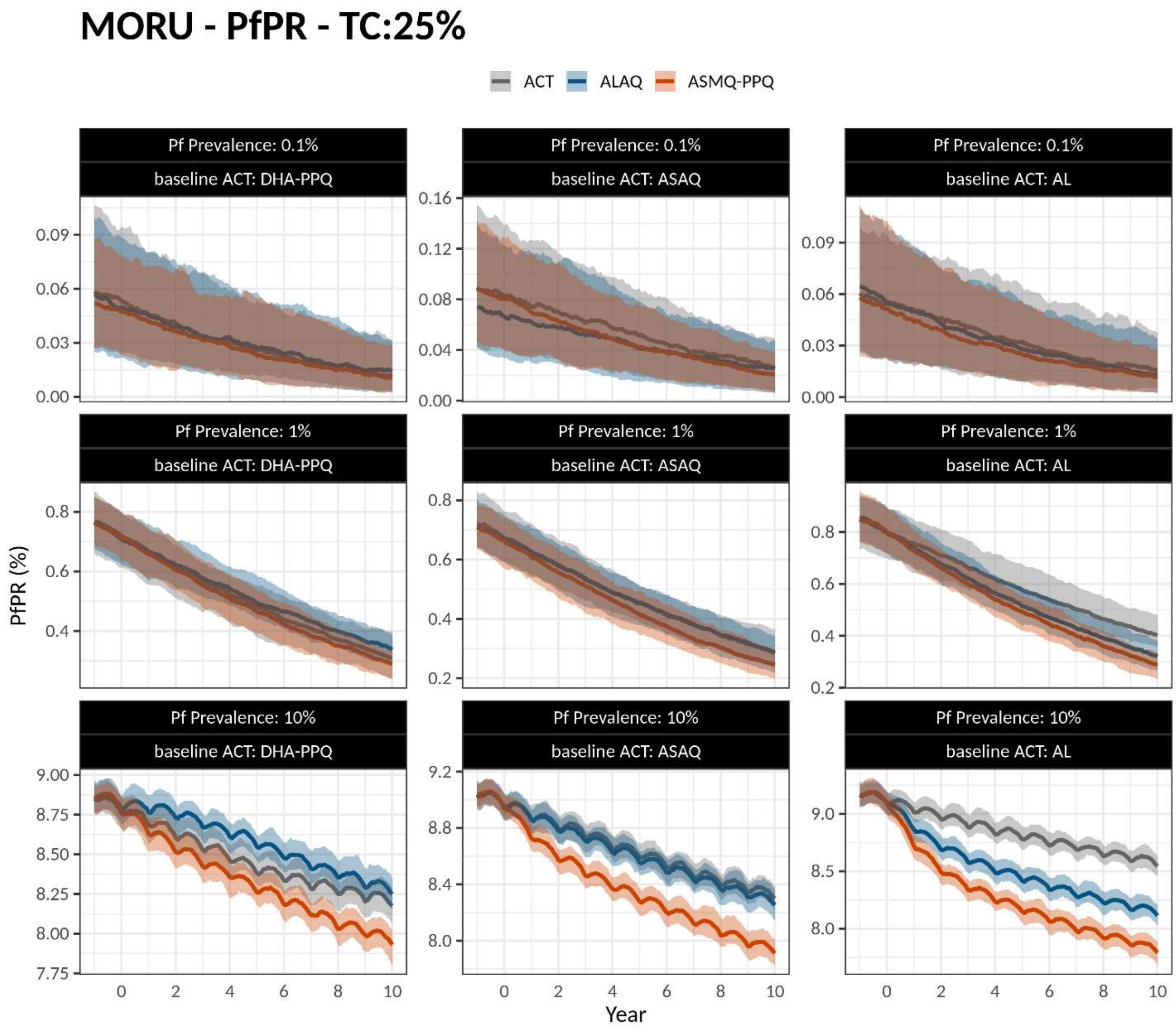

**Figure S15.** Simulations settings as in Figure S2. Figure title shows model used, outcome tracked, and treatment coverage (TC) level. Panels show all-ages prevalence as measured by microscopy (*PfPR*) over 10 years.

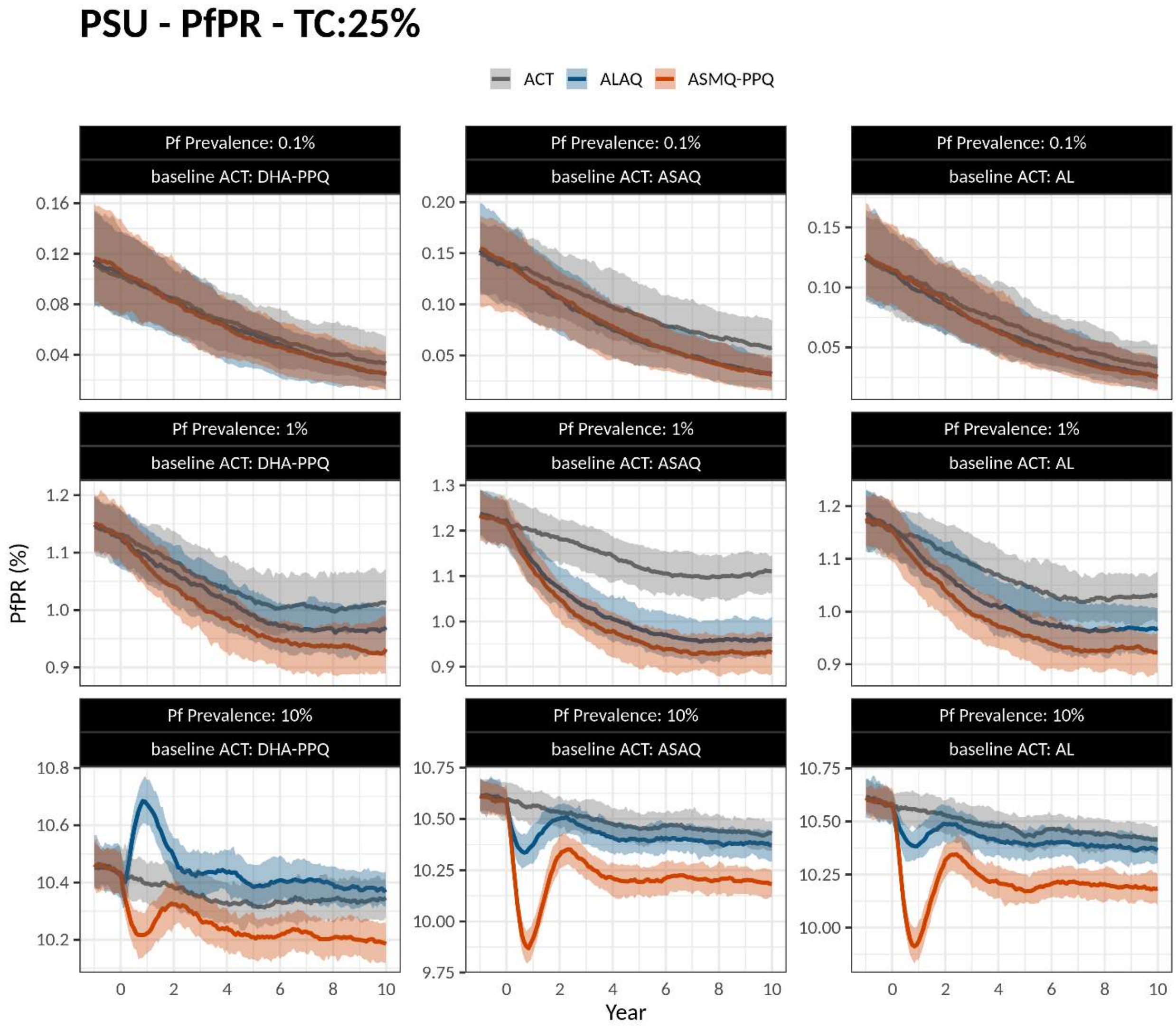

**Figure S16.** Simulations settings as in Figure S2. Figure title shows model used, outcome tracked, and treatment coverage (TC) level. Panels show all-ages *PfPR* over 10 years.

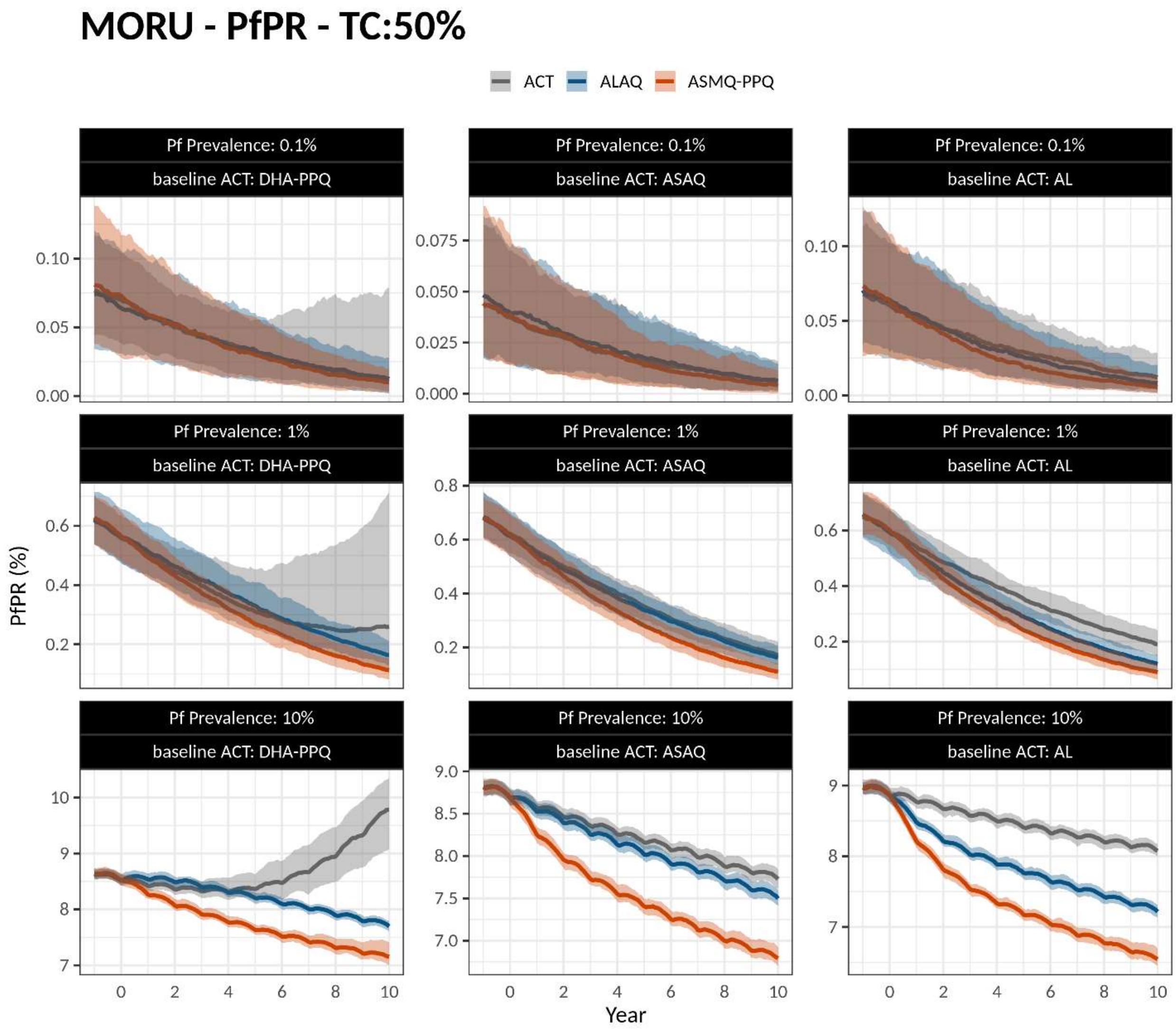

**Figure S17.** Simulations settings as in Figure S2. Figure title shows model used, outcome tracked, and treatment coverage (TC) level. Panels show all-ages *PfPR* over 10 years.

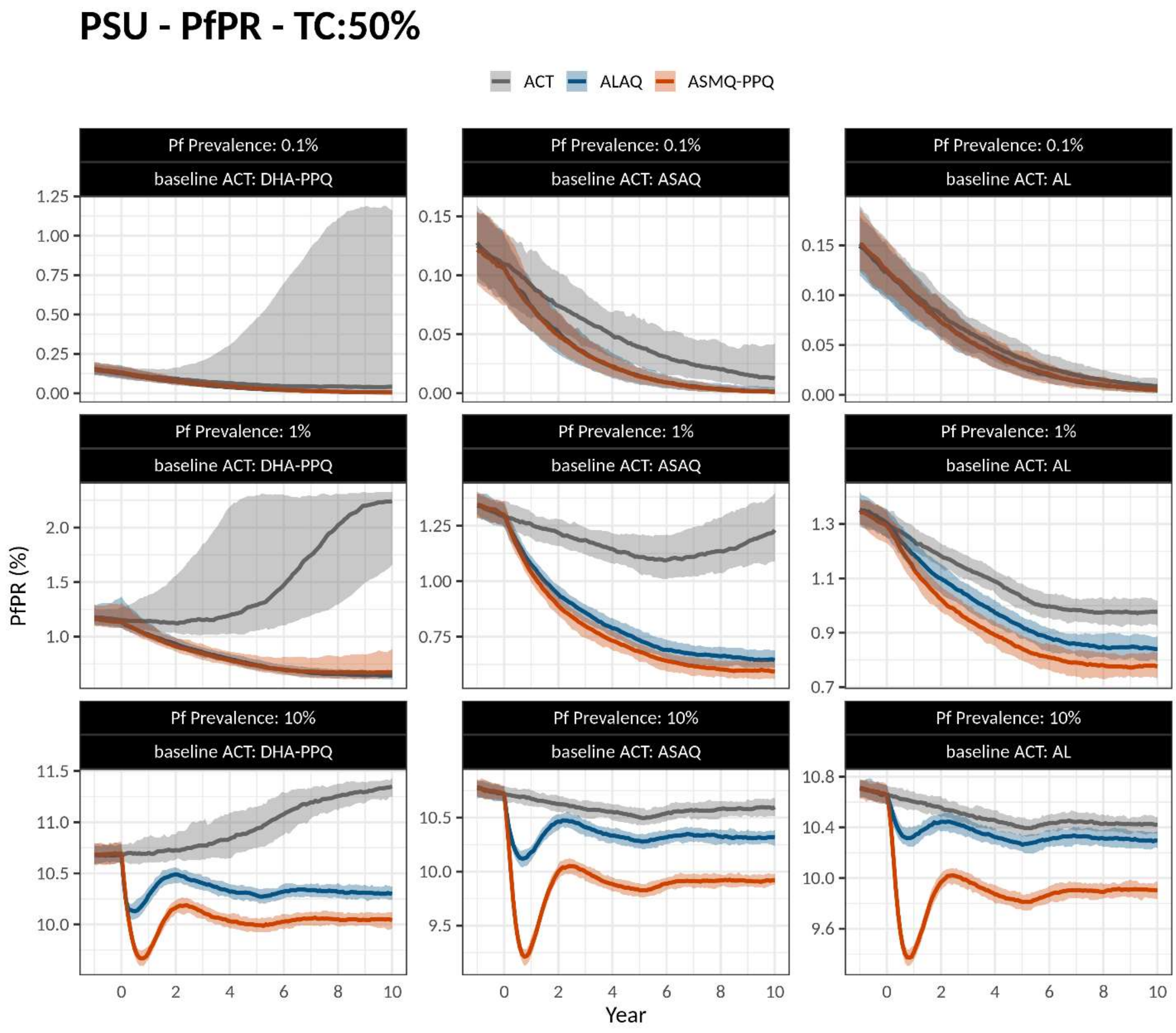

**Figure S18.** Simulations settings as in Figure S2. Figure title shows model used, outcome tracked, and treatment coverage (TC) level. Panels show all-ages *PfPR* over 10 years.

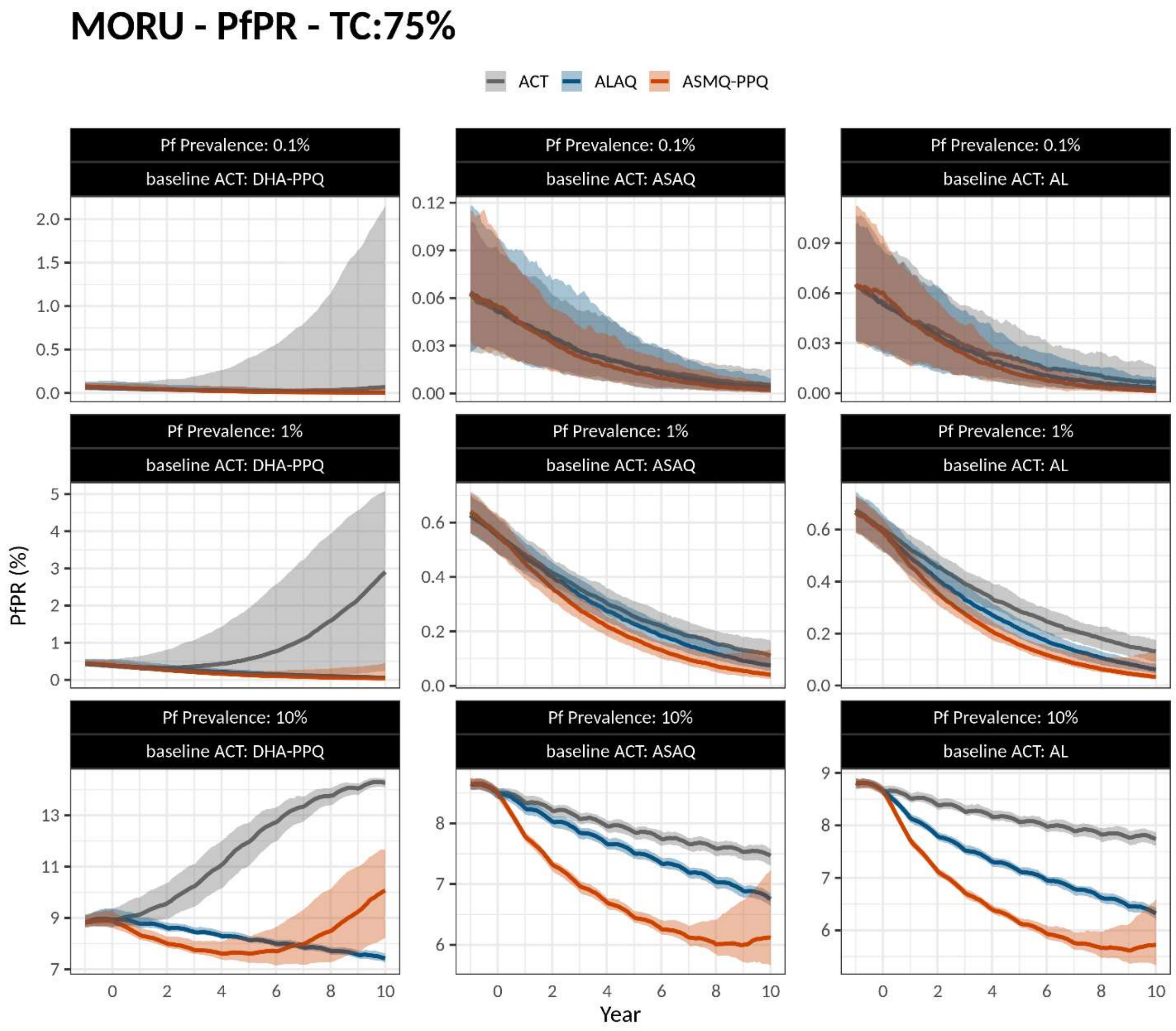

**Figure S19.** Simulations settings as in Figure S2. Figure title shows model used, outcome tracked, and treatment coverage (TC) level. Panels show all-ages *PfPR* over 10 years.

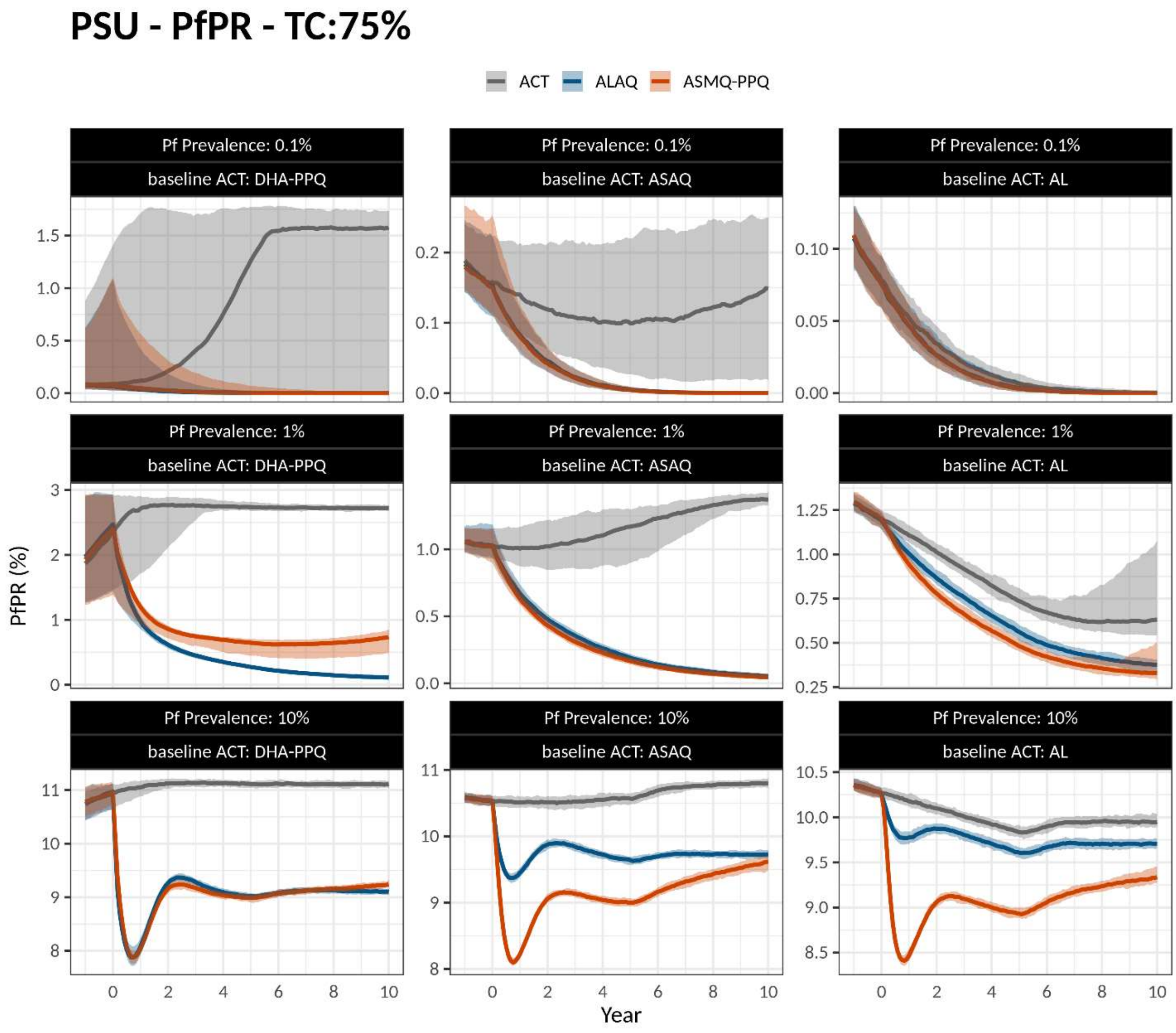

**Figure S20.** A total of 108 comparisons are done between a TACT outcome (here 580Y frequency at year 10) and continued ACT deployment: 3 different baseline ACTs of DHA-PPQ, ASAQ, AL; 3 different treatment coverages; 3 different transmission settings; 2 TACTs; and 2 models used. The Mann-Whitney U test was used to compare the 580Y frequency at year 10 between continued baseline ACTs use and ASMQ-PPQ use (left graph) or ALAQ use (right graph). Panels show the Mann-Whitney p-value for each comparison, and the colors show the results from the two models. All p-values lower than $10^{-6}$ are aggregated at the bottom-most tick on the graph.

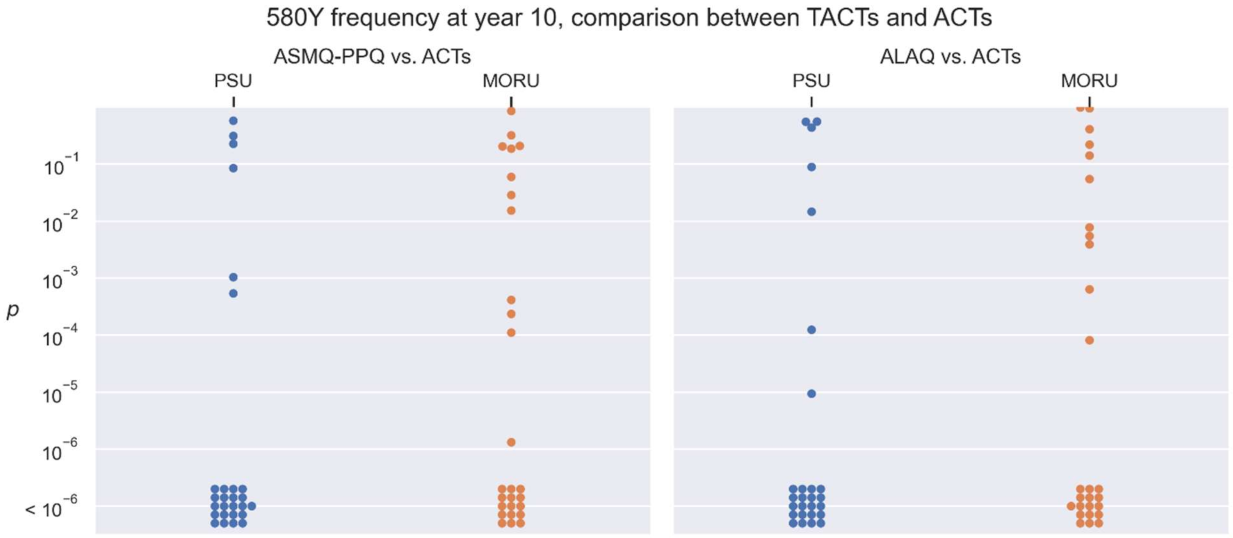

**Figure S21.** A total of 108 comparisons are done between a TACT outcome (here, the treatment failure rate at year 10) and continued ACT deployment: 3 different baseline ACTs of DHA-PPQ, ASAQ, AL; 3 different treatment coverages; 3 different transmission settings; 2 TACTs; and 2 models used. The Mann-Whitney U test was used to compare the 580Y frequency at year 10 between continued baseline ACTs use and ASMQ-PPQ use (left graph) or ALAQ use (right graph). Panels show the Mann-Whitney p-value for this comparison, and the colors show the results from the two models. All p-values lower than $10^{-6}$ are aggregated at the bottom-most tick on the graph.

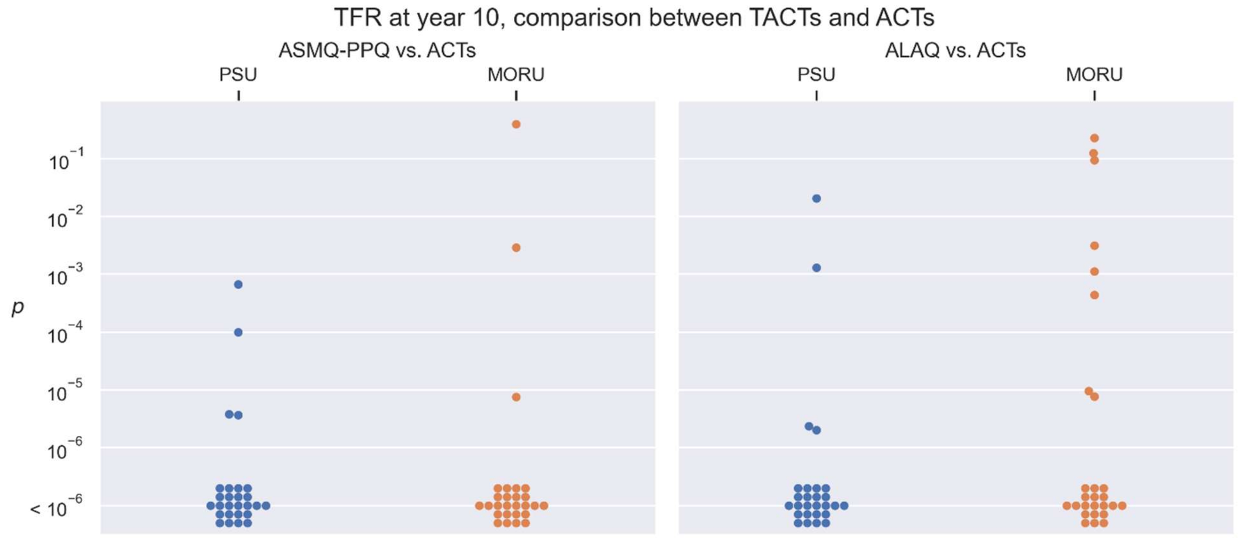

**Figure S22.** 580Y frequency 10 years after TACT deployment or continued ACT deployment, in three baseline scenarios of ACT use (columns) and three prevalence settings (rows). Treatment coverage is 25%; The leftmost pair of boxplots in each panel (in gray) show the 580Y frequency ten years later under a status quo ACT policy. The blue (ALAQ) and red (ASMQ-PPQ) boxplots show 580Y frequency outcomes after 10 years of a TACT policy. Boxplot pairs have MORU model results on the left and PSU model results on the right. All boxplots summarize 100 simulations.

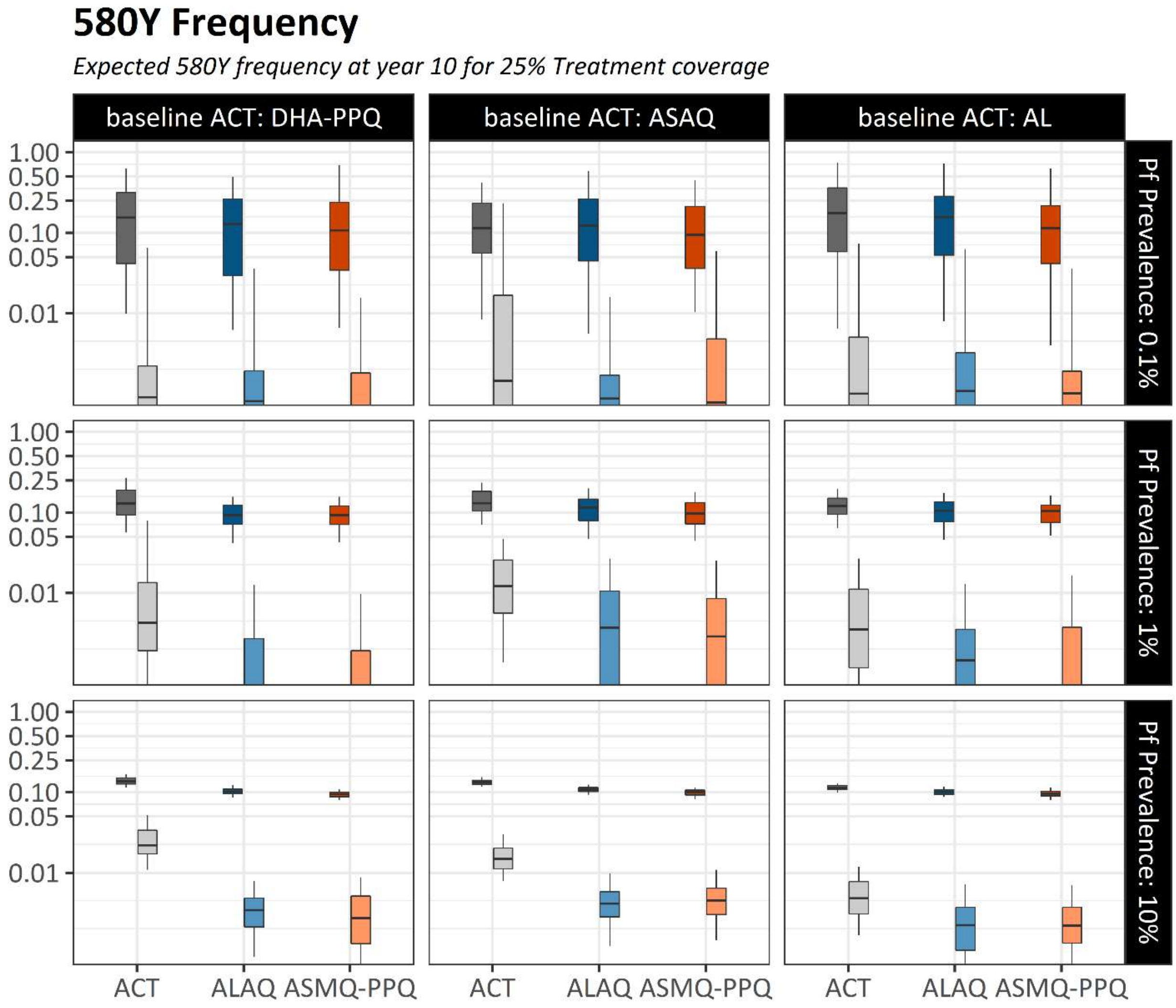

**Figure S23.** 580Y frequency 10 years after TACT deployment or continued ACT deployment, in three baseline scenarios of ACT use (columns) and three prevalence settings (rows). Treatment coverage is 75%; The leftmost pair of boxplots in each panel (in gray) show the 580Y frequency ten years later under a status quo ACT policy. The blue (ALAQ) and red (ASMQ-PPQ) boxplots show 580Y frequency outcomes after 10 years of a TACT policy. Boxplot pairs have MORU model results on the left and PSU model results on the right. All boxplots summarize 100 simulations.

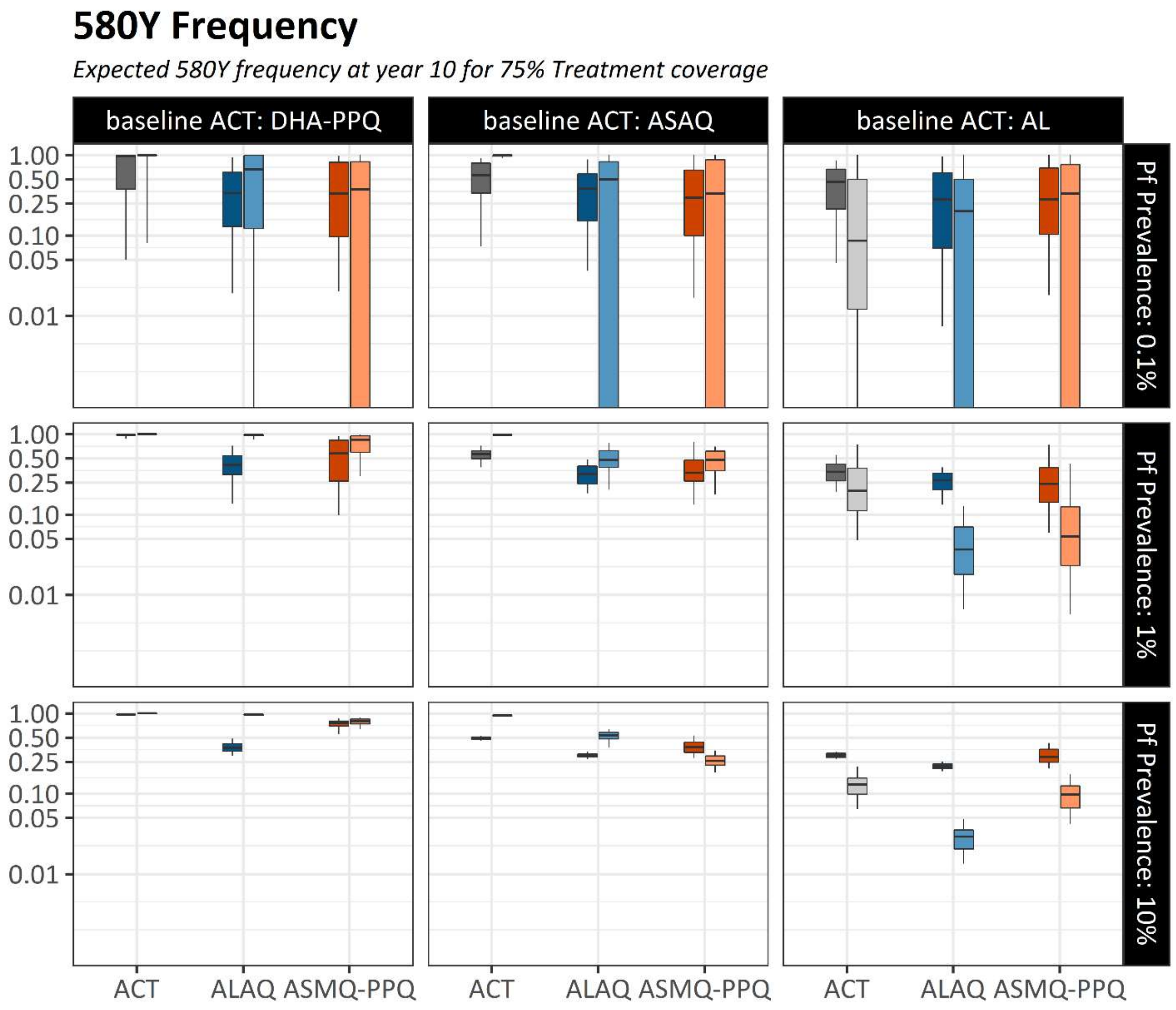

**Figure S24.** Treatment failure rates after 10 years of TACT deployment or ACT deployment, in three baseline scenarios of ACT use (columns) and three prevalence settings (rows). Treatment coverage is 25%; The leftmost pair of boxplots in each panel (in gray) show the TF rates ten years later under a status quo ACT policy. The blue (ALAQ) and red (ASMQ-PPQ) boxplots show treatment failure rate outcomes after 10 years of a TACT policy. Boxplot pairs have MORU model results on the left and PSU model results on the right. All boxplots summarize 100 simulations.

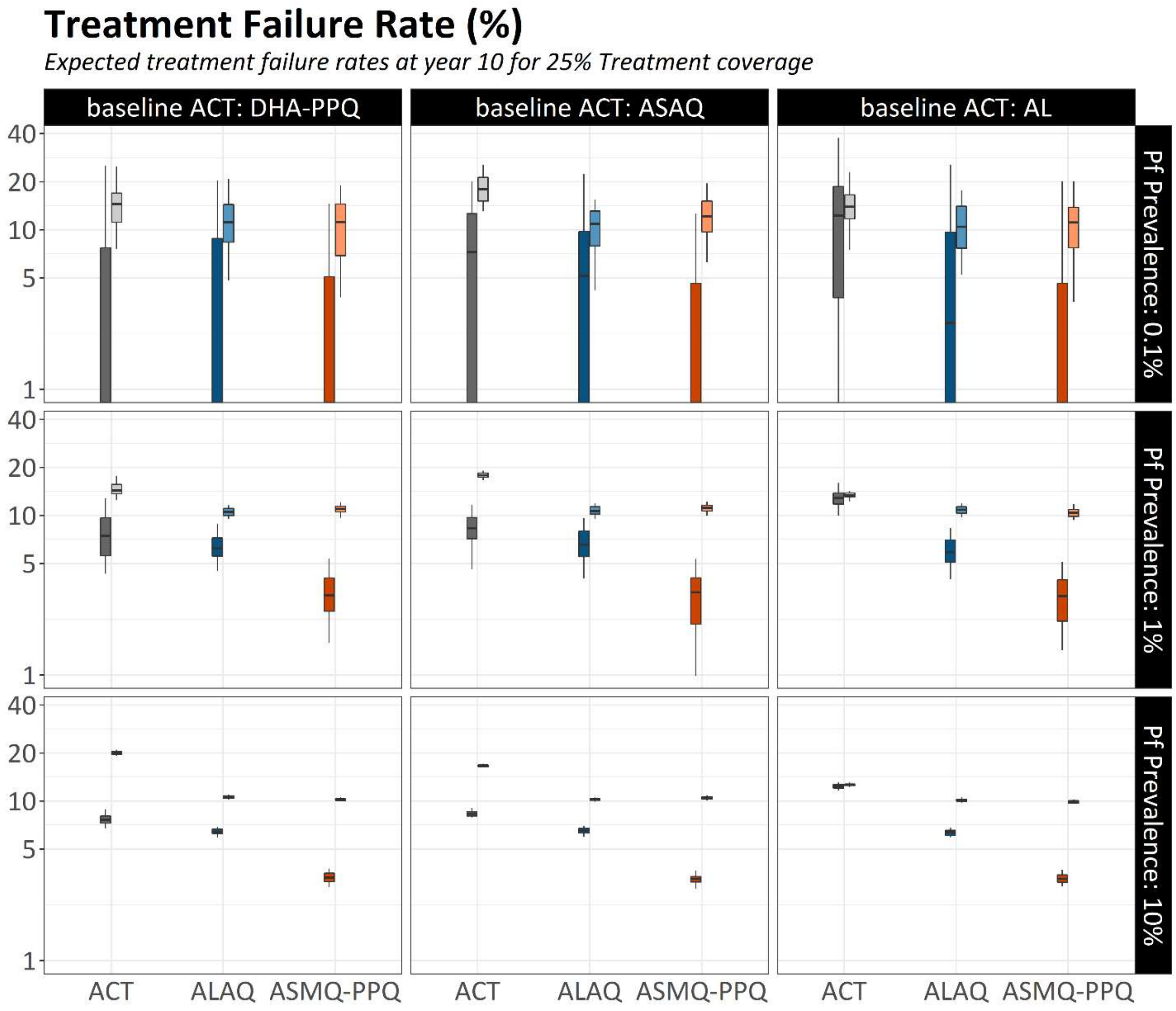

**Figure S25.** Treatment failure rates after 10 years of TACT deployment or ACT deployment, in three baseline scenarios of ACT use (columns) and three prevalence settings (rows). Treatment coverage is 75%; The leftmost pair of boxplots in each panel (in gray) show the TF rates ten years later under a status quo ACT policy. The blue (ALAQ) and red (ASMQ-PPQ) boxplots show treatment failure rate outcomes after 10 years of a TACT policy. Boxplot pairs have MORU model results on the left and PSU model results on the right. All boxplots summarize 100 simulations.

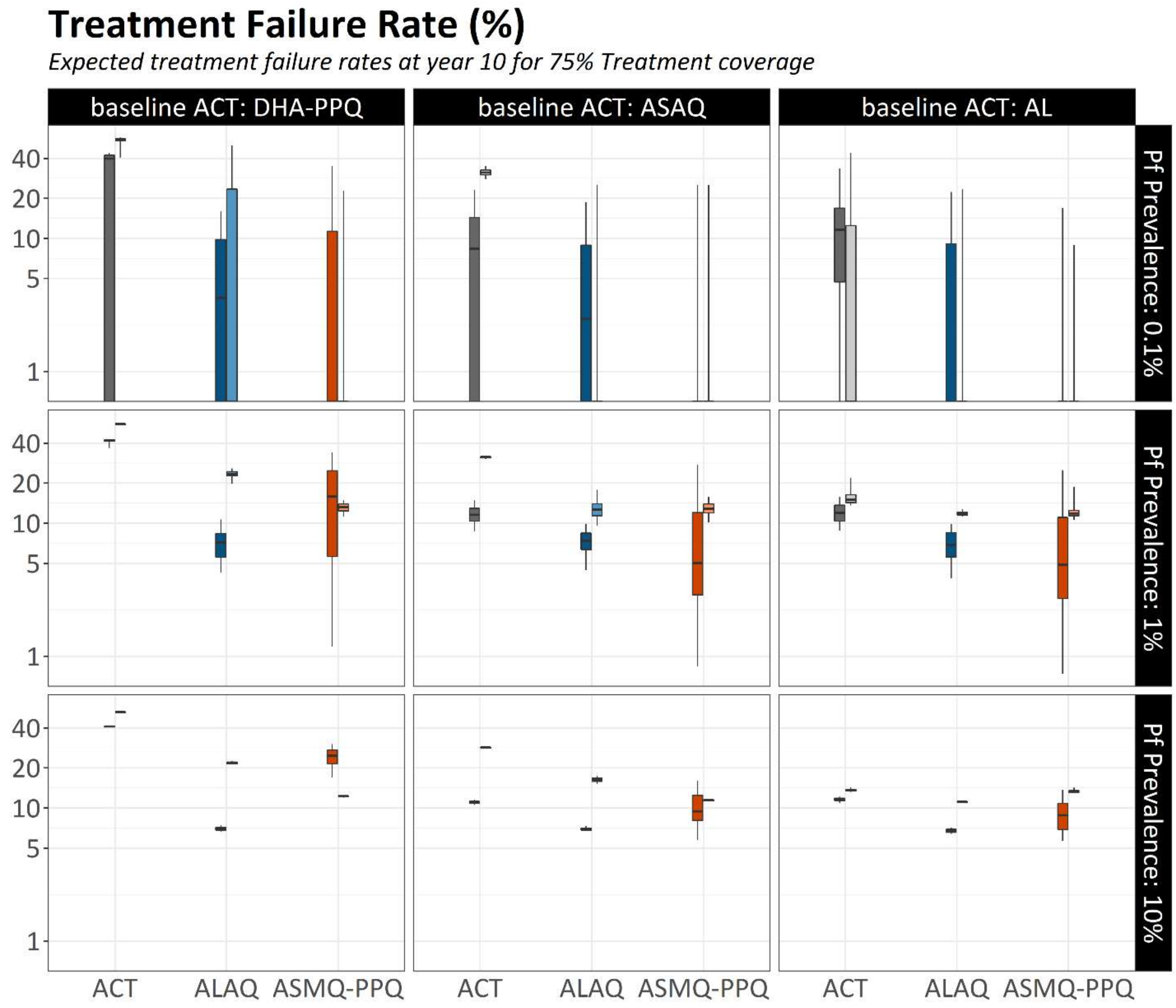

**Figure S26.** Pf prevalence after 10 years of TACT deployment or ACT deployment, in three baseline scenarios of ACT use (columns) and three prevalence settings (rows). Treatment coverage is 50%; The leftmost pair of boxplots in each panel (in gray) show Pf prevalence ten years later under a status quo ACT policy. The blue (ALAQ) and red (ASMQ-PPQ) boxplots show prevalence outcomes after 10 years of a TACT policy. Boxplot pairs have MORU model results on the left and PSU model results on the right. All boxplots summarize 100 simulations. The percent reduction in median Pf prevalence from ACT to TACT is shown next to the median of each TACT boxplot.

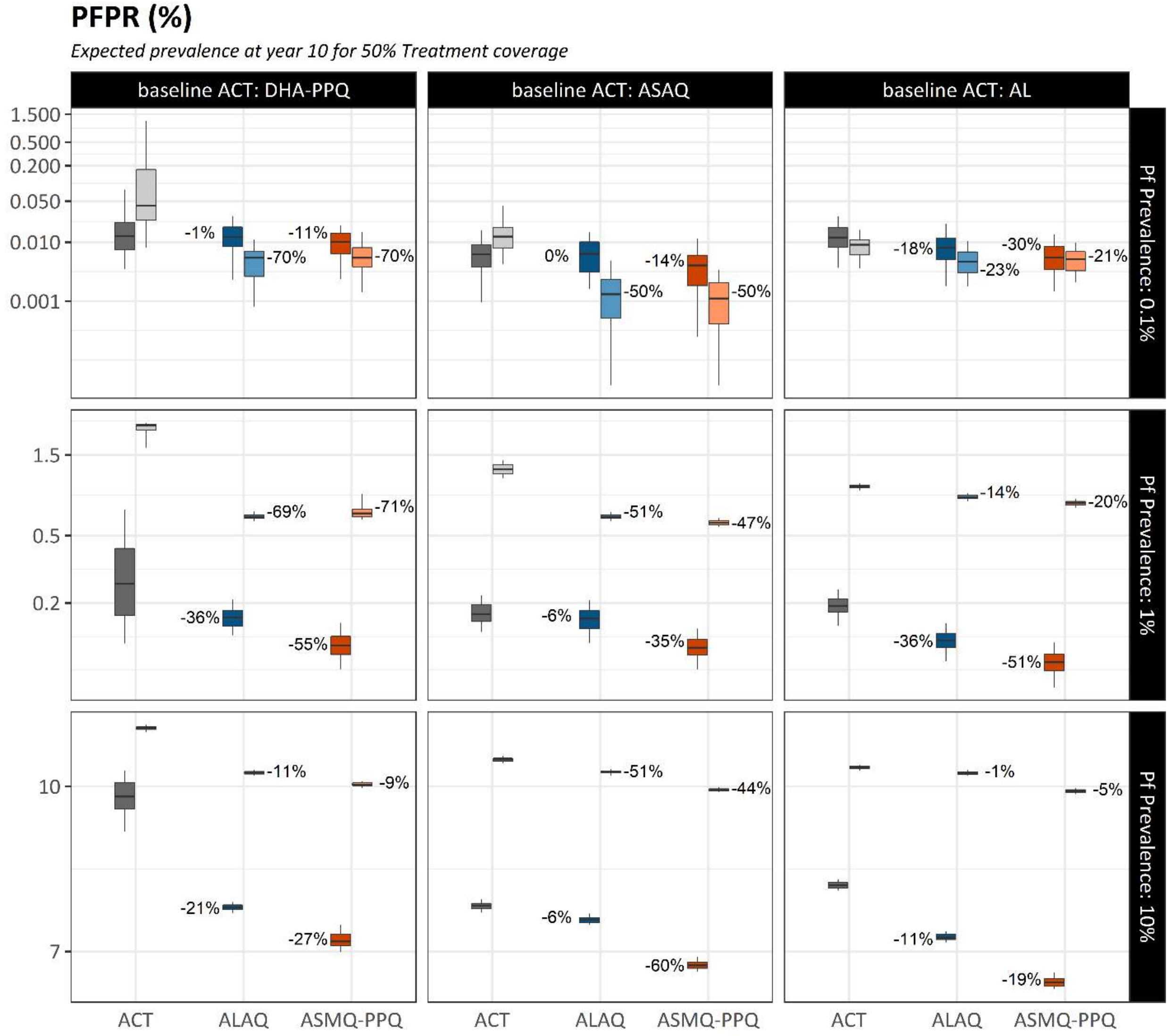

**Figure S27.** Pf prevalence after 10 years of TACT deployment or ACT deployment, in three baseline scenarios of ACT use (columns) and three prevalence settings (rows). Treatment coverage is 25%; The leftmost pair of boxplots in each panel (in gray) show Pf prevalence ten years later under a status quo ACT policy. The blue (ALAQ) and red (ASMQ-PPQ) boxplots show prevalence outcomes after 10 years of a TACT policy. Boxplot pairs have MORU model results on the left and PSU model results on the right. All boxplots summarize 100 simulations.

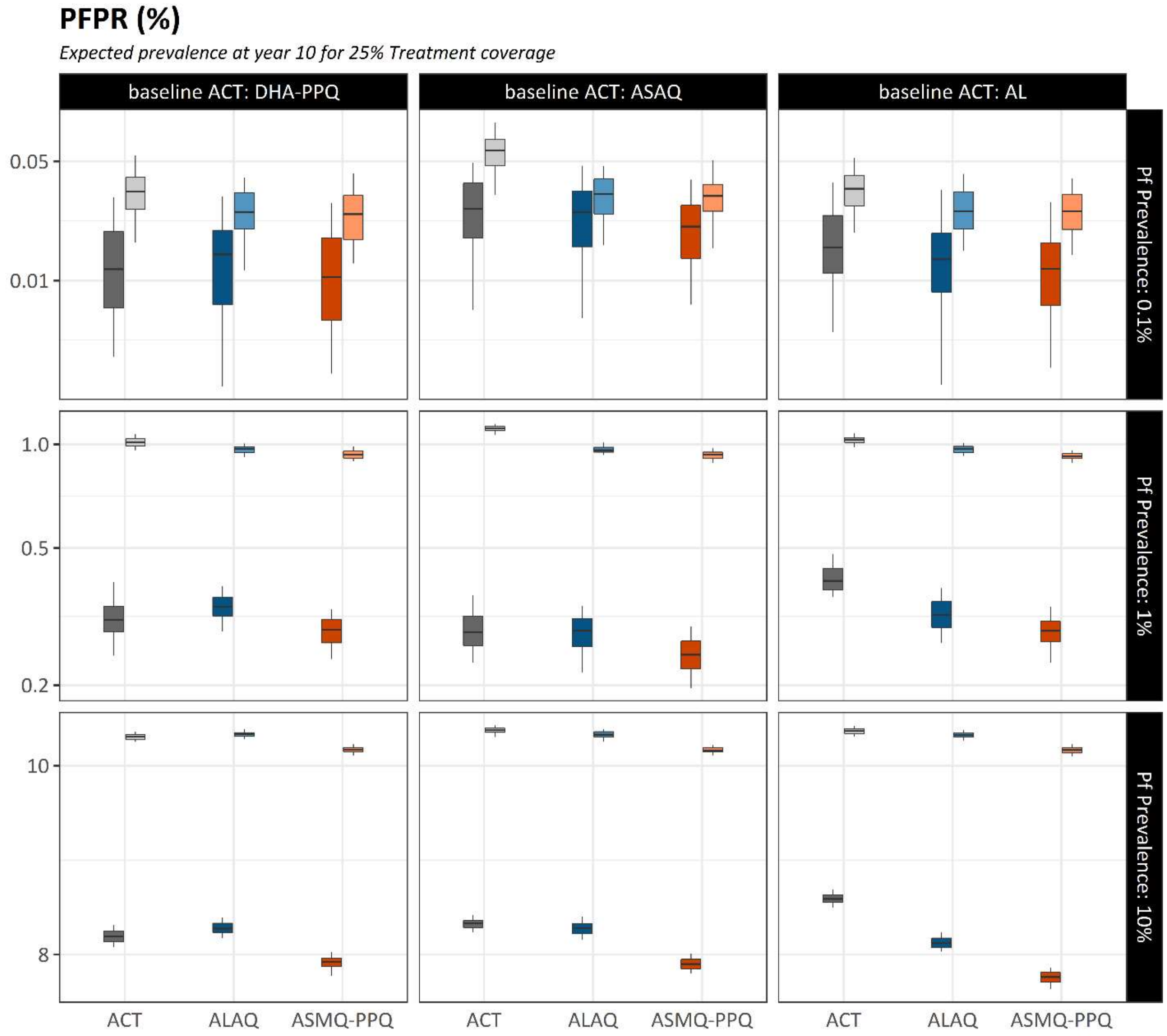

**Figure S28.** Pf prevalence after 10 years of TACT deployment or ACT deployment, in three baseline scenarios of ACT use (columns) and three prevalence settings (rows). Treatment coverage is 75%; The leftmost pair of boxplots in each panel (in gray) show Pf prevalence ten years later under a status quo ACT policy. The blue (ALAQ) and red (ASMQ-PPQ) boxplots show prevalence outcomes after 10 years of a TACT policy. Boxplot pairs have MORU model results on the left and PSU model results on the right. All boxplots summarize 100 simulations.

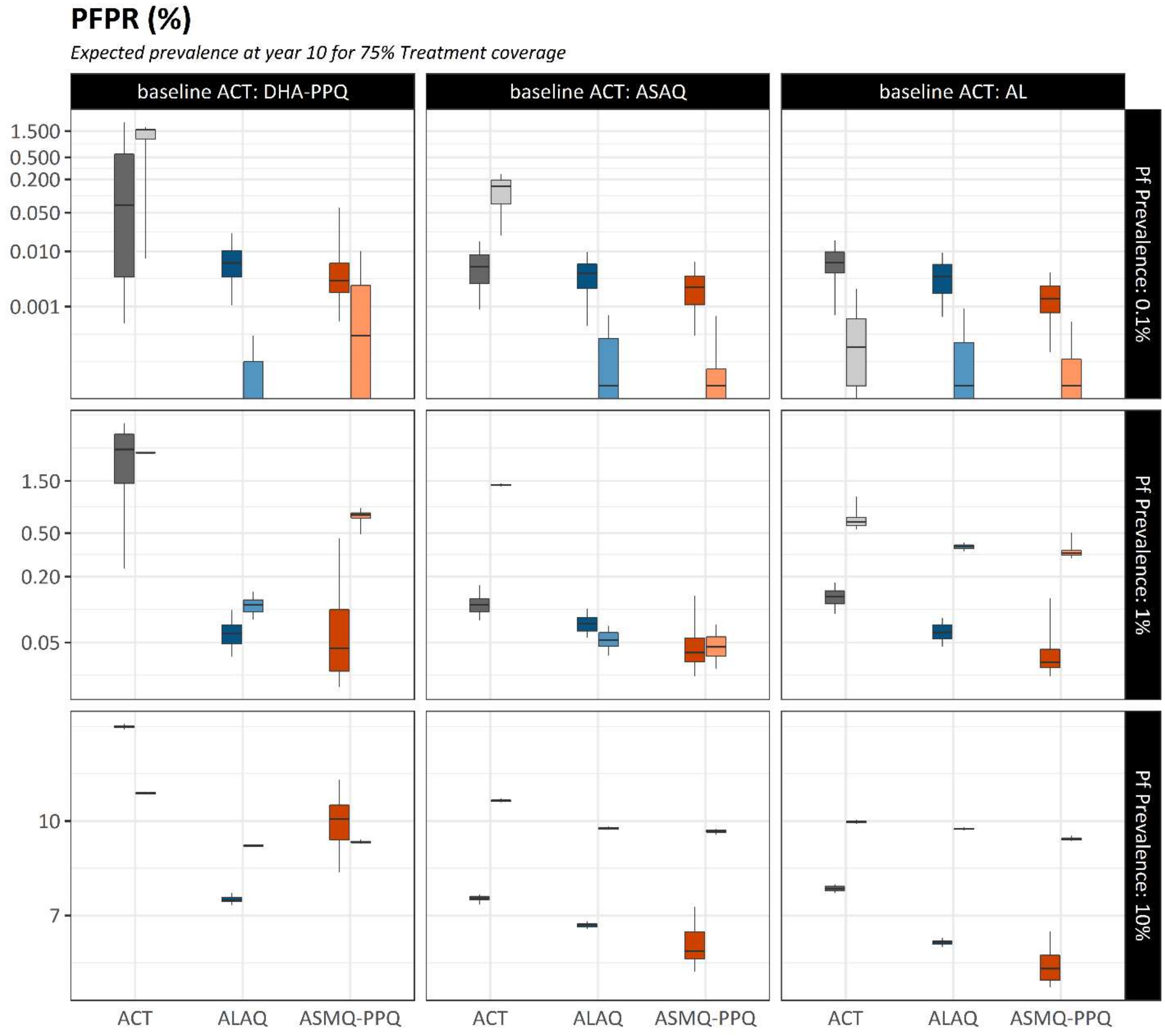

**Figure S29. Expected 580Y frequency dynamics for continued ACT use.** Each line represents the median 580Y frequency over a 10-year period with continued use of the ACT regimen identified by each column in a setting with Pf prevalence identified in each row. Shaded areas are bounded by the 25$^{th}$ and 75$^{th}$ percentiles obtained from 100 simulations for each scenario. Treatment coverage is set to 50%.

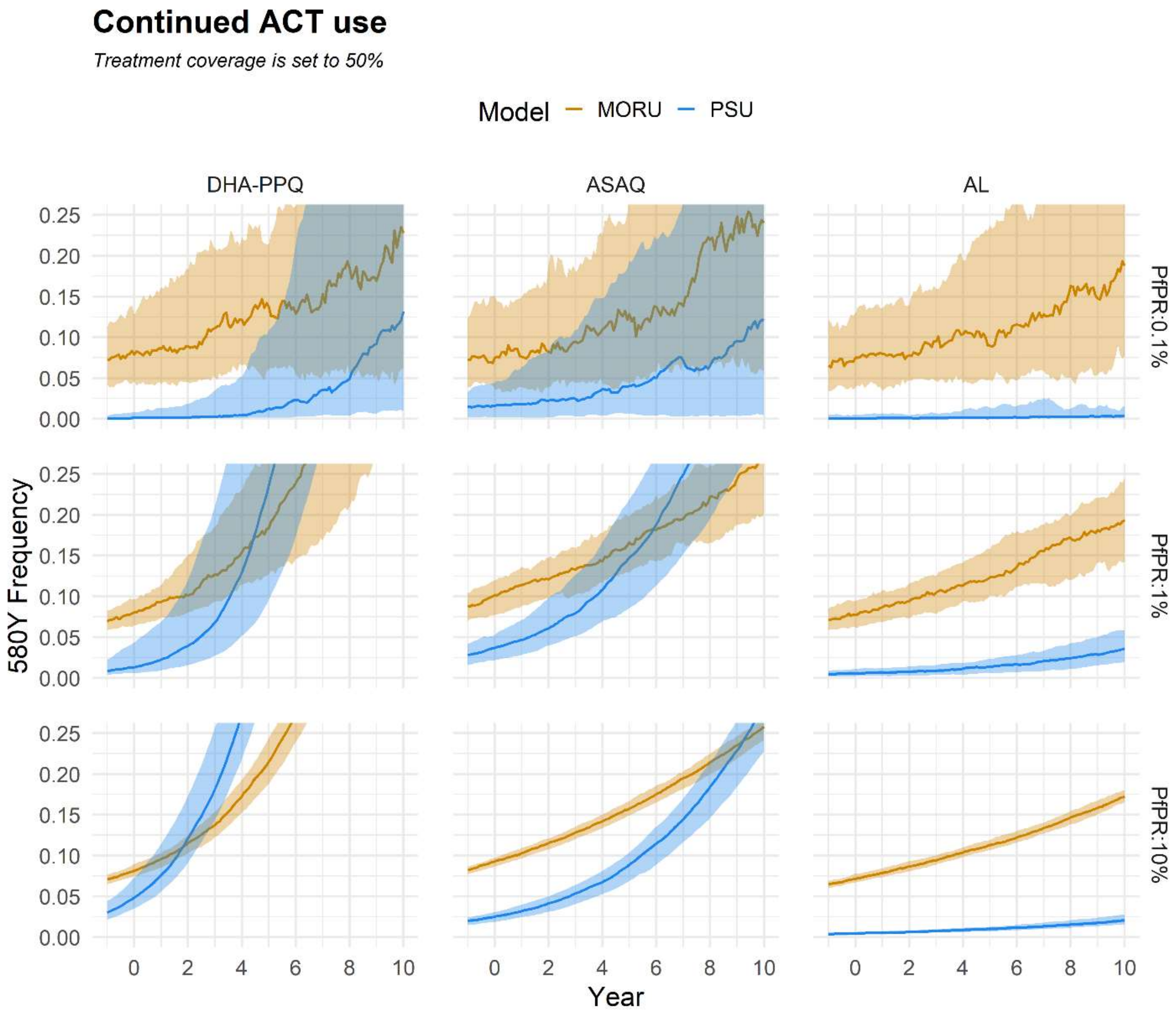

**Figure S30. Expected 580Y frequency dynamics for continued ACT use.** Each line represents the median 580Y frequency over a 10-year period with continued use of the ACT regimen identified by each column in a setting with Pf prevalence identified in each row. Shaded areas are bounded by the 25th and 75th percentiles obtained from 100 simulations for each scenario. Treatment coverage is set to 25%.

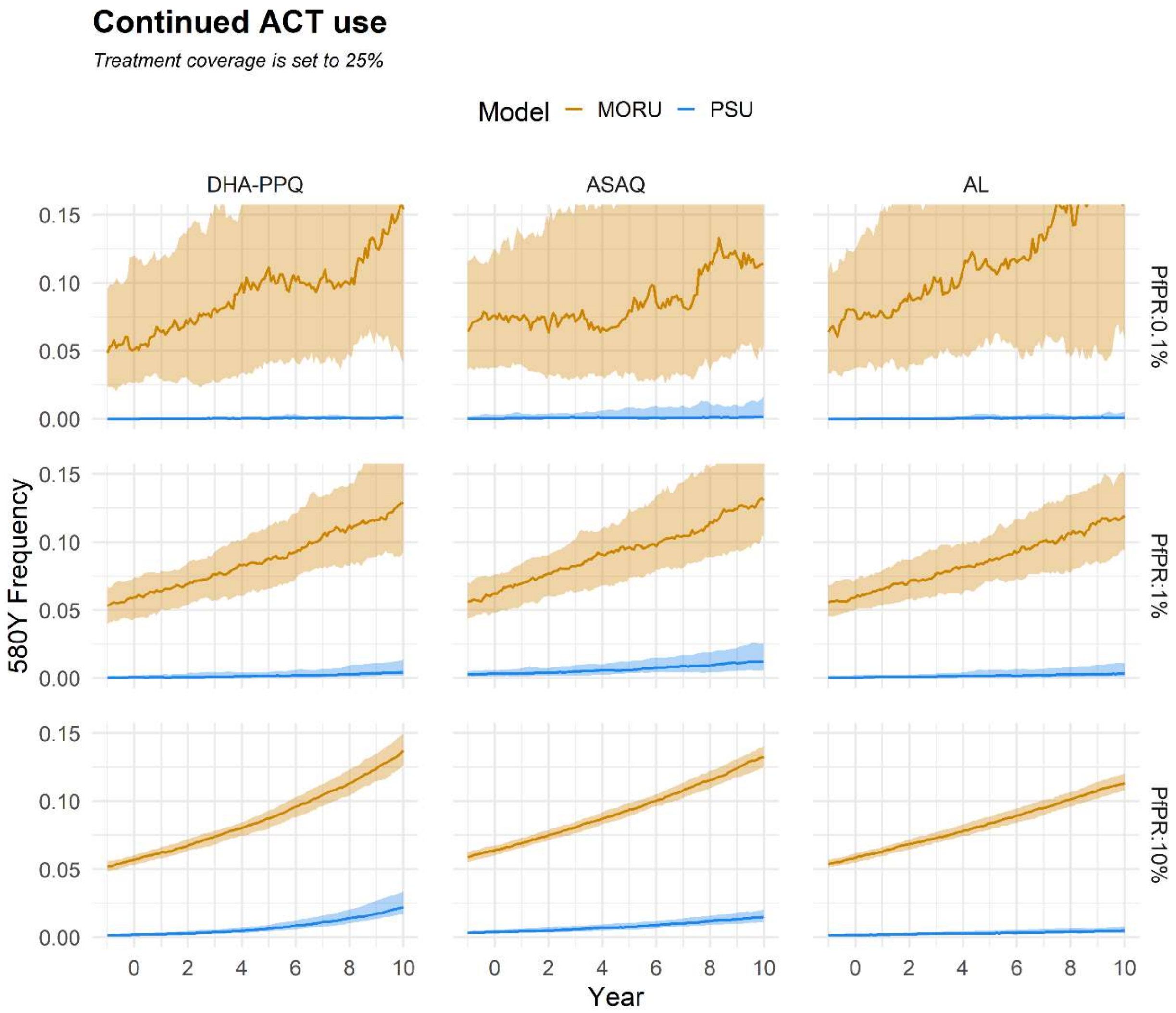

**Figure S31. Expected 580Y frequency dynamics for continued ACT use.** Each line represents the median 580Y frequency over a 10-year period with continued use of the ACT regimen identified by each column in a setting with Pf prevalence identified in each row. Shaded areas are bounded by the 25th and 75th percentiles obtained from 100 simulations for each scenario. Treatment coverage is set to 75%.

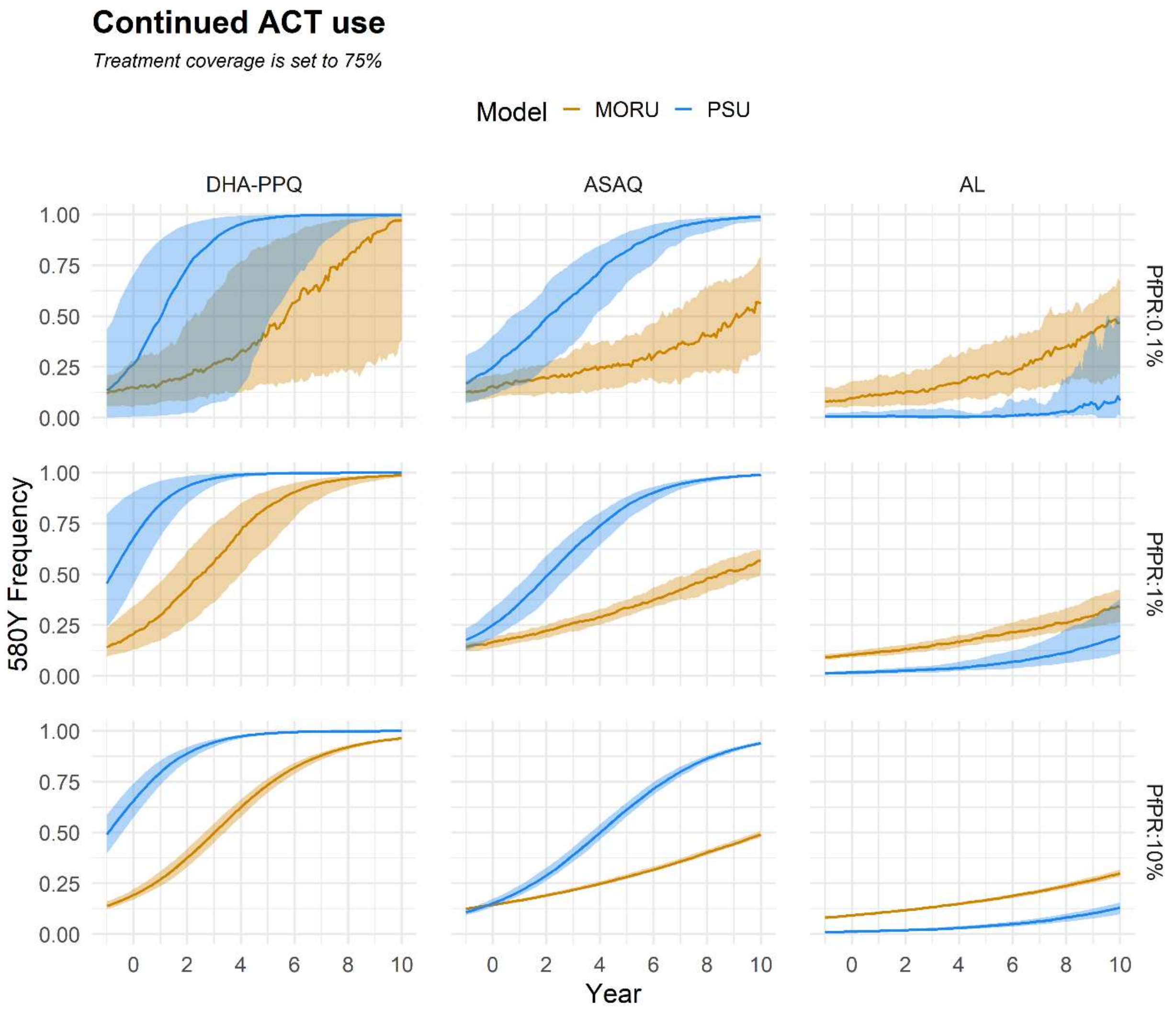

**Figure S32. Expected treatment failure dynamics for continued ACT use.** Each line represents the median treatment failure rate over a 10-year period with continued use of the ACT regimen identified by each column in a setting with Pf prevalence identified in each row. Shaded areas are bounded by the 25th and 75th percentiles obtained from 100 simulations for each scenario. Treatment coverage is set to 25%.

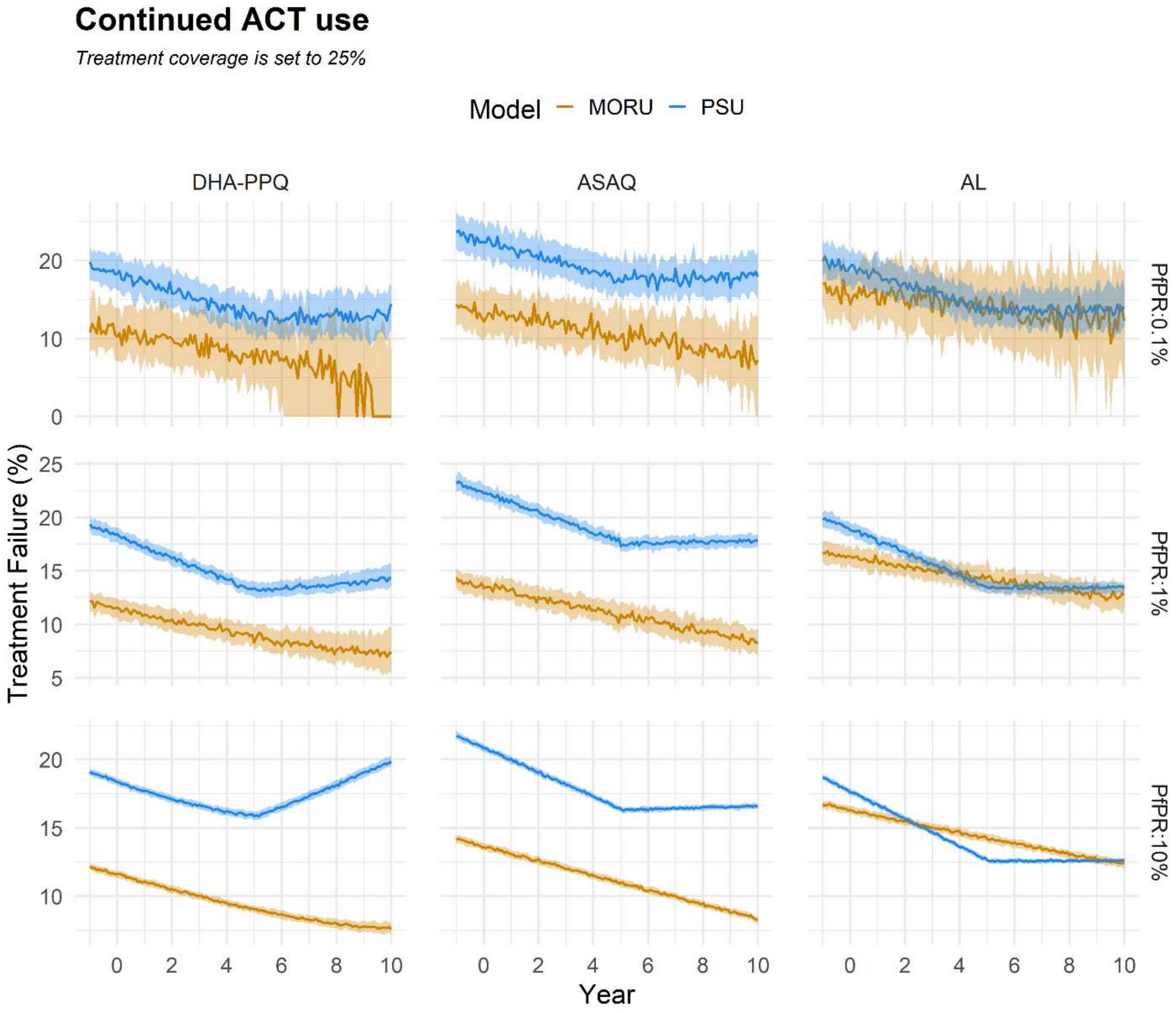

**Figure S33. Expected treatment failure dynamics for continued ACT use.** Each line represents the median treatment failure rate over a 10-year period with continued use of the ACT regimen identified by each column in a setting with Pf prevalence identified in each row. Shaded areas are bounded by the 25th and 75th percentiles obtained from 100 simulations for each scenario. Treatment coverage is set to 75%.

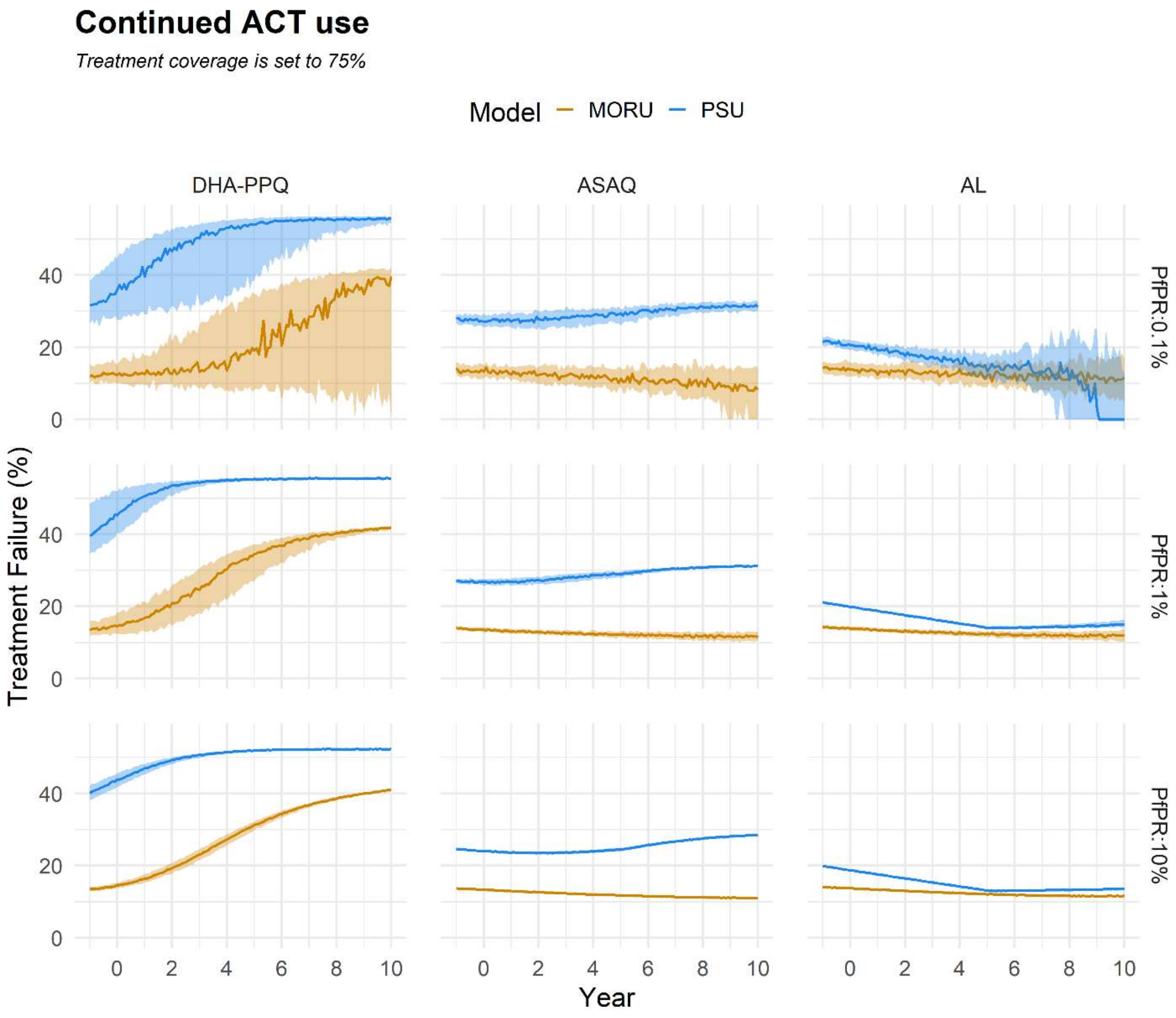

**Figure S34. Expected evolutionary dynamics of each monitored allele with ACT or TACT use.** Each column determines the drug regimen that is put in place for a 10-year period. Both TACTs explored here (ALAQ and ASMQ-PPQ) are compared to DHA-PPQ. Moderate selection pressure is defined by a combination of 0.1% Pf prevalence and 50% treatment coverage, whereas high selection pressure refers to settings with 10% Pf prevalence alongside 75% treatment coverage. Each dotted line illustrates changes in median allelic frequency over time from a set of 100 simulations.

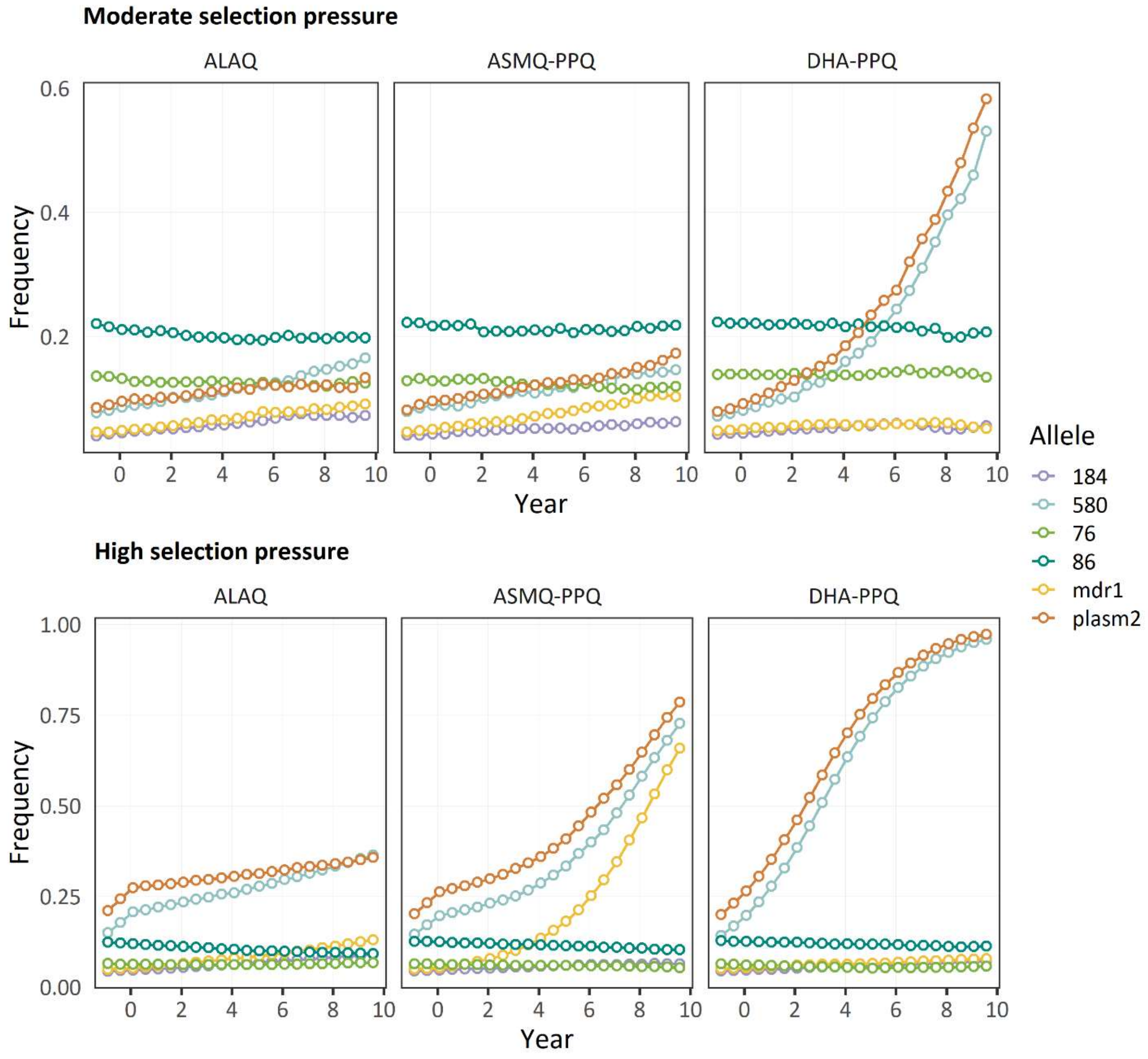

**Figure S35. Expected evolutionary dynamics of each monitored allele with ACT or TACT use.** Each column determines the drug regimen that is put in place for a 10-year period. Both TACTs explored here (ALAQ and ASMQ-PPQ) are compared to AL. Moderate selection pressure is defined by a combination of 0.1% Pf prevalence and 50% treatment coverage, whereas high selection pressure refers to settings with 10% Pf prevalence alongside 75% treatment coverage. Each dotted line illustrates changes in median allelic frequency over time from a set of 100 simulations.

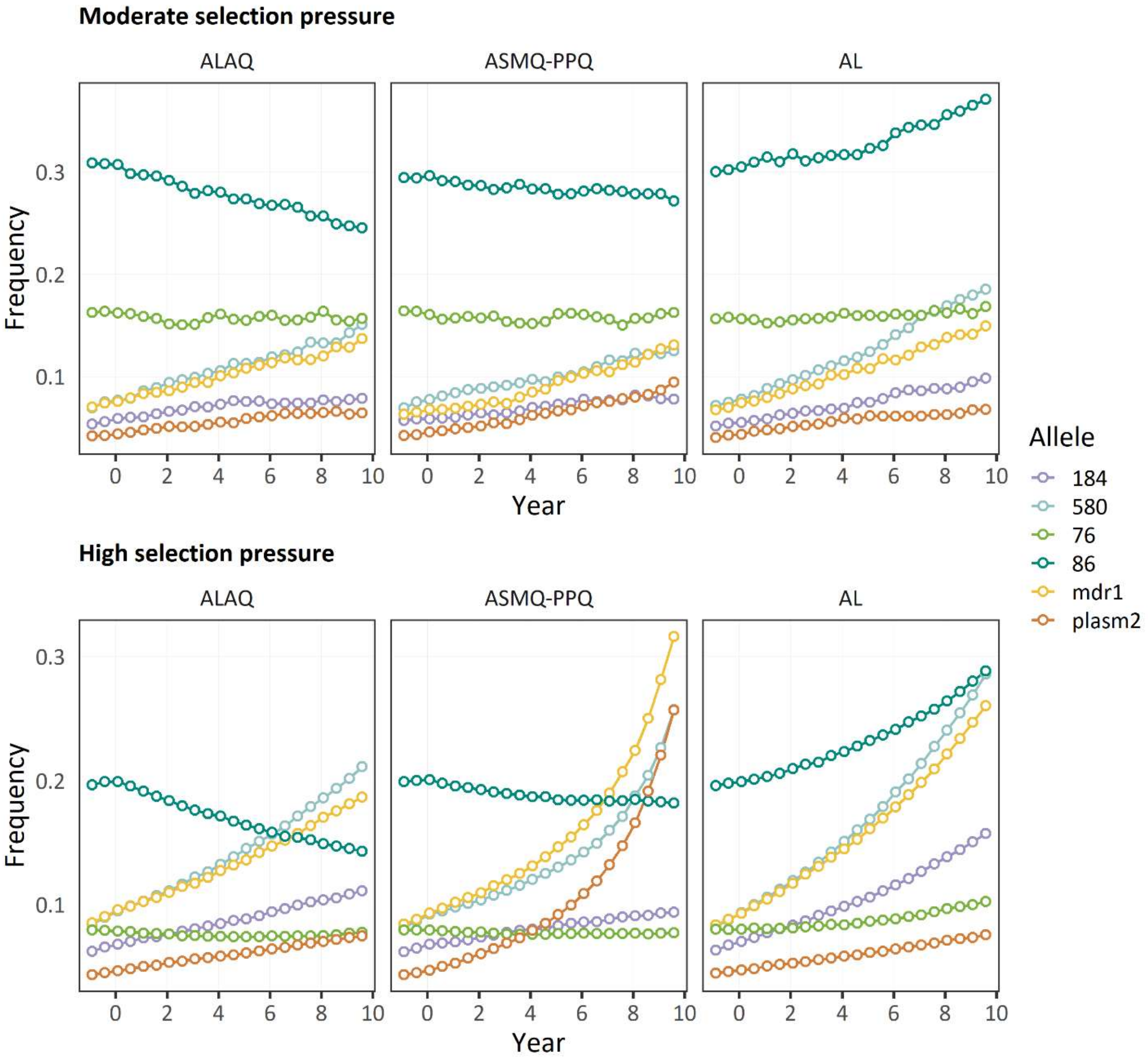

**Figure S36. Expected evolutionary dynamics of each monitored allele with ACT or TACT use.** Each column determines the drug regimen that is put in place for a 10-year period. Both TACTs explored here (ALAQ and ASMQ-PPQ) are compared to ASAQ. Moderate selection pressure is defined by a combination of 0.1% Pf prevalence and 50% treatment coverage, whereas high selection pressure refers to settings with 10% Pf prevalence alongside 75% treatment coverage. Each dotted line illustrates changes in median allelic frequency over time from a set of 100 simulations.

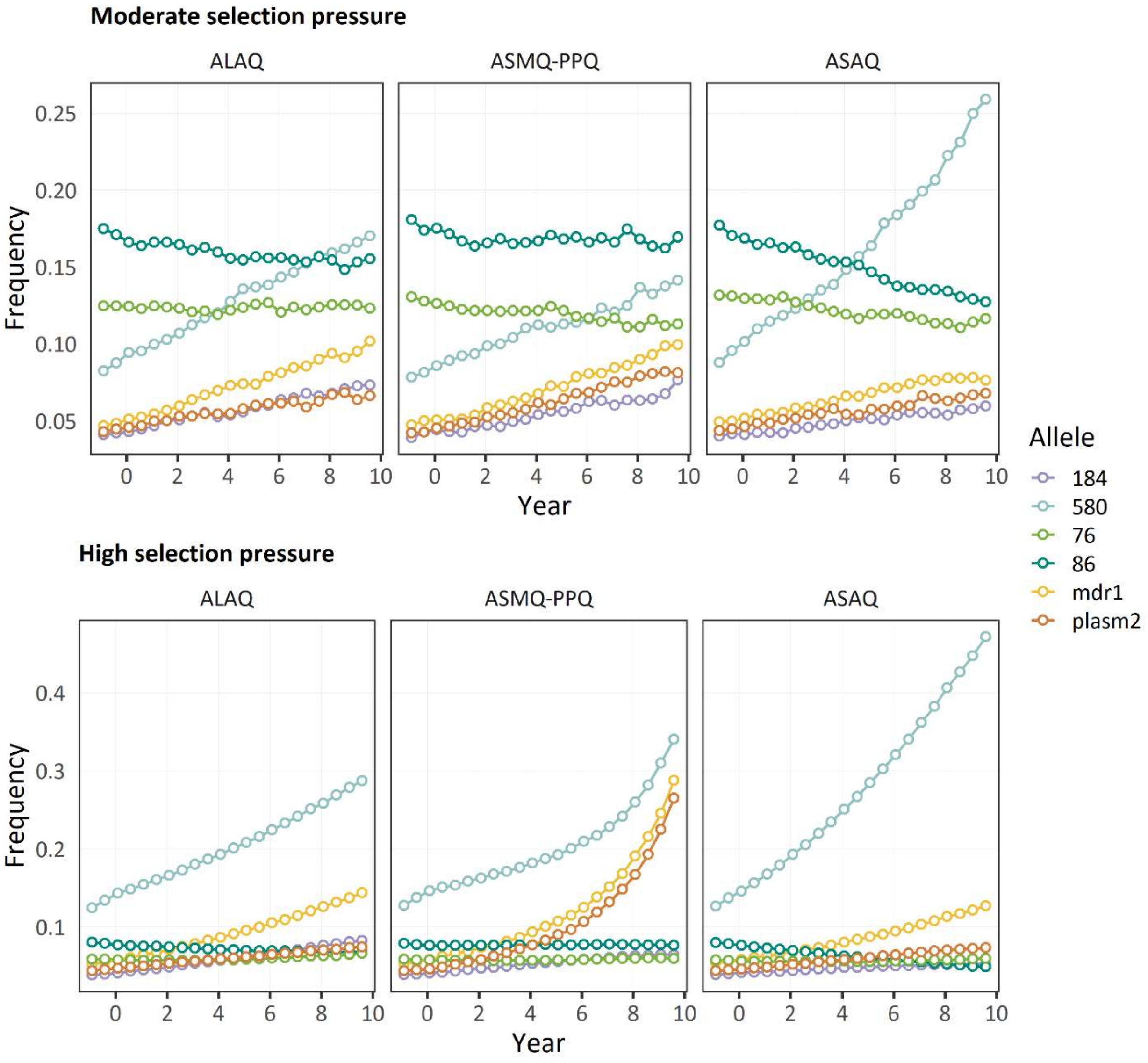

**Figure S37. Relationship between treatment failure rate and 580Y frequency**. Lines connect the median X-Y values for both treatment failure rate and 580Y over a 10-year period from a set of 100 simulations for settings with a given combination of ACT (column) and Pf prevalence (rows). Treatment coverage is set at 50%.

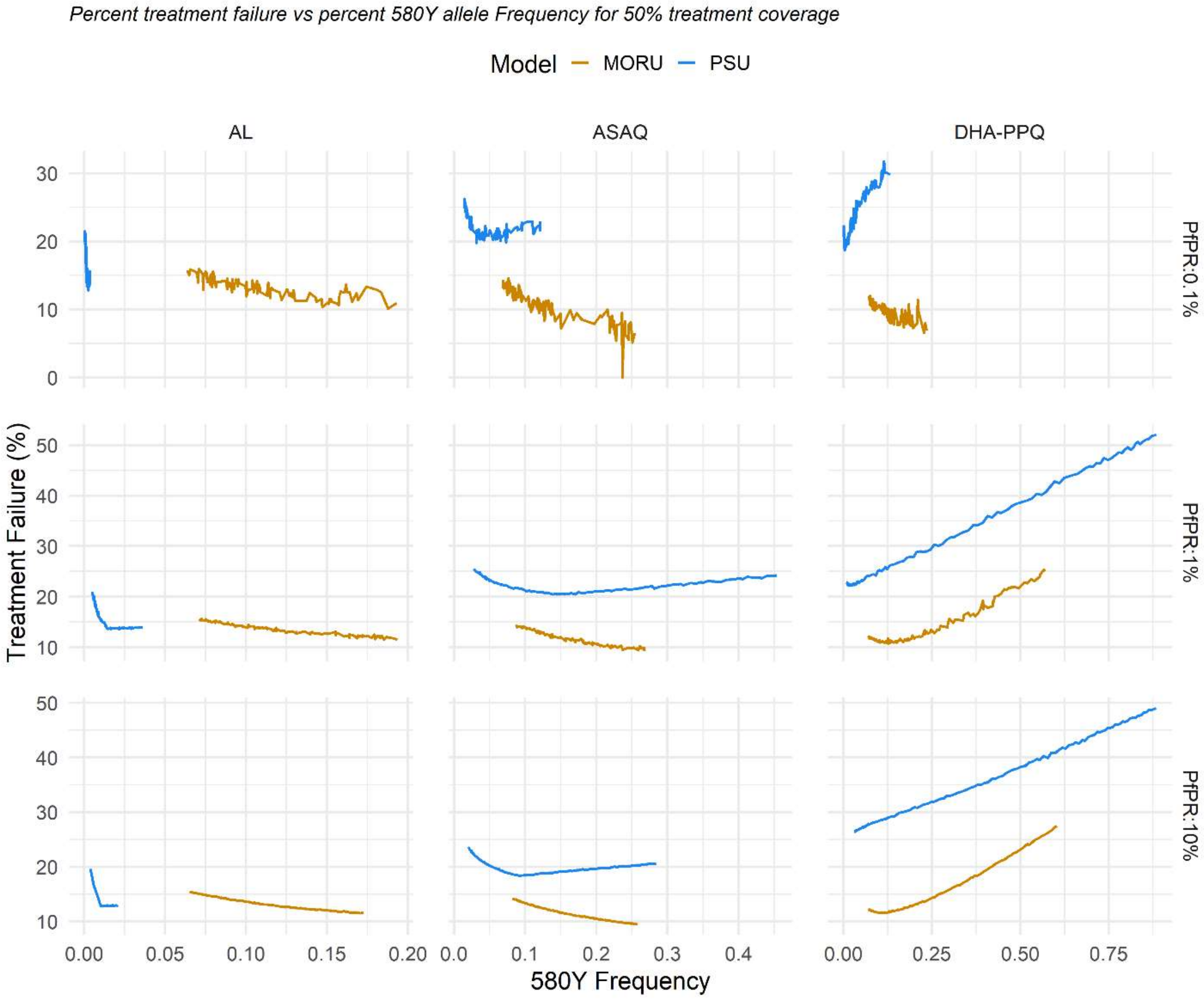

**Figure S38. Relationship between treatment failure rate and 580Y frequency**. Lines connect the median X-Y values for both treatment failure rate and 580Y over a 10-year period from a set of 100 simulations for settings with a given combination of ACT (column) and Pf prevalence (rows). Treatment coverage is set at 25%.

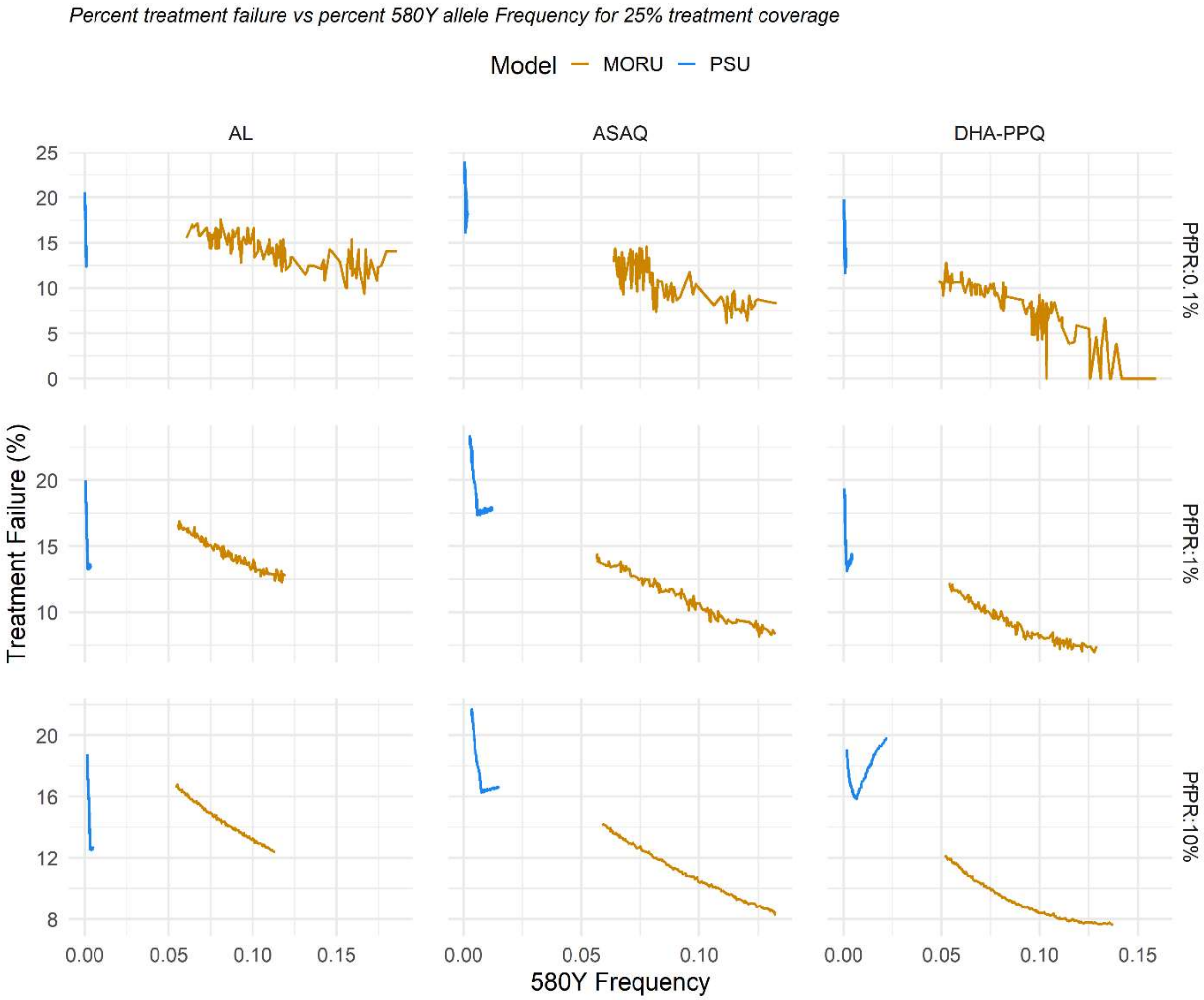

**Figure S39. Relationship between treatment failure rate and 580Y frequency**. Lines connect the median X-Y values for both treatment failure rate and 580Y over a 10-year period from a set of 100 simulations for settings with a given combination of ACT (column) and Pf prevalence (rows). Treatment coverage is set at 75%.

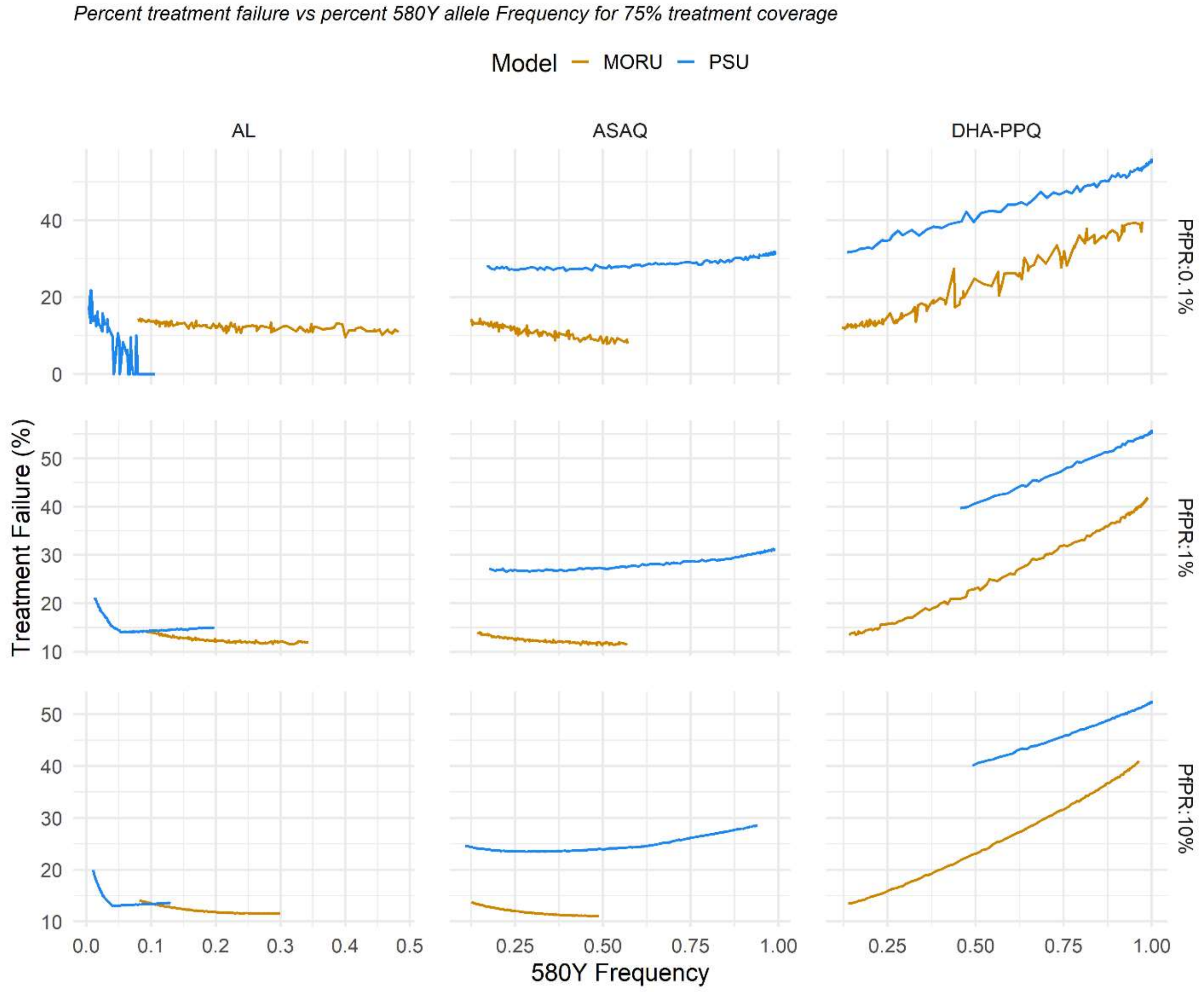

**Figure S40. Effects of delaying TACT introduction predicted by the MORU model**. In a population of 1,000,000 individuals, panels show the evolution of 580Y allele frequency over a period of 10 years under different transmission intensities: 0.1% PfPR (top row), 1% PfPR (middle row), 10% PfPR (bottom row). In these scenarios, treatment coverage is 50%, and DHA-PPQ was used as first-line ACT before switching to TACTs. The colors show the differences in delayed adoption of the TACT ranging from zero years delay (light yellow) to a five-year delay (black) delay. Left panels show results when switching from DHA-PPQ to ALAQ, while right panels show results when switching from DHA-PPQ to ASMQ-PPQ scenarios. The dots on the graph show the switch points for each trajectory; six lines are shown for delays of 0, 1, 2, 3, 4, and 5 years.

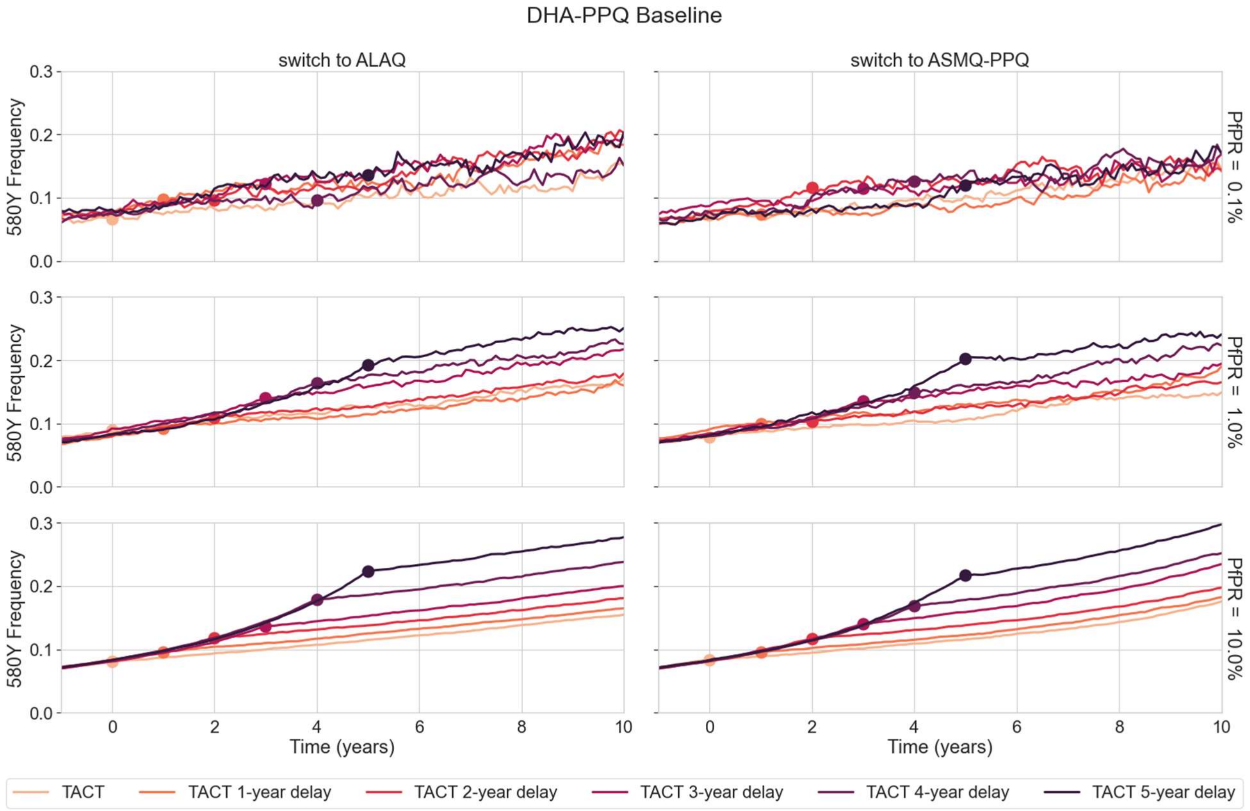

**Figure S41.** Same as Figure S40, with the results from PSU model.

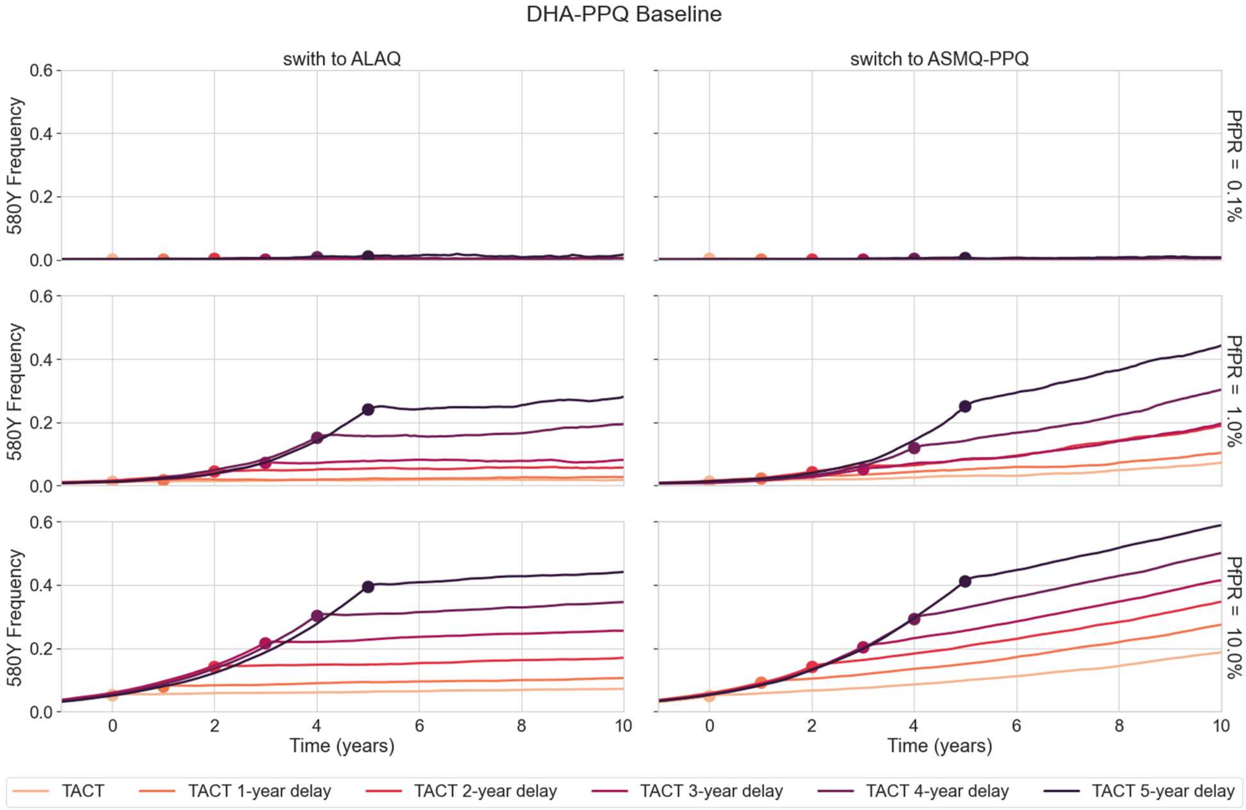

**Figure S42.** Same as Figure S40, with the results from MORU model. In these scenarios, ASAQ was used as first-line ACT before switching to TACTs.

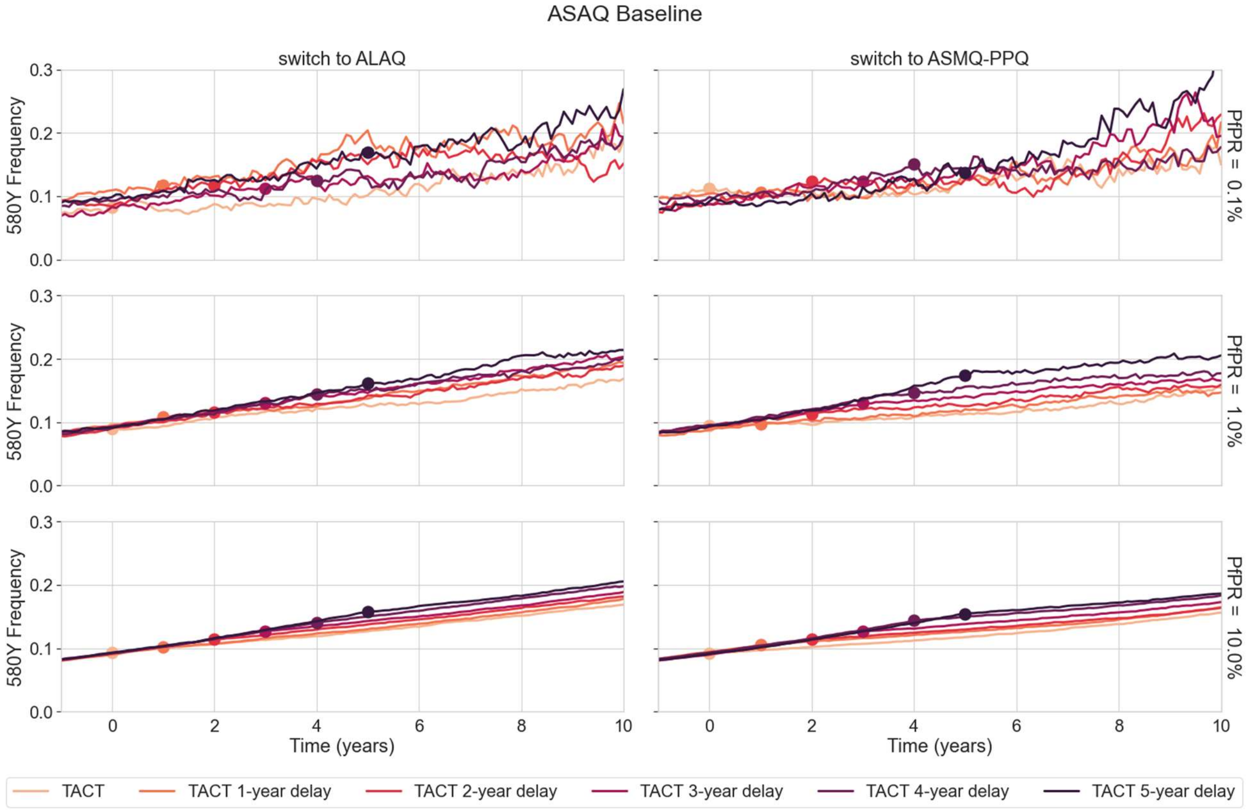

**Figure S43.** Same as Figure S42, with the results from PSU model.

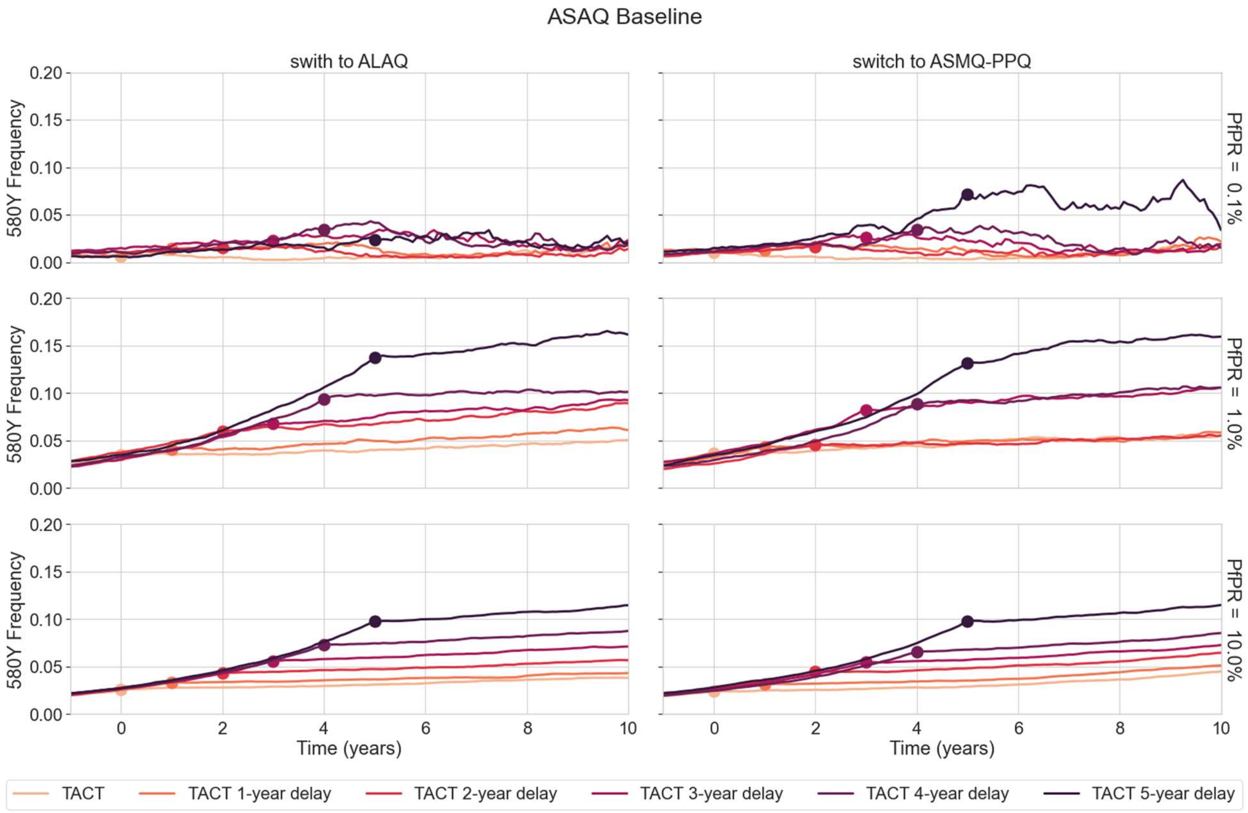

**Figure S44.** Same as Figure S40, with the results from MORU model. In these scenarios, AL was used as first-line ACT before switching to TACTs.

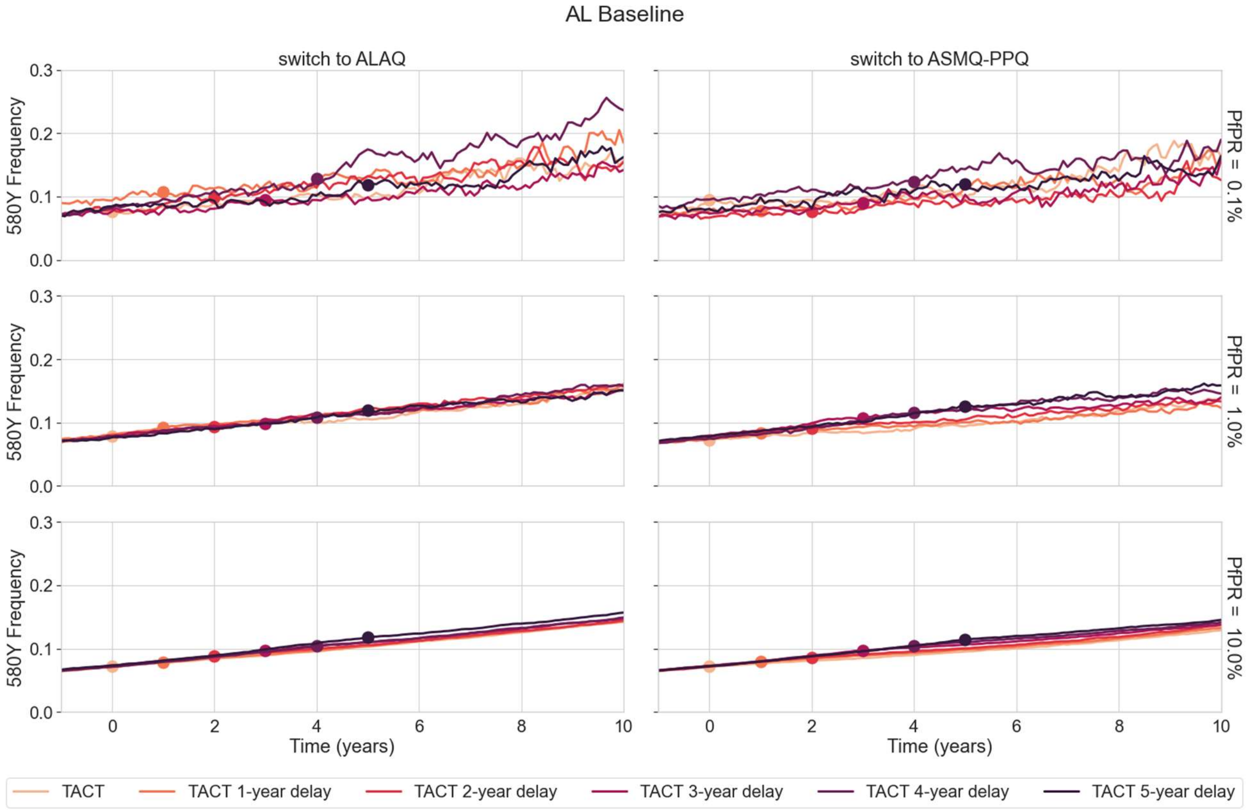

**Figure S45.** Same as Figure S44, with the results from PSU model.

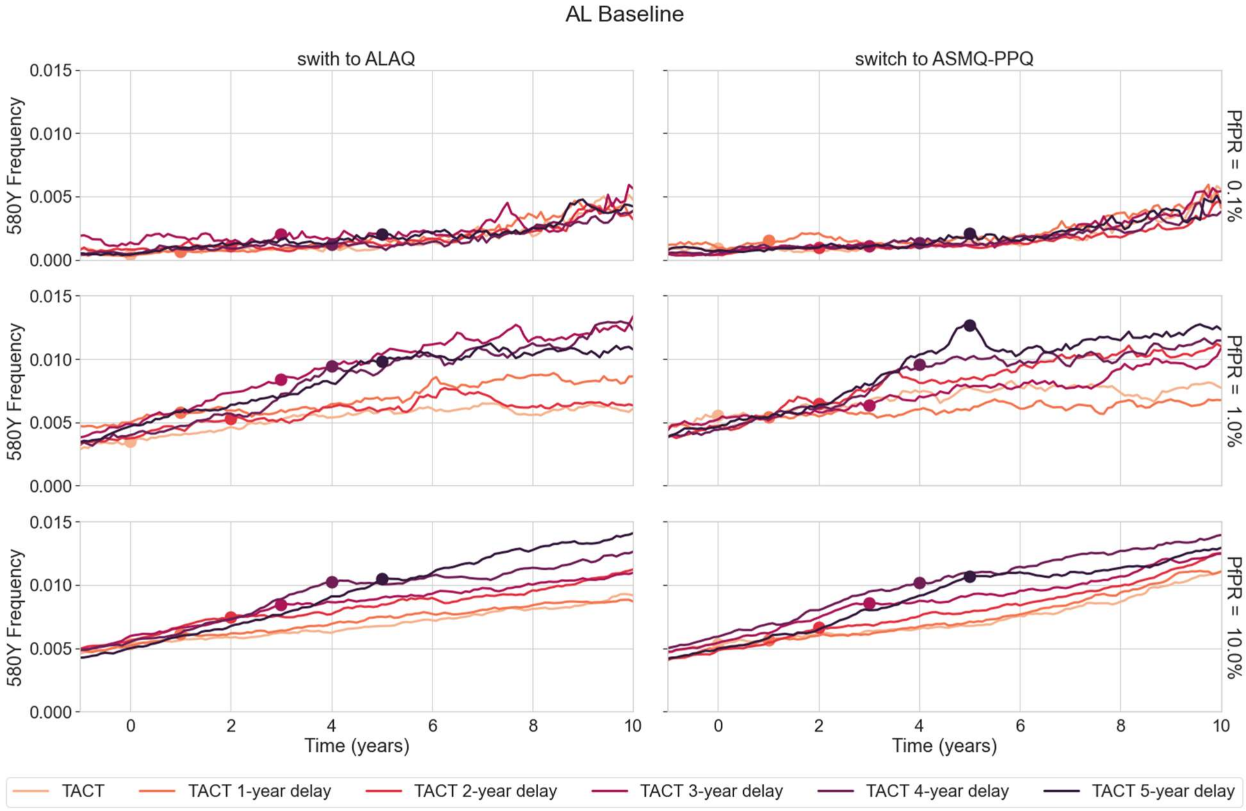

**Figure S46.** Same with Figure S40, with results from MORU model. The panels show the treatment failures rate over a period of 10 years. In those scenarios, DHA-PPQ was used as first-line ACT before switching to TACTs.

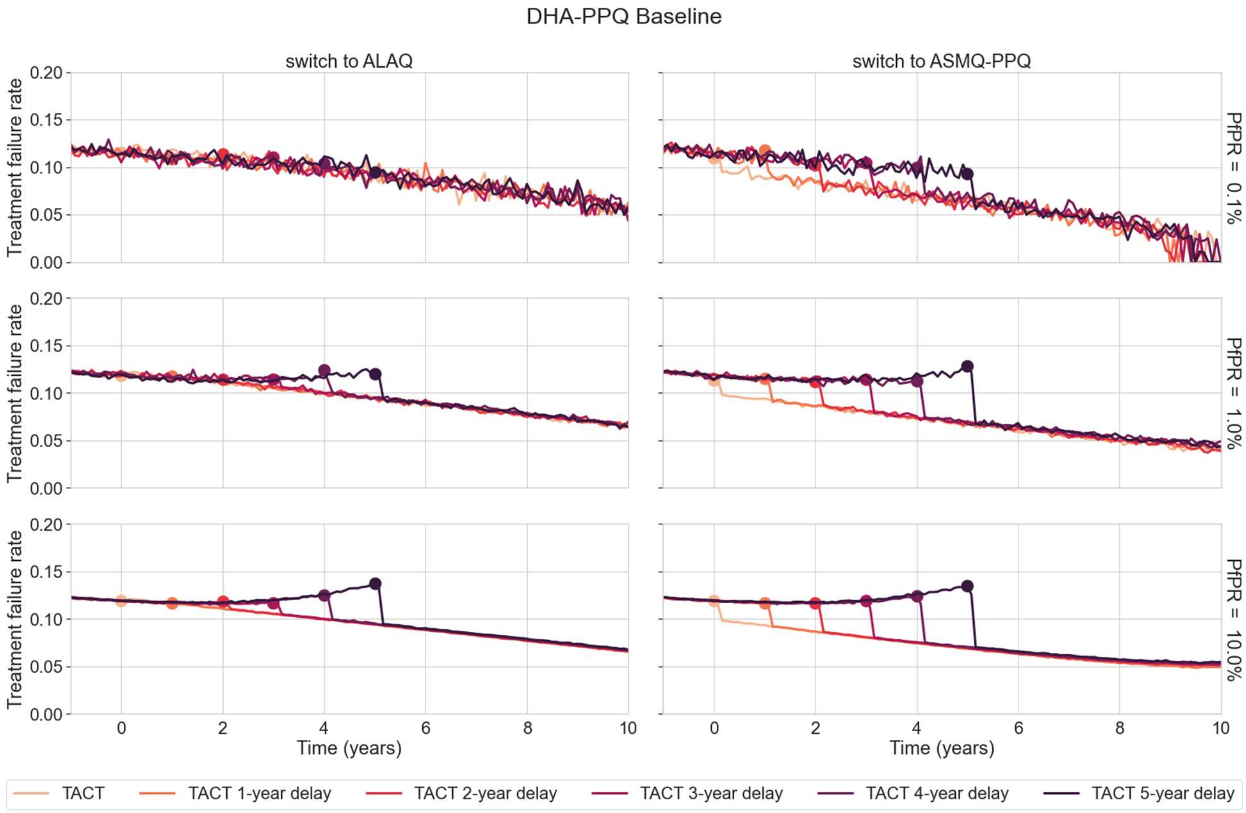

**Figure S47.** Same as Figure S46, with the results from PSU model.

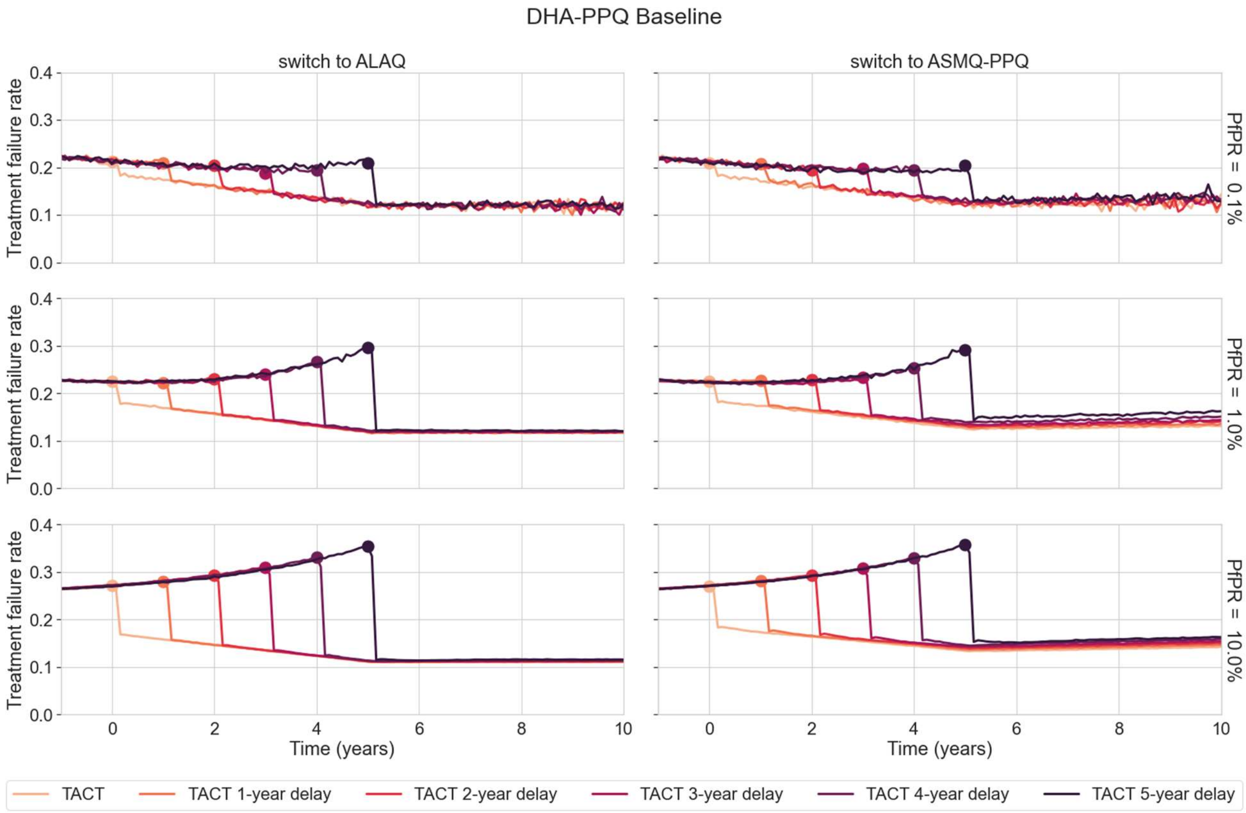

**Figure S48.** Same as Figure S46, with the results from MORU model. In these scenarios, ASAQ was used as first-line ACT before switching to TACTs.

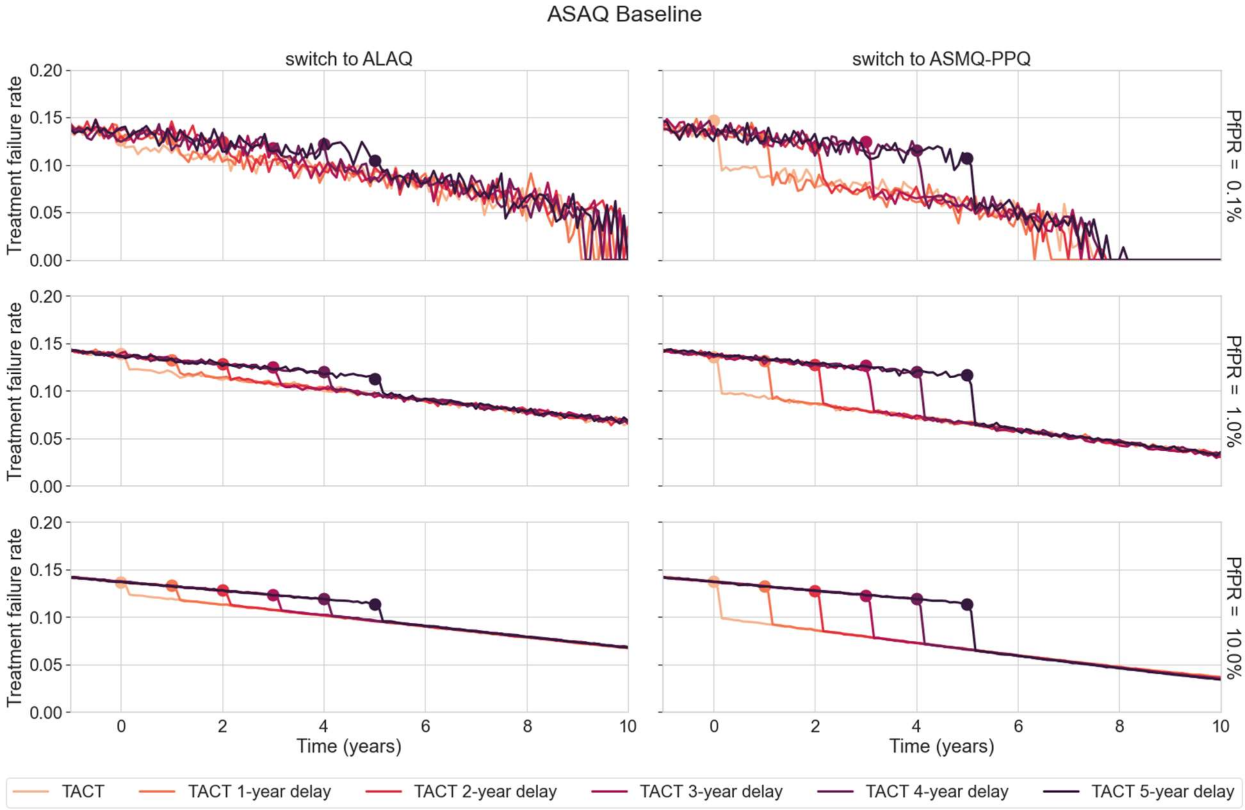

**Figure S49.** Same as Figure S48, with the results from PSU model.

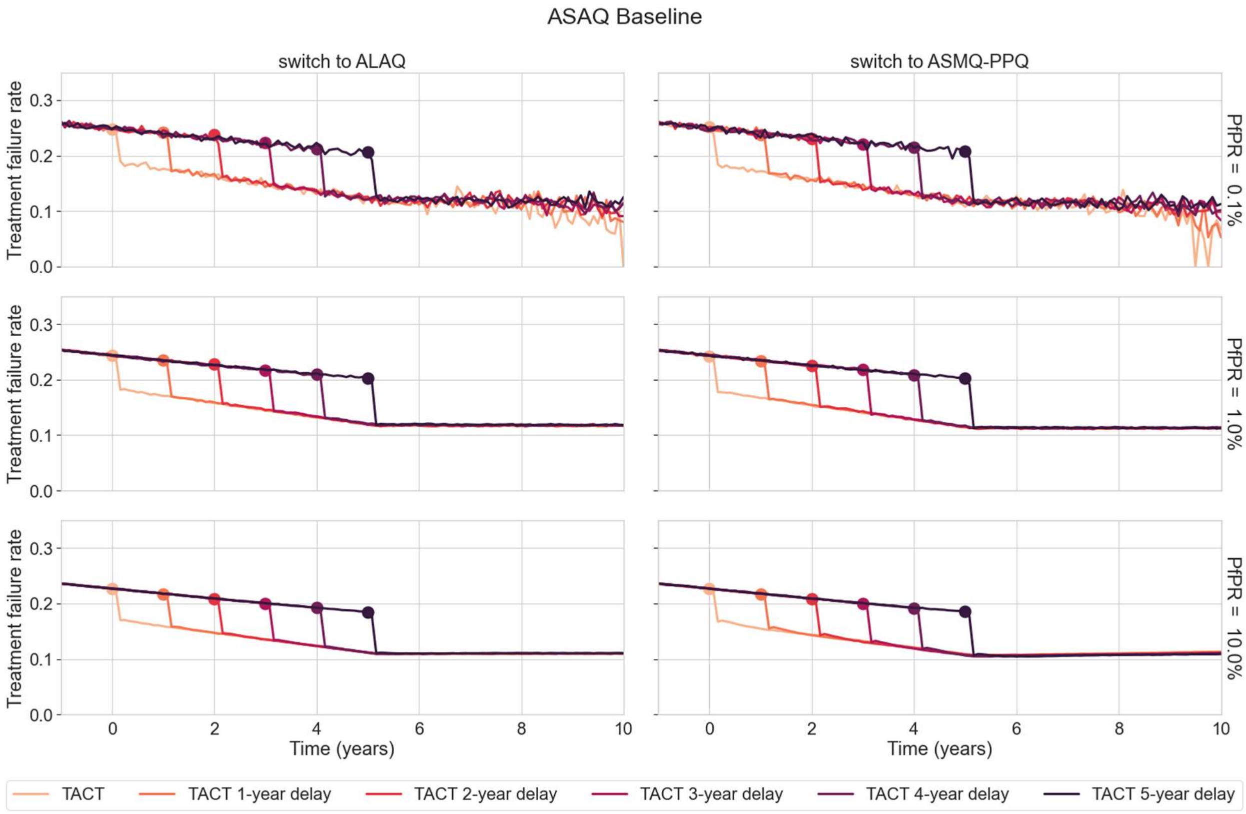

**Figure S50.** Same as Figure S46, with the results from MORU model. In these scenarios, AL was used as first-line ACT before switching to TACTs.

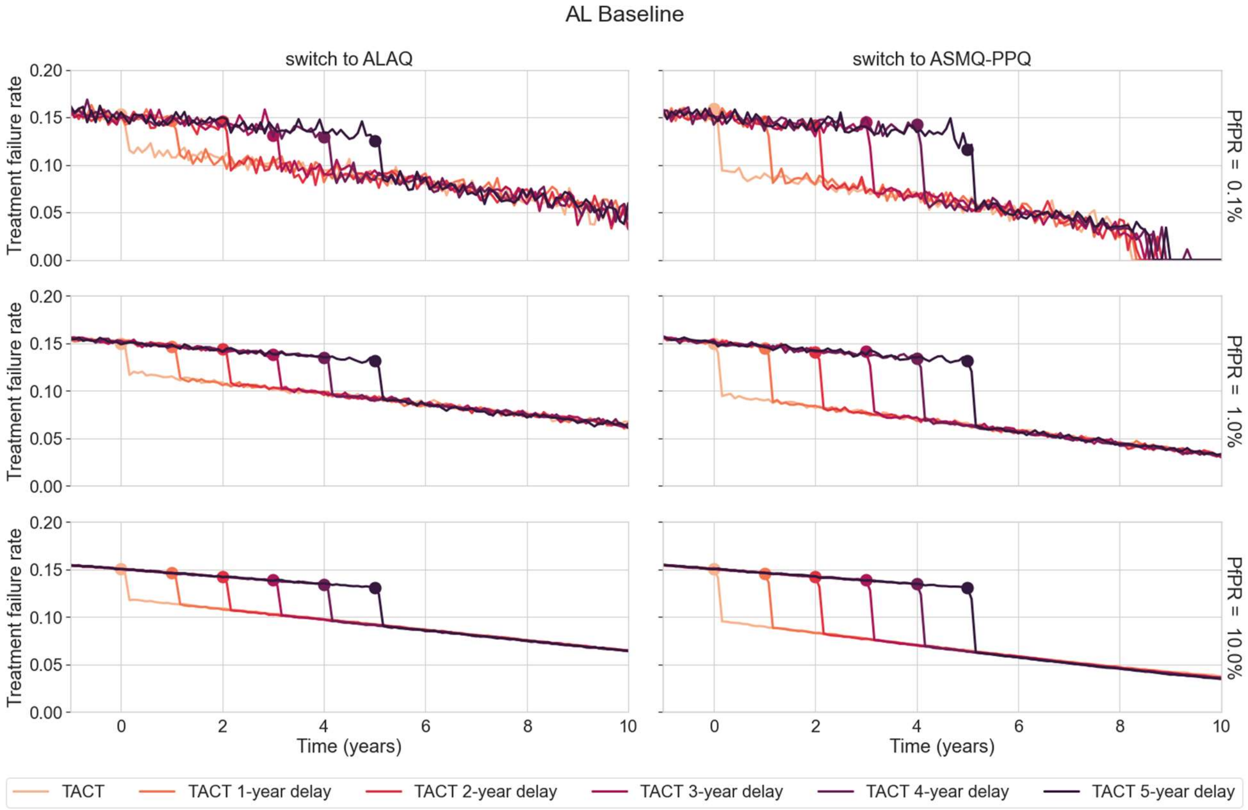

**Figure S51.** Same as Figure S50, with the results from PSU model.

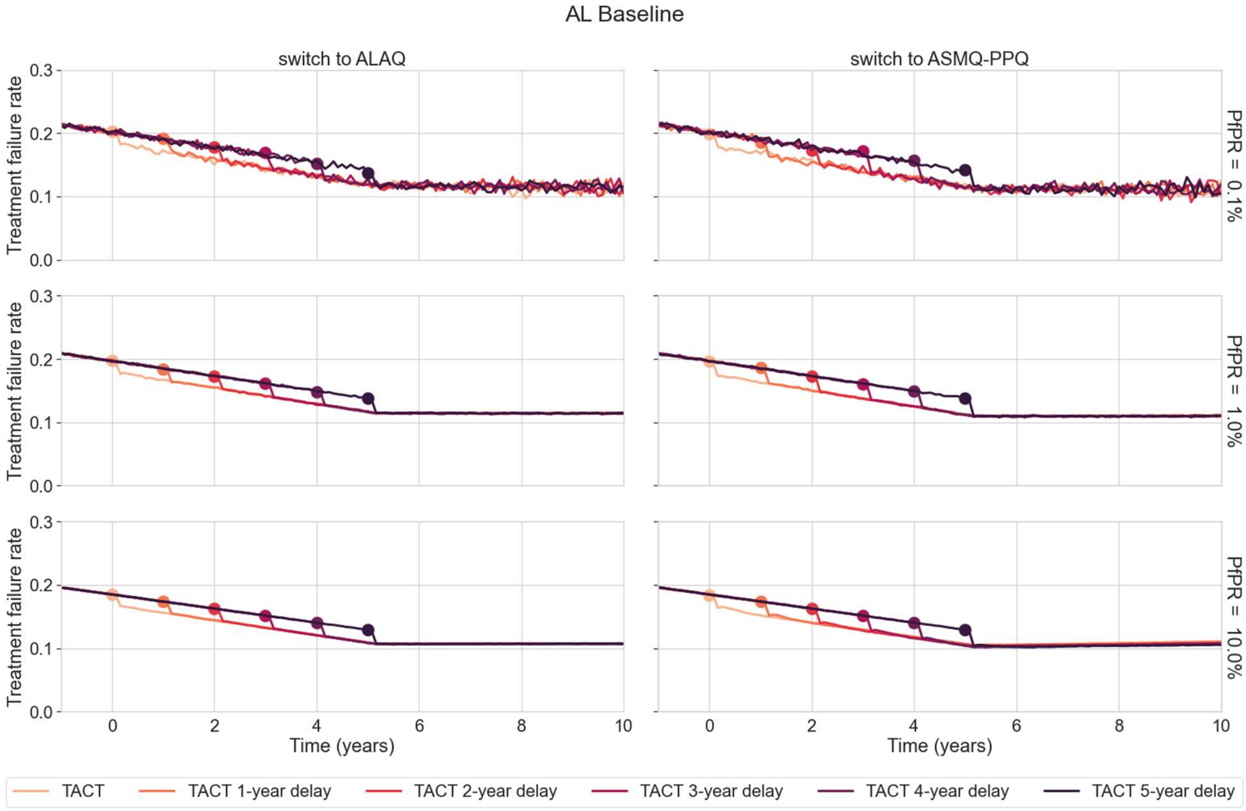

**Figure S52. Comparison of ACT and TACT deployment at 1% PfPR and 50% treatment coverage.** AL is used as the baseline ACT before TACTs are deployed at year zero; grey lines (medians from 100 simulations) show the evolution of the pfkelch13 580Y allele or treatment failure rates under continued DHA-PPQ use. Red lines show how these processes are slowed down by deployment of ASMQ-PPQ. Blue lines show how these processes are slowed down by deployment of ALAQ. All shaded areas show inter-quartile ranges. Panels above each graph show an individual's relative risk of 580Y infection (under TACT deployment versus ACT deployment) after 2, 5, and 10 years of deployment; bars show 95% confidence intervals assuming n=1000 for each deployment.

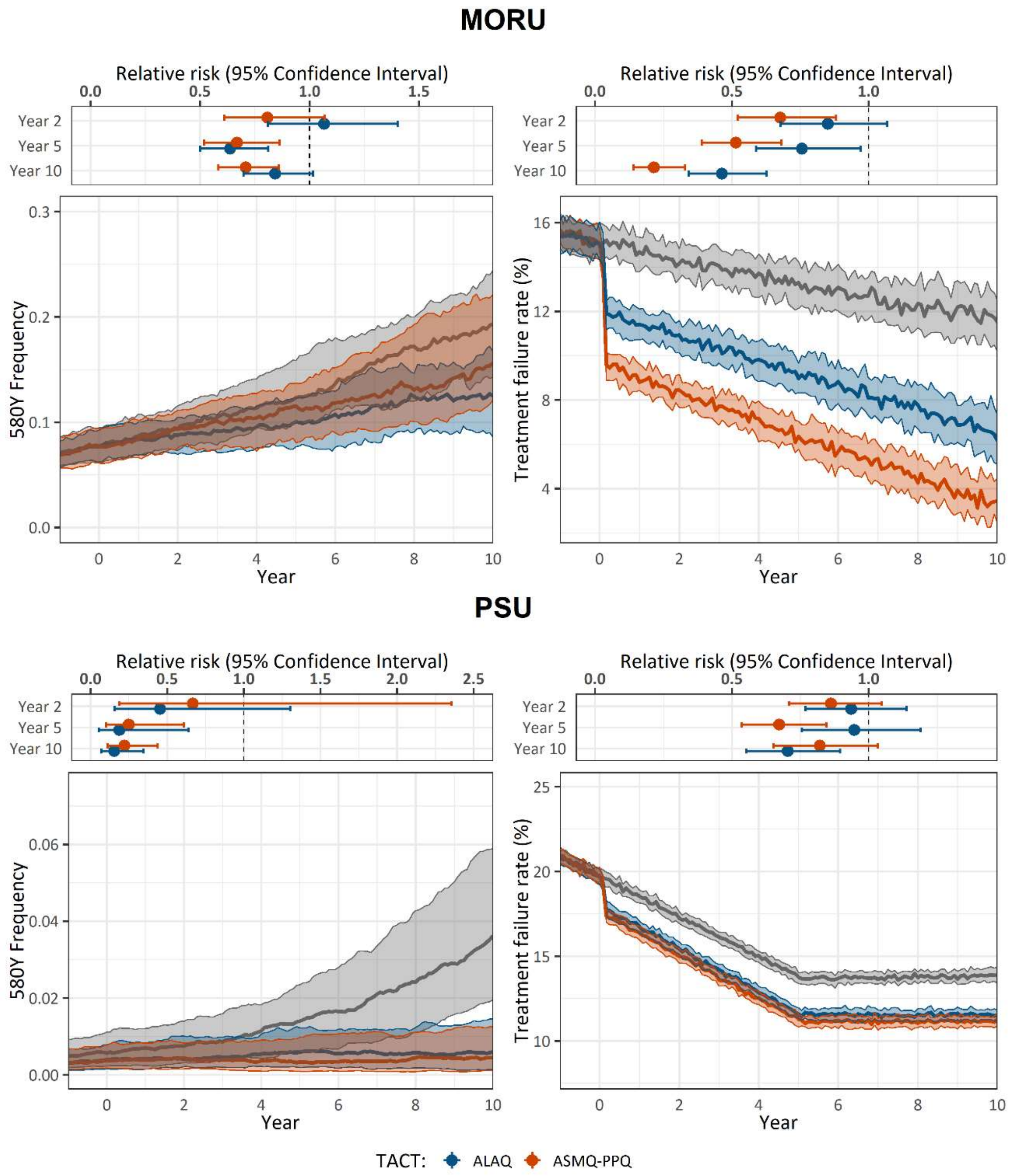

**Figure S53. Comparison of ACT and TACT deployment at 1% PfPR and 50% treatment coverage.** ASAQ is used as the baseline ACT before TACTs are deployed at year zero; grey lines (medians from 100 simulations) show the evolution of the pfkelch13 580Y allele or treatment failure rates under continued DHA-PPQ use. Red lines show how these processes are slowed down by deployment of ASMQ-PPQ. Blue lines show how these processes are slowed down by deployment of ALAQ. All shaded areas show inter-quartile ranges. Panels above each graph show an individual's relative risk of 580Y infection (under TACT deployment versus ACT deployment) after 2, 5, and 10 years of deployment; bars show 95% confidence intervals assuming n=1000 for each deployment.

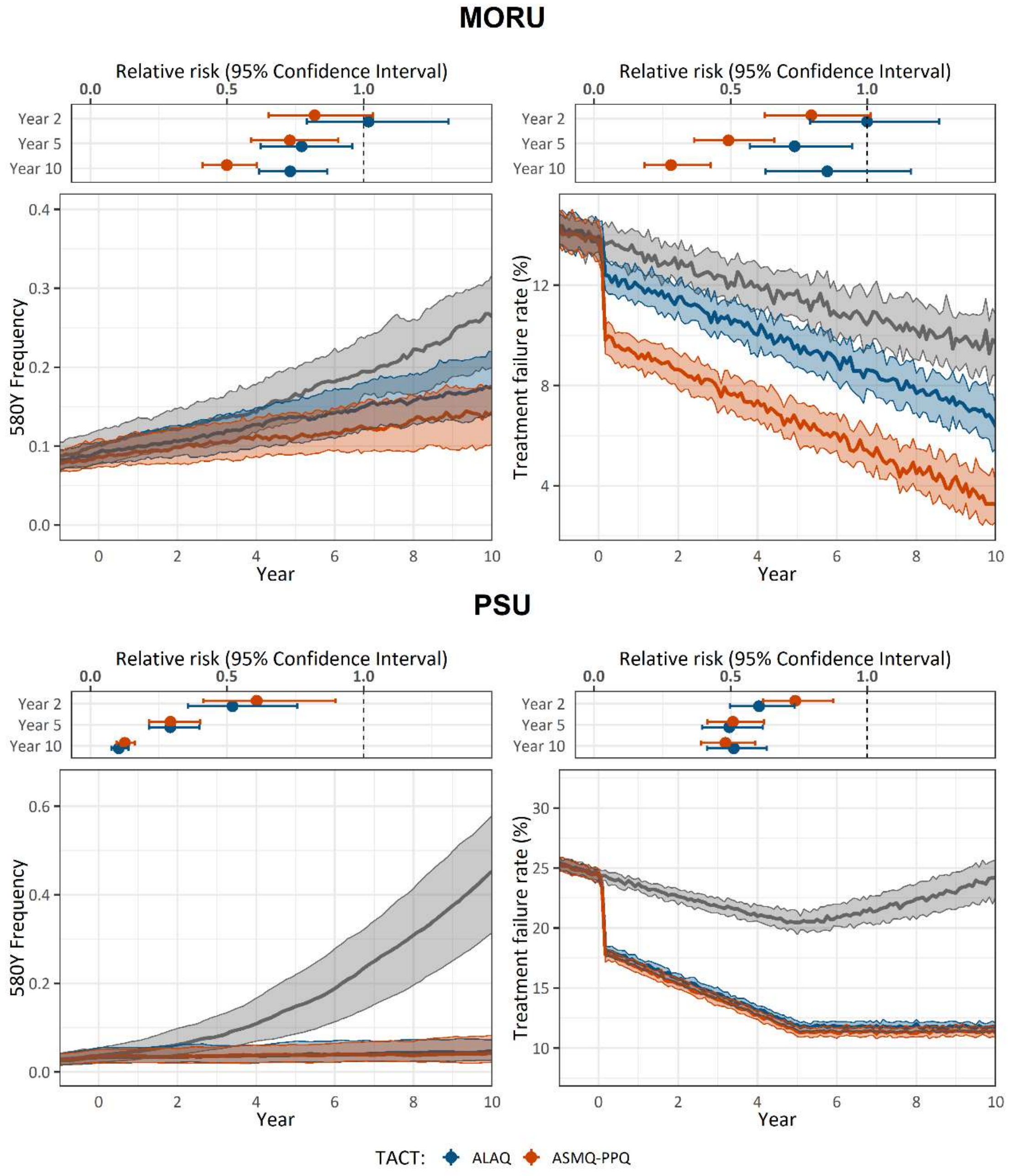

**Figure S54. Effect of gradual TACT adoption on 580Y frequency and treatment failure.**

In a population of 1,000,000 individuals, panels show the comparisons of 580Y allele frequency (top row) and treatment failure rate (bottom row) at year 10 under different transmission intensities: 0.1% PfPR (left column), 1% PfPR (middle column), 10% PfPR (right column). In these scenarios, DHAPPQ is used as the baseline therapy before TACTs starts to be adopted at year zero. The adoption of TACTs occurs in the public sector only with the initial proportion is 10% TACT and 90% regular ACT. Then, the adoption takes place over 3, 5, 7, or 9 years. Colors shows the TACTs used.

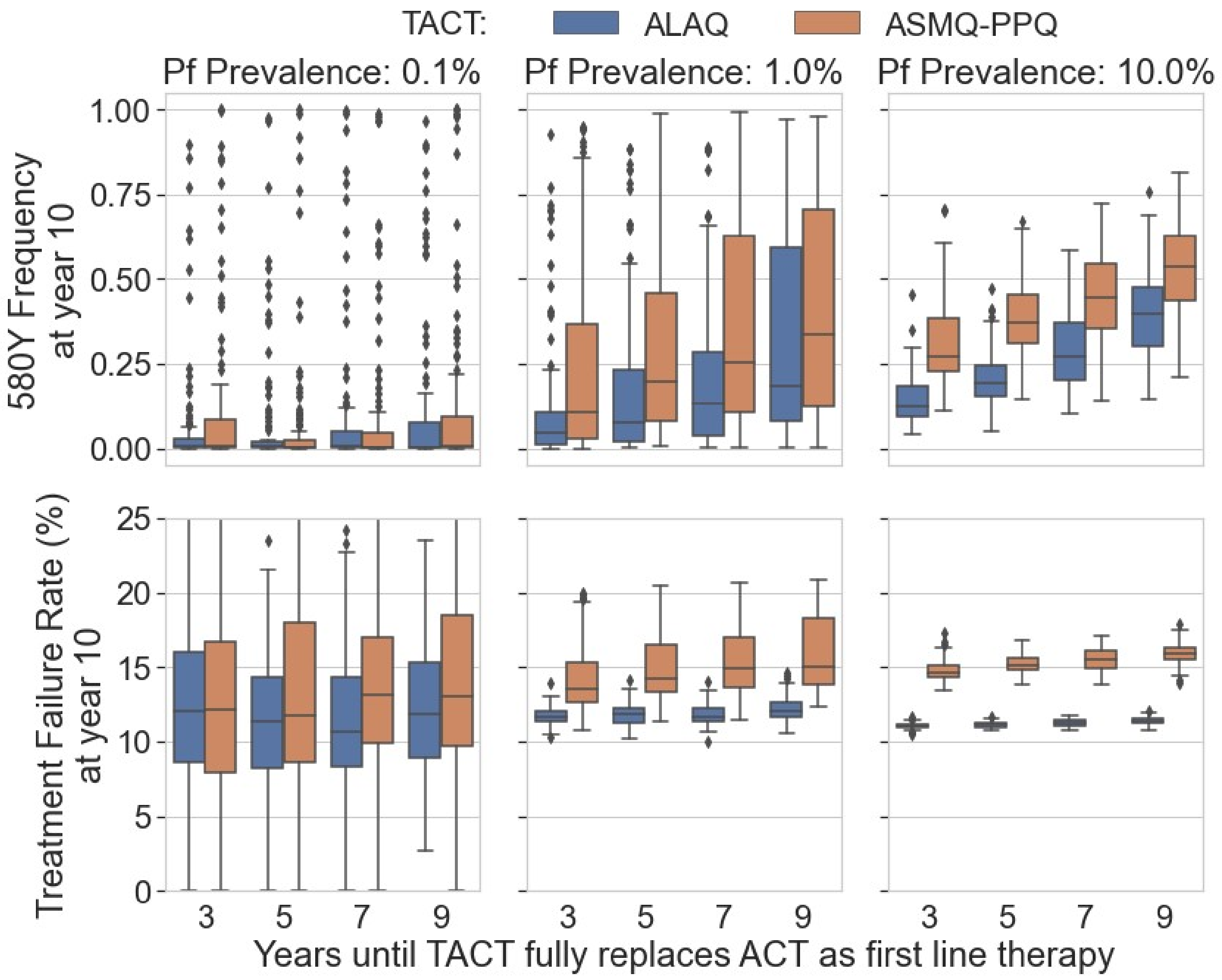
